# Approaches to improving mental health care for autistic people: a systematic review

**DOI:** 10.1101/2023.03.10.23287101

**Authors:** Sofia Loizou, Tamara Pemovska, Theodora Stefanidou, Una Foye, Ruth Cooper, Ariana Kular, Anna Greenburgh, Helen Baldwin, Jessica Griffiths, Katherine R. K. Saunders, Phoebe Barnett, Matilda Minchin, Gráinne Brady, Nafiso Ahmed, Jennie Parker, Beverley Chipp, Rachel Rowan Olive, Robin Jackson, Amanda Timmerman, Suzi Sapiets, Eva Driskell, Bethany Parsons, Debbie Spain, Vaso Totsika, Will Mandy, Richard Pender, Philippa Clery, Kylee Trevillion, Brynmor Lloyd-Evans, Alan Simpson, Sonia Johnson

**Author notes:** Corresponding author: Sofia Loizou. Contributed equally to the work and share first authorship (Sofia Loizou, Tamara Pemovska, Theodora Stefanidou).

## Abstract

**Background:** Autistic people have a high likelihood of developing mental health difficulties but a low chance of receiving effective mental health care. Therefore, there is a need to identify and examine strategies to improve mental health care for autistic people.

**Aims:** To identify strategies that have been implemented to improve access, experiences of care and mental health outcomes for autistic adults and examine evidence on their acceptability, feasibility and effectiveness.

**Method:** A co-produced systematic review was conducted. MEDLINE, PsycINFO, CINHAL, medRxiv and PsyArXiv were searched. We included all study designs reporting acceptability or feasibility outcomes and empirical quantitative study designs reporting effectiveness outcomes. Data were synthesised using a narrative approach.

**Results:** A total of 29 articles were identified. These included 16 studies of adapted mental health interventions, seven studies of service improvements and six studies of bespoke mental health interventions developed for autistic people. There was no conclusive evidence on effectiveness. However, most bespoke and adapted approaches appeared to be feasible and acceptable. Identified adaptations appeared to be acceptable and feasible, including increasing knowledge and detection of autism, providing environmental adjustments and communication accommodations, accommodating individual differences, and modifying the structure and content of interventions.

**Conclusion:** Many identified strategies are feasible and acceptable and can be readily implemented in services with the potential to make mental health care more suitable for autistic people, but important research gaps remain. Future research should address these and investigate a co-produced package of service improvement measures.

## Introduction

Autistic^1^ people (1) experience a high rate of mental health difficulties but are less likely to receive effective mental health support (2). For autistic people, co-occurring mental health difficulties can lead to negative outcomes including poor quality of life (3–5) and increased risk of suicide (6–8). Accessing appropriate support from mental health services can be a critical step towards addressing mental health difficulties in autistic people. Dissatisfaction with care (9), high levels of unmet needs (10), and harmful effects (11) suggest that mental health services do not currently provide sufficient support for many autistic individuals with co-occurring mental health difficulties. This can erode trust in services and prevent help-seeking in the future (11).

Clinicians may struggle to distinguish autistic traits from symptoms of mental health conditions due to similarities in external presentation (8,12–14), which together with lack of clinician knowledge of autism (15) creates difficulties for autistic people accessing and receiving appropriate mental health support. This may lead to delayed or missed autism diagnosis, misdiagnosis, and ultimately ineffective treatment and support (16). Hence, there is a need to identify strategies to facilitate the detection of autism within mental health services.

Another potential barrier to obtaining high quality and appropriate mental health support is the lack of tailored care, as autistic people and their families recurrently report that services and treatment approaches are rarely adapted to their needs (15). Impediments to delivering appropriate support include lack of staff training and knowledge on how services could be adapted, and lack of evidence on best approaches (11,17–20). This scarcity of tailored approaches and lack of information about effective tailored care further highlights the need to identify strategies used to improve access, experiences of care and mental health outcomes for autistic people.

The aim of this review was to lay foundations for care that better meets the needs of autistic people by identifying and examining strategies that intend to improve mental health treatment and care for autistic adults. This was addressed through the following research questions:

1. What strategies, including bespoke and adapted mental health interventions, service adaptations and strategies to detect autism, have been developed to improve mental health care for autistic people?
2. What is the acceptability and feasibility of strategies to improve mental health care for autistic people?
3. What is the effectiveness of strategies to improve mental health care for autistic people?

## Methods

Our systematic review was conducted in accordance with the Preferred Reporting Items for Systematic Reviews and Meta-analyses (PRISMA) guidelines (21). A PRISMA checklist for this review is available in Table S1. The review was commissioned from the National Institute for Health and Care Research (NIHR) Mental Health Policy Research Unit (a research unit funded to deliver evidence to inform mental health policy in England) to meet an identified need for more evidence to guide policy in this area. We developed a review protocol that was prospectively registered on PROSPERO (CRD42022347690). The review protocol was developed through consultations with the review’s working group, which included lived experience researchers (i.e., people drawing on relevant lived experience to inform research), academics, clinicians and policy experts. All members of the working group had personal or professional expertise in autism and/or systematic review methodology. The working group met once a month from the inception of the review in July 2022 until its completion in March 2023. In the current paper, we have presented the findings regarding autistic adults and regarding mixed samples of adults and children and young people (CYP) populations in which only combined outcomes were reported. The findings for CYP will be reported in a separate paper.

### Search Strategy

Three electronic databases (MEDLINE (01/01/1994-DATE), PsycINFO (01/01/1994-DATE) and CINHAL (01/01/1994-DATE) and two pre-print servers (medRxiv and PsyArXiv) were searched using a combination of keyword and subject heading searches of terms for autism spectrum disorders and services/treatments and mental health problems. The search was restricted to studies published since 1994, to cover the Diagnostic and Statistical Manual of Mental Disorders fifth (DSM-IV) and fourth (DSM-V) edition periods. The reference lists of identified relevant systematic reviews were searched, and experts including academics and lived experience networks were contacted to identify relevant papers. The full search strategy can be found in Tables S2-S4.

### Inclusion and exclusion criteria

#### Population

For studies evaluating the effectiveness and/or acceptability/feasibility of treatment, we included adults (18+ years) and mixed samples of adults and CYP with an autism diagnosis, or who suspected they were autistic, or who were identified by clinicians as potentially autistic. Perspectives of carers and clinicians providing treatment to this population were also included. In all studies apart from those that explored detection of autism in mental health services, we excluded those with samples that included both autistic people and non-autistic people, unless data were reported separately for the autistic group.

#### Strategies

We included any bespoke or adapted mental health intervention (pharmacological, non-pharmacological or combinations) specifically for autistic people receiving mental health care from specialist mental health services and/or in primary care. We included studies describing and characterising bespoke or adapted mental health interventions for autistic people, or reporting on adaptations intended to improve access, experiences of care and mental health outcomes or strategies to identify autism in mental health services. Studies with any kind of comparison group (e.g., standard care, bespoke interventions or adapted approaches), or without a comparison group, were included.

#### Outcomes

We included any quantitative measure or qualitative account of feasibility (e.g., recruitment adherence and retention rates), service use (e.g., engagement), acceptability of care and experiences and satisfaction with care at end of treatment or follow-up for the second review question (RQ2). We also included any quantitative measure of mental health, detection of autism, quality of life, service use (e.g., inpatient admission, acute crisis care), and social outcomes (e.g., social functioning) at end of treatment or follow-up for the third review question (RQ3). Studies measuring physical health outcomes only were excluded.

#### Study types

All study designs and service descriptions reporting acceptability and feasibility outcomes were eligible for RQ2. Only empirical quantitative study designs, including service evaluations and clinical audits were eligible for RQ3. We excluded systematic or narrative reviews, small-N case studies, commentaries, book chapters, editorials, letters, conference abstracts and theses.

### Study selection

Two members of the review team (TS, PB) piloted the selection strategy. Title and abstract screening was conducted by members of the review team (AK, TS, KS, AG, TP, UF) with 10% of the search records reviewed in duplicates. The full text of eligible articles were then screened by members of the review team (TP, AG, AK, TS, DS, KS, SL, RC, JG, HB, UF), with 10% reviewed in duplicate. Conflicts were resolved by discussion and consultation with a third reviewer (SJ or VT) and with the working group. In instances where the setting or the intervention was unclear, study authors were contacted to determine eligibility. Study selection was carried out in Rayyan (22).

### Data extraction

Following study selection, members of the review team including lived experience researchers (TP, AG, AK, TS, DS, UF, SL, JG, HB, AT, MM, GB, RC) extracted the following: study design, aims, setting, sample size, participant characteristics (e.g., age, ethnicity, sex, diagnosis), outcome measures, strategies or adaptations (e.g., type and brief description) and relevant findings (feasibility, acceptability and effectiveness). The data extraction form was first piloted on 10% of the included studies (SL, RC) and revised accordingly based on feedback from the working group.

In response to the observation by lived experience researchers working on the study that a bias towards neurotypical people’s perspectives on autistic people’s difficulties was apparent in multiple studies, a decision was made in the working group to pilot a method of assessing this. Criteria for exploring the potential neurotypical bias in the included studies were developed by a lived experience researcher (RRO) based on a combination of existing literature and personal experience and piloted by members of the team (SL, TP). The five criteria used to assess the potential neurotypical bias were: 1) reported involvement from people with lived experience in the design, conduct, or write-up of the study; 2) for studies with qualitative elements, reported adjustments made to the data collection process (23); 3) for studies with quantitative elements, reported adjustments made to the data collection tools (24); 4) for studies with quantitative elements, reported adaptations or validity of relevant outcome measures for autistic people; 5) for studies with quantitative elements, perceived focus of the tested intervention/strategy on masking/changing autistic traits which might have not inherently impacted quality of life or caused distress (e.g., use of outcome measures relating to social skills or explicitly seeking to reduce autism symptoms) (25). Data for all criteria were extracted from the included studies by two researchers (AG, AK), with lived experience researchers (RJ, JP) as second assessors of the final criterion.

### Quality assessment and certainty of evidence

The Mixed Methods Appraisal Tool (MMAT) (26) was used to assess study quality. This is an established tool for evaluating quantitative, qualitative and mixed methods studies. Scores range from zero (low quality) to five (high quality). The Grading of Recommendations Assessment, Development and Evaluation (GRADE) system (27), adapted for narrative synthesis (28) was used to assess the strength of the evidence contributing to effectiveness outcomes. Two reviewers independently (TP, SL) assessed each outcome and addressed inconsistencies before reaching consensus.

### Data synthesis

A narrative synthesis was undertaken following Economic and Social Research Council (ESRC) guidelines (29). All identified intervention-level and service-level adaptations were grouped based on shared commonalities (e.g., environmental adjustments, adapted communication) to develop top-level and sub-level categories. Category development was informed by the input of lived experience researchers. Two meetings were held with lived experience researchers, in addition to the monthly working groups meetings, to share their views on the generated categories. Lived experience researchers were also given the opportunity to provide written feedback. To present extracted data, articles were categorised by the type of strategy (i.e., adaptations to mental health interventions, service improvements/adaptations or bespoke strategies) and study design (i.e., randomised controlled trials [RCTs], non-randomised controlled trials, surveys, before-and-after comparisons, and service evaluations). Extracted data related to the five criteria used to assess the potential neurotypical bias were synthesised descriptively. The review findings presented below have been refined and interpreted through discussions with and feedback from the working group.

## Results

The PRISMA flow diagram is shown in Figure 1. In total, 29 articles met the inclusion criteria. A list of included studies can be found in Table S5.

**Figure 1.**
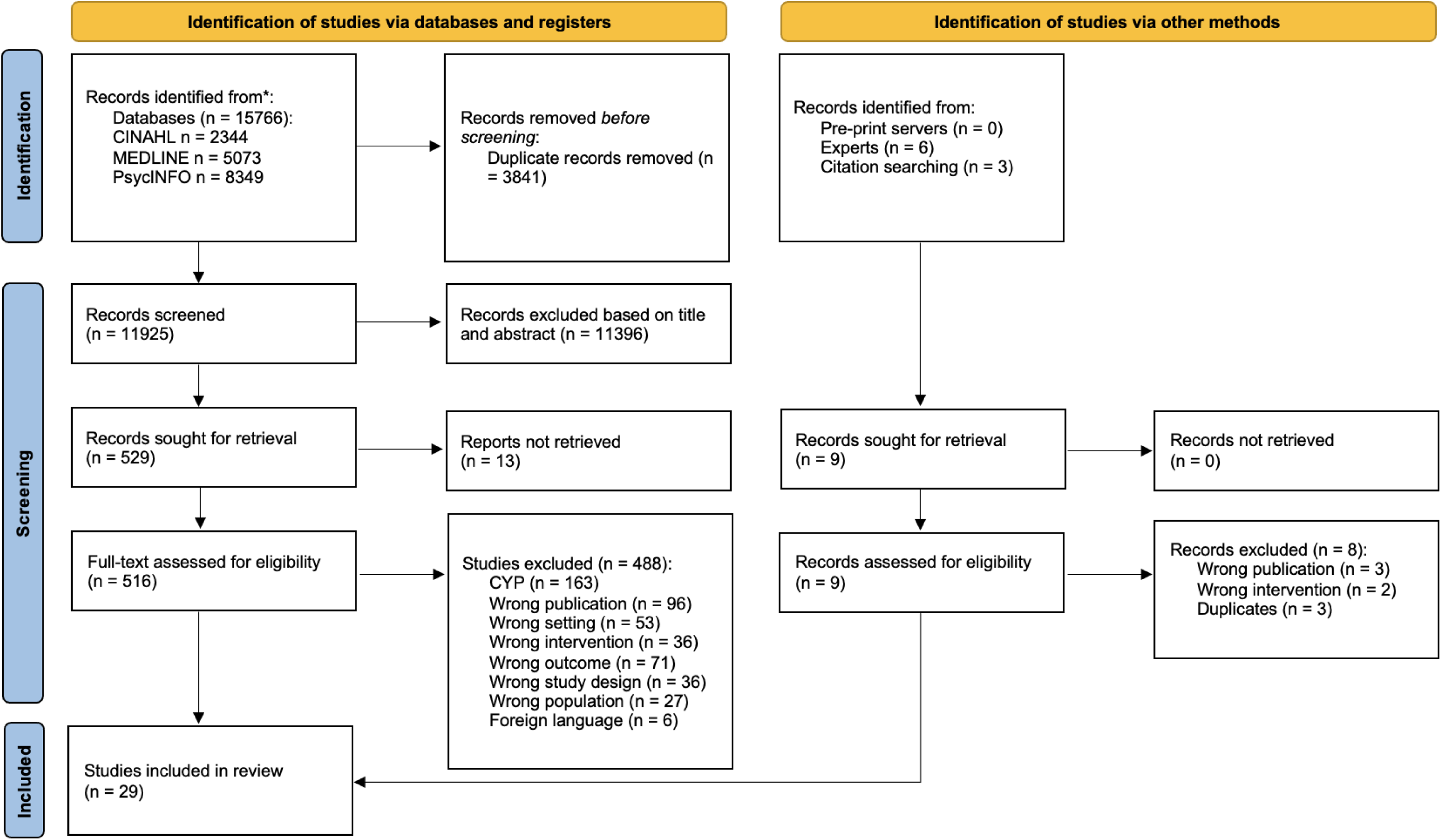
PRISMA flowchart

### Study design

Of the 29 articles, two were RCTs (30,31), three were pilot RCTs (32–34), three were non-randomised controlled trials (35–37), two were qualitative (38,39), one was retrospective analytical cross-sectional (40), four were surveys (41–44), four were service evaluations (45–48), and 10 were before-after comparison studies (49–58). Two of these articles were from the same trial (33,38). Study characteristics are described in detail in Tables 1 and S6.

**Table 1.**
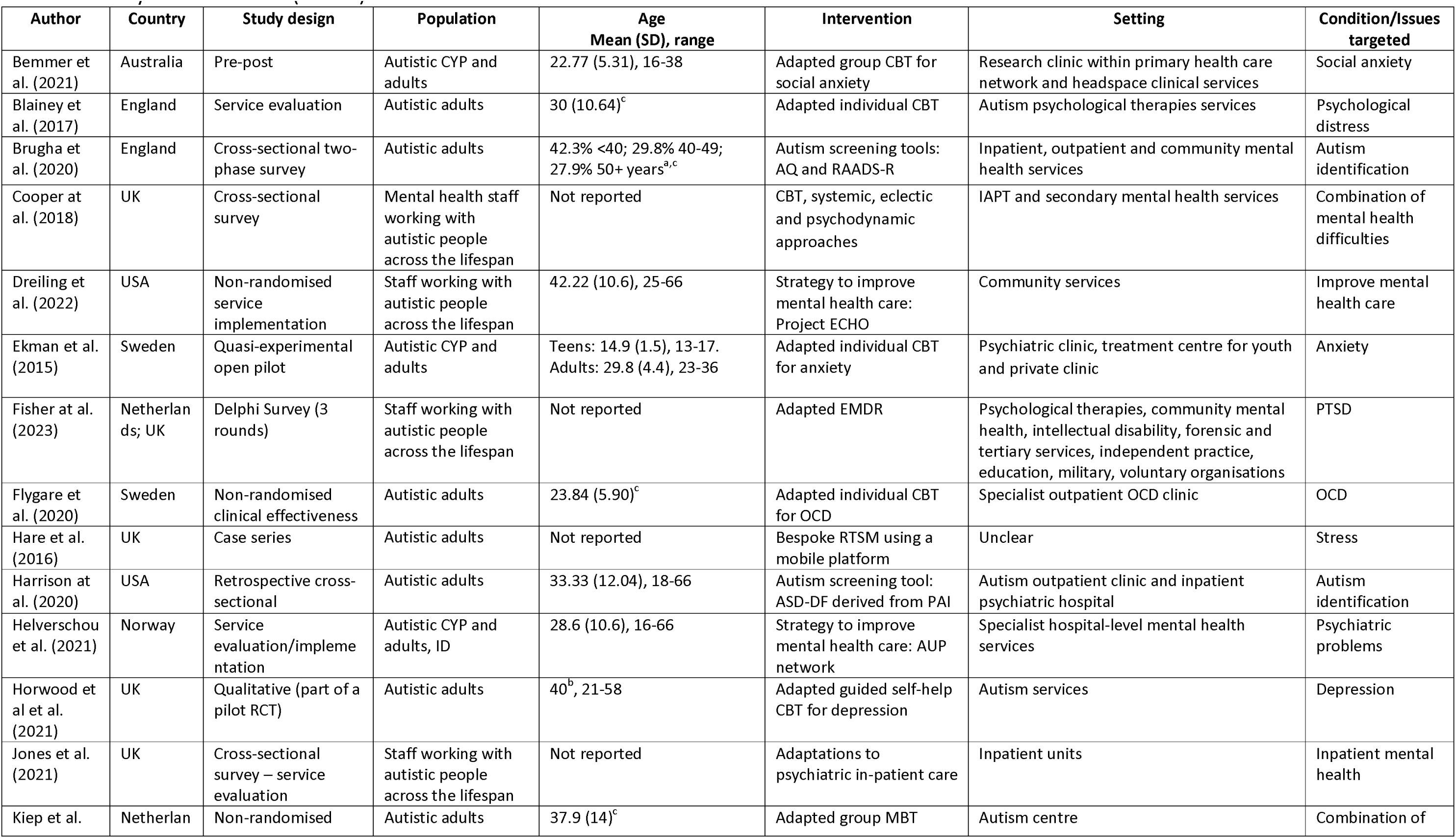
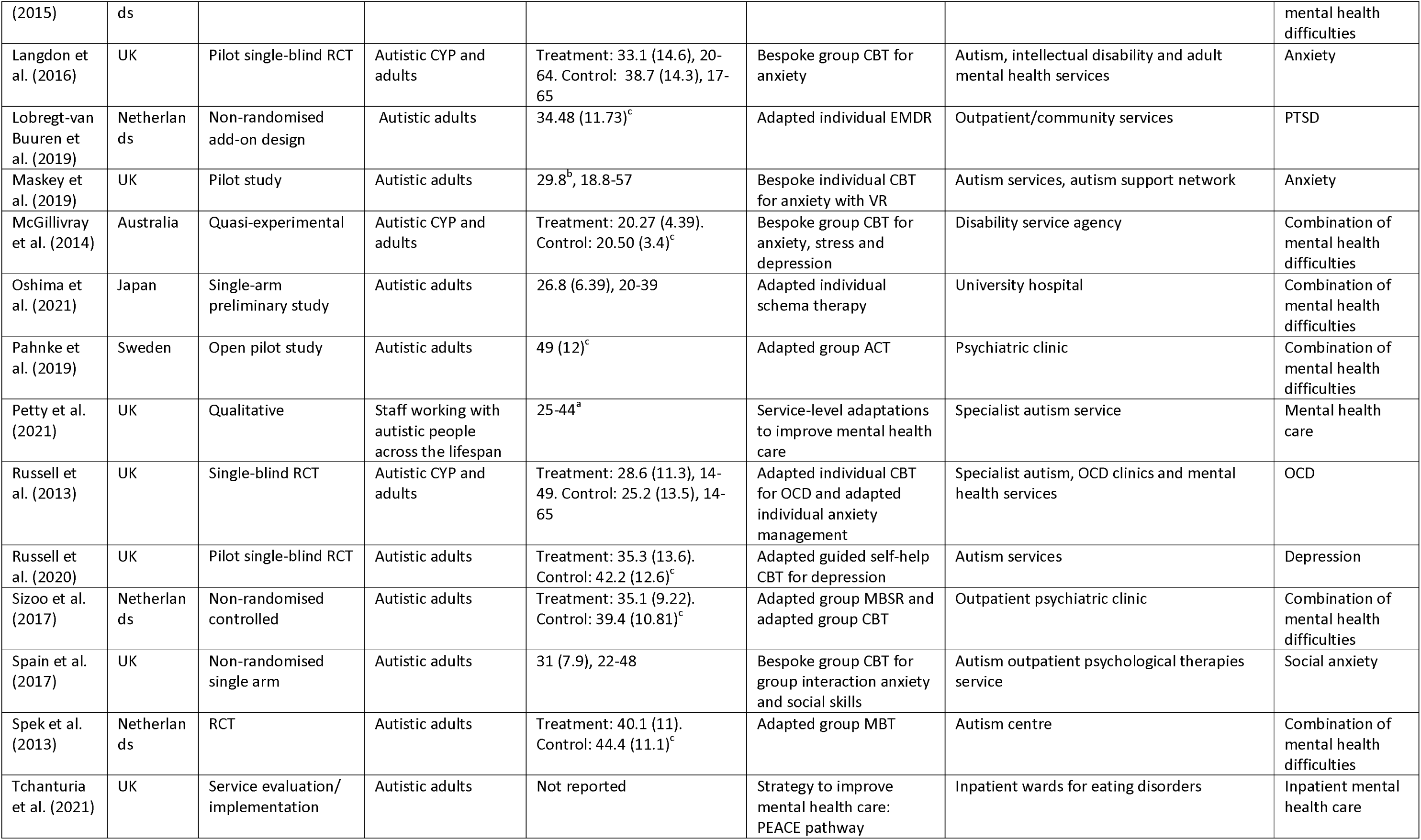
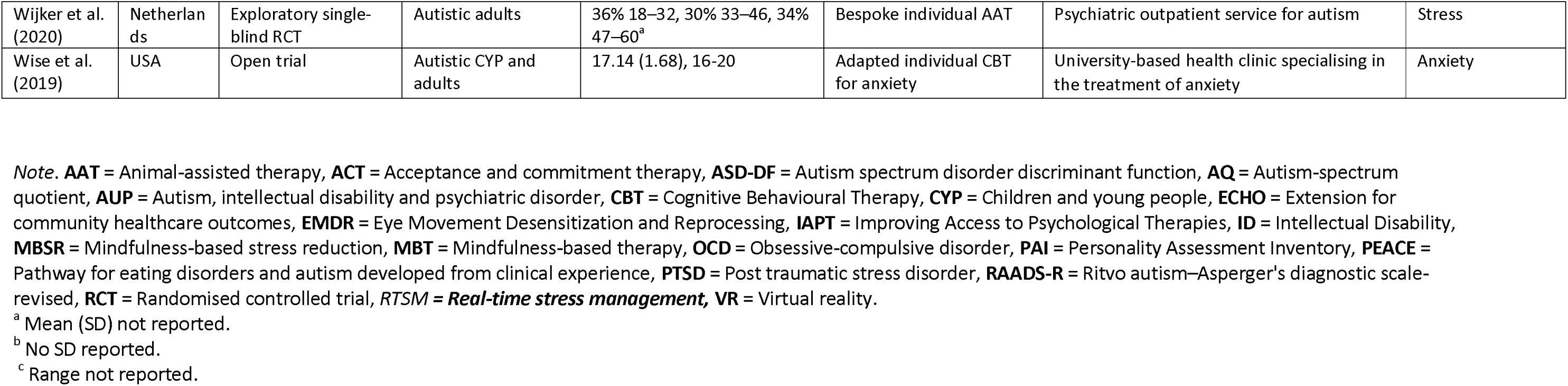
Study characteristics (*N* = 29)

### Quality assessment

According to appraisal using the MMAT (26), 18 studies (four randomised controlled, six non-randomised, three quantitative descriptive, three mixed, two qualitative) were of high quality (≥4 criteria met), six studies (all non-randomised) were of moderate quality (3 criteria met) and five studies (four non-randomised, one mixed) were of low quality (≤2 criteria met). All MMAT ratings can be seen in Table S7.

### Assessment of neurotypical bias

Seven out of 29 studies (24%) reported that autistic people were involved in conducting the study. None of the eight studies with a qualitative element reported any adjustments to the data collection process (e.g., allowing non-verbal/non-oral communication). Two out of 27 studies (7%) with a quantitative element reported making some adjustments to the data collection tools (e.g., adapting Likert scales for greater precision, using straightforward language). Ten out of 27 studies (37%) with a quantitative element reported on the psychometric properties or adaptations of the relevant outcome measures to make them more appropriate for autistic individuals. Six out of the 10 studies (60%) used at least one adapted or validated outcome measure relevant to the review, and the remaining four studies (40%) stated that the relevant outcome measures had not been validated or adapted specifically for autistic people. For five of the 21 studies (24%) with a quantitative element that measured outcomes in autistic mental health service users, the intervention/strategy were perceived to involve some focus on masking people’s autistic traits. However, thirteen of the 21 studies (62%) were not perceived to have any evidence to suggest such a focus, and this was unclear for three of the twenty-one studies (14%). See Table S8 for all extracted data related to the neurotypical bias assessment.

### Sample characteristics

Sample sizes at baseline were small across most studies, ranging from 7 to 1487 (median 103, n = 28 studies). Twenty-four studies included participants who were service users. All participants of these studies had a diagnosis of autism except for two studies relating to the detection of autism, which included people who had not been diagnosed as autistic at the time study data were obtained (40,41). Only one study included participants with intellectual disability (ID) (47).

Twenty studies reported on co-occurring mental health conditions at baseline: obsessive compulsive disorder (OCD) (30,51), depression (33,38), anxiety (32,54), post-traumatic stress disorder (PTSD) (36), stress (34), eating disorders (48) and a combination of mental health difficulties (31,35,37,40,41,45,50,53,55,56,58). Sixteen papers reported on adults (31,33,34,36–38,40,45,48,51–57) seven papers reported on both CYP and adults (30,32,35,47,49,50,58). Five papers reported on staff perspectives of how to adapt and deliver better care for autistic people across the lifespan (39,42–44,46). Sample characteristics are described in detail in Table S6.

### Types of strategies used to improve mental health care in autism

#### Intervention-level and service-level adaptations

Studies tended to use several adaptations; hence most studies were found to be relevant to multiple categories. The following six top-level adaptation categories were identified: communication accommodations (n = 16), intervention content (n = 13), intervention structure (n = 9), increase knowledge and detection of autism (n = 8), accommodate individual differences (n = 8) and environmental adjustments (n = 4). Tables 2 and S9 describe the adaptations in each study. Most studies reported a general rationale for adaptations (e.g., to address barriers and needs of autistic people). Individual adaptations often lacked a comprehensive description and rationale.

**Table 2.** All service-level and intervention-level adaptations (simplified version) (*N* = 23)

| Top-level categories | Sub-categories | Summary | *N* studies |
| --- | --- | --- | --- |
| Increase knowledge and detection of autism | Clinician training and skills | Training to administer measures, tailor treatment to individual needs and increase self-efficacy, knowledge of autism and skills. Use of skills such as normalising experiences and prioritising therapeutic relationship. | 5 |
|  | Introduction of screening tools for the detection of autism | Use of assessments such as the Autism Spectrum Disorder-Discriminant Function (ASD-DF), the Autism-spectrum Quotient (AQ) and the Ritvo autism-Asperger's diagnostic scale-revised (RAADS-R). | 3 |
| Environmental adjustments | Provide environmental and practical adjustments | Provide adjustments to ensure consistency such as offering sessions at the same time and place and minimise sensory distractions such as a low stimulus area, adjustments to noise, decor, odor, lighting and meals. | 4 |
|  | Normalise the use of sensory resources and stimming | Provide sensory materials such as ear defenders, weighted blankets, stress ball, relaxing music and sensory box. Encourage use of stimming behaviour. | 4 |
| Communication accommodations | Plan in advance | Share a plan in advance, ensure the client is prepared, find out about the client in advance, use of appointment reminders, pay extra attention to planning and discuss issues. | 5 |
|  | Clear communication | Provide clear instructions and guidance, repetition, be more directive, monitor, adapt and slow the pace of communication. | 5 |
|  | Use of simple and preferred language | Avoid use of metaphors, abstract language, awareness of the language, use of plain and preferred language. | 10 |
|  | Use of simple, written material and visual aids | Use of written information and external cues such as use of a whiteboard, color-coded worksheets, timers, agendas and calendars. Use of visual aids such as drawings, pictures, videos and leaflets. | 11 |
|  | Provide communication support | Host wellbeing groups and use of communication passports and social stories. | 2 |
| Accommodate individual differences | Evaluate individual needs and preferences | Evaluate preferences, sensitivities, needs, likes and dislikes, coping strategies and daily habits. | 5 |
|  | Encourage individual's hobbies and interests | Include and ask about the individual's special interests and hobbies in therapy. | 4 |
|  | Tailor practice to individual needs and preferences | Tailor care plans and practice to individual differences such as incorporating approaches targeted at neurodevelopmental comorbidities, being flexible with the treatment manual and the session timings and ensuring that resources are appropriate for the person's gender. | 6 |
| Intervention structure adaptations | Format of intervention | Reduce or increase the number and duration of sessions and exercises, additional support by therapists. | 6 |
|  | Family/caregiver/other involvement | Involve important people such as family members, partners, teachers throughout therapy. | 6 |
| Intervention content adaptations | Simplified and structured content | Remove or simplify psychoeducation and cognitive elements and ensure that there is structure. | 6 |
|  | Taking it slow | Taking a slow/progressive approach to treatment, regular breaks. | 2 |
|  | Consider the role of autism | Consider the role of autism, develop an understanding of autism such as its characteristics and impact on daily life. | 2 |
|  | Integration of emotion-focused strategies | Provide psychoeducation on emotions, arousal and feeling physiologically overwhelmed and exercises to access emotions. | 6 |
|  | Integration of cognitive-behavioural approaches | Provide cognitive and behavioural strategies including building a positive self-image, coping strategies, and making links between behaviour, thoughts and feelings. | 6 |
|  | Integration of social skills training | Integration of social skills training such as entering and maintaining conversations and managing disagreements | 3 |

Sixteen articles described studies of mental health interventions that had been adapted to make them more appropriate for autistic people. These included adaptations of Cognitive Behavioural Therapy (CBT) for anxiety (49,50,58), CBT for OCD (30,51), mindfulness-based therapy for autism spectrum disorders (MBT-AS) (31,53), Eye Movement Desensitisation and Reprocessing (EMDR) (36,43), guided self-help CBT for depression (33,38), acceptance and commitment therapy (ACT) (56), Schema therapy (55), mindfulness-based stress reduction (MBSR) (37), CBT for anxiety and depression (37), CBT aimed to reduce general psychological distress (42,45). No trials directly compared adapted and non-adapted mental health interventions; hence no conclusions could be drawn as to whether specific adaptations resulted in better outcomes.

Seven articles described studies investigating service adaptations, largely related to autism-specific training of staff and environmental adjustments. These studies examined clinical pathways (48), models (46), networks (47) and general adaptations (39,44) to improve quality of mental health care for autistic people, and initiatives to improve the detection of autism (40,41).

#### Bespoke mental health interventions

Six studies examined bespoke mental health interventions designed for autistic people. These were individual Real-Time Stress Management (RTSM) using a mobile platform (52), CBT for anxiety (32), CBT for social anxiety (57), CBT for anxiety in combination with Virtual Reality (54), CBT for anxiety, stress and depression (35) and animal assisted therapy (AAT) (34).

### Evaluation of strategies used to improve mental health care in autism

#### Certainty of evidence for effectiveness of strategies

Results from twenty-three studies contributed to the GRADE assessment. The certainty of evidence for the effectiveness of strategies (n = 19), as rated using the GRADE system (27), ranged from very low to moderate (Table S10). No strategies were assessed to have high-certainty evidence for effectiveness and only three out of 19 (16%) strategies were deemed to have moderate-certainty effectiveness evidence, i.e., adapted individual CBT interventions for anxiety (50,58) and for OCD (30,51), and strategies for detection of autism (40,41). Certainty of evidence for effectiveness was generally very low or low for the remaining included strategies (53% and 32%, respectively).

#### Adaptations to mental health interventions

Overall, most adapted mental health interventions were evaluated as feasible and acceptable, except for one study of CBT for OCD, which showed limited feasibility (51). Individual adaptations were largely viewed positively (38,50), and clinicians reported frequently used adaptations to CBT (42) and EMDR (43). Evidence on effectiveness was inconclusive as RCTs were not sufficiently powered and most were before-and-after comparison studies. The main findings of adapted mental health interventions can be found in Tables 3 and 4, and detailed results of individual studies are available in Table S11.

**Table 3.** Main findings of adaptations to individual mental health interventions

| Study design<br>(N studies) | Study characteristics (N studies) | Participant characteristics<br>(N studies) | Strategy vs Comparison<br>(if relevant)<br>(N studies) | Top-level adaptations categories and sub-categories<br>(N studies) | Main Findings<sup>b</sup><br>& Certainty of Evidence |
| --- | --- | --- | --- | --- | --- |
| **Adapted individual CBT for anxiety** |  |  |  |  |  |
| **Before-and-after comparison (2)**<br>(Ekman et al., 2015; Wise et al., 2019) | Sample size: 7-18 (2).<br>Country: USA (1); Sweden (1)<br>Setting: Psychiatric clinic, treatment centre for youth and private clinic (1); University-based health clinic specialising in the treatment of anxiety (1) | Diagnoses: clinical ASC diagnosis (2); anxiety (2).<br>Demographics: Adults (1); CYP and adults (1); 39-57% male (2); 14% non-white (1).<br>Type of participants: Service users (2). | Individual adapted CBT for anxiety (2) | **Communication accommodations (2)**<br>- Use of simple, written material and visual aids (2)<br>**Accommodate individual differences (1)**<br>- Encourage individual's hobbies and interests (1)<br>**Intervention structure (1)**<br>- Family/caregiver/other involvement (1)<br>**Intervention content (1)**<br>- Integration of emotion-focused strategies (1)<br>Integration of cognitive-behavioural approaches (1);<br>Integration of social skills training (1) | **Acceptability:** use of visualisation was perceived as helpful by participants (1).<br>**Feasibility:** low drop-out rate (1).<br>**Effectiveness:** Significant pre-post improvements in mental health outcomes (2); social outcomes (1). No significant pre-post change in mental health outcomes (1)<br>**- Certainty of evidence (2):** low. |
| **Adapted individual CBT for OCD** |  |  |  |  |  |
| **RCT (1); Before-and-after comparison (1)**<br>(Russell et al., 2013; Flygare et al., 2017) | Sample size: 19-46 (2).<br>Country: UK (1); Sweden (1)<br>Setting: Specialist outpatient OCD clinic (1); Specialist autism, OCD clinics and mental health services (1). | Diagnoses: clinical ASC diagnosis (2); OCD (2).<br>Demographics: Adults (1); CYP and adults (1); 42-70% male (2).<br>Type of participants: Service users (2). | Individual CBT for OCD vs Anxiety management (1); Individual CBT for OCD (1) | **Communication accommodations (2)**<br>- Clear communication (1); Use of simple and preferred language (2); Use of simple, written material and visual aids (2)<br>**Accommodate individual differences (1)**<br>- Evaluate individual needs and preferences (1); Encourage individual's hobbies and interests (1)<br>**Intervention structure (1)**<br>- Family/caregiver/other involvement (1)<br>**Intervention content (2)**<br>- Integration of emotion-focused strategies (2); Simplified and structured content (1); Integration of cognitive-behavioural approaches (1) | **Acceptability:** there were no significant differences in treatment satisfaction between adapted CBT for OCD and anxiety management (moderate treatment satisfaction was reported for both). Significantly more patients in the anxiety management group requested crossover to the CBT group than vice versa (1), good homework compliance (1)<br>**Feasibility:** high attendance rate (1), low drop-out rate (2), low attendance rate (1)<br>**Effectiveness:** The adapted CBT for OCD was not superior to anxiety management group on any of the primary and secondary mental health and social outcomes (1). Significant pre-post improvements in mental health outcomes (1). No significant pre-post change in social outcomes (1), quality of life (1).<br>**- Certainty of evidence (2):** moderate. |

| Adapted EMDR |  |  |  |  |  |
| --- | --- | --- | --- | --- | --- |
| **Non-randomised controlled trial (1); Survey (1)**<br>(Lobregt-van Buuren et al. 2019; Fisher et al., 2023) | Sample size: 27 (1), 103 (1).<br>Country: Netherlands & UK (1); Netherlands (1).<br>Setting: Mixed mental health care setting (1); Outpatient/community services (1). | Diagnoses: clinical ASC diagnosis (1); PTSD (1).<br>Demographics: Adults (1); 62% male (1).<br>Type of participants: Staff working with autistic individuals across the lifespan (1), Service users (1). | EMDR + TAU vs TAU (1); EMDR (1) | **Communication accommodations (2)**<br>- Use of simple and preferred language (2); Plan in advance (1); Clear communication (1); Use of simple, written material and visual aids (1)<br>**Increase knowledge and detection of autism (1)**<br>- Clinician training and skills (1)<br>**Environmental adjustments (1)**<br>- Provide environmental and practical adjustments (1); Normalise the use of sensory resources and stimming (1)<br>**Accommodate individual differences (1)**<br>- Evaluate individual needs and preferences (1); Encourage individual's hobbies and interests (1); Tailor practice to individual needs and preferences (1)<br>**Intervention structure (1)**<br>- Family/caregiver/other involvement (1)<br>**Intervention content (1)**<br>- Taking it slow (1); Consider the role of autism (1); Integration of emotion-focused strategies (1); Integration of cognitive-behavioural approaches (1) | **Acceptability:** EMDR sessions were perceived as stressful, but a high majority would choose to do it again (1).<br>**Feasibility:** Most adaptations were frequently used by staff across studies (1); low drop-out rate (1).<br>**Effectiveness:** Significant pre-post improvements in mental health outcomes (1)<br>**- Certainty of evidence (1):** low. |
| Adapted Schema therapy |  |  |  |  |  |
| **Before-and-after comparison (1)**<br>(Oshima et al., 2021) | Sample size: 12 (1).<br>Country: Japan (1).<br>Setting: University hospital (1). | Diagnoses: clinical ASC diagnosis (1).<br>Demographics: Adults (1); 50% male (1).<br>Type of participants: Service users (1). | Schema therapy (1) | **Intervention content (1)**<br>- Consider the role of autism (1) | **Feasibility:** low drop-out rate (1).<br>**Effectiveness:** Significant pre-post improvements in social outcomes. No significant pre-post change in mental health outcomes and quality of life (1).<br>**- Certainty of evidence (1):** very low. |
| Adapted individual CBT for general psychological distress |  |  |  |  |  |
| **Before-and-after comparison (1); Survey (1)**<br>(Blainey et | Sample size: 50-81 (2).<br>Country: UK (2).<br>Setting: Autism psychological therapies services (1); IAPT and secondary mental health | Diagnoses: clinical ASC diagnosis (1).<br>Demographics: Adults (1); 74.1% male (1); 40.7% non-White British | CBT (2) | **Communication accommodations (2)**<br>- Use of simple, written material and visual aids (2); Clear communication (1); Use of simple and preferred language (1)<br>**Accommodate individual differences (2)**<br>- Tailor practice to individual needs and preferences (1); | **Acceptability:** CBT was favoured by therapists compared to systemic, eclectic and psychodynamic approaches (1).<br>**Feasibility:** high attendance rate (1); most adaptations were frequently used by staff across studies (1). Adaptations that were used less |
| al., 2017; Cooper et al., 2018) | services (1). | (1).<br>Type of participants: Staff working with autistic individuals across the lifespan (1), Service users (1). |  | Encourage individual's hobbies and interests (1)<br>**Intervention structure (2)**<br>- Format of intervention (2); Family/caregiver/other involvement (2)<br>**Intervention content (2)**<br>- Integration of social skills training (1); Simplified and structured content (1); Integration of emotion-focused strategies (1); Integration of cognitive-behavioural approaches (1) | consistently included: shorter or longer sessions, avoidance of metaphors in therapy, involving a family member or partner in sessions, use of cognitive strategies.<br>**Effectiveness:** Significant pre-post improvements in mental health outcomes (1).<br>- **Certainty of evidence (1):** very low. |
| **Adapted individual guided self-help CBT for depression** |  |  |  |  |  |
| **RCT (1); Qualitative (1)**<br>(Horwood et al., 2021; Russell et al., 2020)<sup>a</sup> | Sample size: 26-70 (2).<br>Country: UK (2).<br>Setting: Autism services (1). | Diagnoses: clinical ASC diagnosis (2); depression (2).<br>Demographics: Adults (2), 69-81% male (2); 0-6% non-white (2)<br>Type of participants: Service users only (1); service users and staff (1). | Guided self-help for depression vs TAU (2) | **Communication accommodations (1)**<br>- Plan in advance; Use of simple, written material and visual aids (1)<br>**Accommodate individual differences (1)**<br>- Evaluate individual needs and preferences (1); Tailor practice to individual needs and preferences (1)<br>**Intervention structure (1)**<br>- Format of intervention (1)<br>**Intervention content (1)**<br>- Simplified and structured content (1); Integration of cognitive-behavioural approaches (1) | **Acceptability:** Guided self-help CBT was viewed positively by participants (1).<br>**Feasibility:** High attendance rate (1); low drop-out rate (1). |

**Note.** ASC = Autism spectrum condition, CBT = Cognitive Behavioural Therapy, EMDR = Eye Movement Desensitization and Reprocessing, IAPT = Improving Access to Psychological Therapies, ID = Intellectual Disability, OCD = Obsessive-compulsive disorder, PTSD = Post traumatic stress disorder.
<sup>a</sup> Two articles were from the same pilot randomised controlled trial, one of which was a qualitative study examining the acceptability of guided self-help CBT for depression.
<sup>b</sup> Studies measuring effect in multiple mental health outcomes are counted to reflect the findings for each outcome comparison conducted.

**Table 4.** Main findings of adaptations to group mental health interventions

| Study design<br>(N studies) | Study characteristics<br>(N studies) | Participant characteristics<br>(N studies) | Strategy vs Comparison<br>(if relevant)<br>(N studies) | Top-level adaptations categories and sub-categories<br>(N studies) | Main Findings<sup>a</sup><br>& Certainty of Evidence |
| --- | --- | --- | --- | --- | --- |
| **Adapted group MBT** |  |  |  |  |  |
| **RCT (1); Non-randomised controlled trial (1); Before-and-after comparison (1)** (Sizoo et al., 2017; Spek et al., 2013; Kiep et al., 2015) | Sample size: 42-59 (3).<br>Country: Netherlands (3).<br>Setting: Autism services (2); Outpatient psychiatric clinic (1) | Diagnoses: clinical ASC diagnosis (3); depression and/or anxiety (3).<br>Demographics: Adults (3), 59-70% male (3).<br>Type of participants: Service users (3). | Group MBT vs wait-list (1); group MBT (1); Group CBT for anxiety and depression vs Group MBSR (1) | **Communication accommodations (3)**<br>- Plan in advance (2); Use of simple and preferred language (3); Clear communication (1)<br>**Intervention structure (2)**<br>- Format of intervention (2)<br>**Intervention content (3)**<br>- Simplified and structured content (2); Taking it slow (1) | **Feasibility:** High attendance rate (2); low drop-out rate (2).<br>**Effectiveness:** Compared to wait-list, participants receiving MBT significantly improved on all mental health outcomes. (1). Participants in both adapted CBT and adapted MBSR significantly improved in all mental health outcomes and there was no significant difference between treatment groups (1). Significant pre-post improvements in mental health outcomes (1).<br>- **Certainty of evidence (3):** moderate. |
| **Adapted group CBT for anxiety and depression** |  |  |  |  |  |
| **Non-randomised controlled trial (1)** (Sizoo et al., 2017) | Sample size: 59.<br>Country: Netherlands<br>Setting: Outpatient psychiatric clinic | Diagnoses: clinical ASC diagnosis; depression and/or anxiety.<br>Demographics: Adults, 59-70% male.<br>Type of participants: Service users. | Group CBT for anxiety and depression vs Group MBSR | **Communication accommodations (1)**<br>- Plan in advance; Clear communication (1); Use of simple and preferred language (1)<br>**Intervention content (1)**<br>- Taking it slow (1) | **Effectiveness:** Participants in both adapted CBT and adapted MBSR significantly improved in all mental health outcomes and there was no significant difference between treatment groups (1).<br>- **Certainty of evidence (1):** very low. |
| **Adapted group CBT for social anxiety** |  |  |  |  |  |
| **Before-and-after comparison (1)** (Bemmer et al., 2021) | Sample size: 84.<br>Country: Australia<br>Setting: Research clinic within primary health care network and headspace | Diagnoses: clinical ASC diagnosis.<br>Demographics: Autistic CYP and adults, 60% male.<br>Type of participants: | Group CBT for social anxiety | **Intervention content (1)**<br>- Simplified and structured content; Integration of cognitive-behavioural approaches (1); Integration of social skills training (1) | **Acceptability:** High enjoyment, positive engagement, though some reported difficulties with a group member who was perceived as disruptive (1).<br>**Feasibility:** Low drop-out rate (1).<br>**Effectiveness:** Significant pre-post improvements in mental |
|  | clinical services. | Service users. |  |  | health outcomes (1).<br>- **Certainty of evidence (1)**: very low. |
| **Adapted group ACT** |  |  |  |  |  |
| **Before-and-after comparison (1)** (Pahnke et al., 2019) | Sample size: 10.<br>Country: Sweden.<br>Setting: Psychiatric clinic. | Diagnoses: clinical ASC diagnosis.<br>Demographics: Autistic adults.<br>Type of participants: Service users. | Group ACT | **Communication accommodations (1)**<br>- Clear communication (1); Use of simple and preferred language (1); Use of simple, written material and visual aids (1)<br>**Intervention structure (1)**<br>- Format of intervention (1)<br>**Intervention content (1)**<br>- Integration of emotion-focused strategies (1) | **Acceptability**: High treatment credibility (1).<br>**Feasibility**: high attendance rate, low drop-out rate, good homework compliance (1).<br>**Effectiveness**: Significant pre-post improvements in mental health outcomes and social outcomes. No significant pre-post change in mental health outcomes, quality of life (1).<br>- **Certainty of evidence (1)**: very low. |

*Note.* **ACT** = Acceptance and commitment therapy; **ASC** = Autism spectrum condition, **CBT** = Cognitive Behavioural Therapy; **MBT** = Mindfulness-based therapy; **MBSR** = Mindfulness-based stress reduction.
<sup>a</sup>Studies measuring effect in multiple mental health outcomes are counted to reflect the findings for each outcome comparison conducted.

#### Randomised controlled trials (RCTs) and pilot RCTs

Three trials, including one pilot, evaluated adapted mental health interventions. One RCT found that compared to waiting list, adapted group MBT-AS led to significant improvements in all mental health outcomes including depression, anxiety, positive affect and rumination (31). The intervention also had a low drop-out rate and a high attendance rate. Another RCT found no significant differences in treatment satisfaction and in primary (clinician-assessed OCD symptoms) and secondary (self-report OCD symptoms, anxiety and depression) mental health post-treatment outcomes between adapted individual CBT for OCD and adapted individual anxiety management, apart from parent-report OCD symptoms that significantly reduced only in the anxiety management group over time (30). Attendance rates were higher in the CBT arm; however, drop-out rates were similar in both groups. Neither RCT was sufficiently powered to demonstrate an effect. A pilot RCT comparing adapted individual guided self-help CBT for depression and TAU found that the former had a lower drop-out rate, whilst also achieving an acceptable attendance rate (33). A subsequent qualitative study (38) using a subset of the sample demonstrated that the intervention was also viewed positively by most participants. However, there were differing views about the pacing of the sessions and the use of pre-defined visual tools.

#### Non-randomised controlled trials

Two non-randomised controlled trials evaluated adapted mental health interventions. A study comparing the effectiveness of adapted group MBSR and adapted group CBT for anxiety and depression reported no significant differences in mental health outcomes of anxiety, depression, positive and negative general mood and rumination at post-treatment (37). Both MBSR and CBT were adapted in the same way (see Tables 4 and S9). A study with a non-randomised add-on design reported significant improvements in PTSD symptoms and psychological distress following EMDR + TAU compared to TAU only (36). The effect remained stable at 6-8 weeks follow-up. The study showed a low drop-out rate and, whilst all participants found EMDR sessions stressful, most indicated that they would choose the therapy again.

#### Surveys investigating perspectives of staff

Two surveys examined the perspectives of staff of adapted interventions to improve mental health care for autistic people. One survey found that most adaptations to CBT targeting psychological distress (e.g., use of plain English, structured and concrete approach) were highly endorsed by therapists, while others appeared to be used less consistently (e.g., shorter or longer sessions, avoidance of metaphors) (42). Findings from a Delphi survey reported an array of adaptations identified by therapists as always, often or sometimes incorporated in EMDR (43). These included environmental adjustments, normalising experiences, communicating clearly, being flexible with the treatment manual, taking a slow approach and considering the role of autism within conceptualisation.

#### Before-and-after comparison studies

Eight before-and-after comparison studies examined adapted mental health interventions. Significant improvements in outcomes over time were reported in all studies (Tables 3 and 4). However, causality cannot be inferred as there were no comparison groups, thus these will not be reported in detail.

A pre-post study of group CBT for social anxiety reported a low drop-out rate and a high participant enjoyment (49). Another study found that group MBT-AS had acceptable attendance and drop-out rates (53). Additionally, high levels of attendance, retention, homework compliance and treatment credibility were reported regarding group ACT (56) One study reported a high attendance rate to adapted CBT for anxiety (45). A study of CBT for anxiety (58) and a study of schema therapy (55) reported low drop-out rates. Most participants found the adaptations (i.e., use of visualisation) of CBT for anxiety to be helpful (50). Drop-out rate of CBT for OCD was low and homework compliance was adequate to good, however, attendance rate was low (51).

### Mental health service adaptations

Overall, strategies to improve clinicians’ knowledge of autism and provide environmental adjustments in services were evaluated as acceptable and feasible (46–48). Several service adaptations were reported as frequently implemented in services by staff (39,44). Additionally, self-report tools were found to discriminate autistic and non-autistic people (40,41). Table 5 presents findings of evaluations of strategies at service level intended to improve care.

**Table 5.**
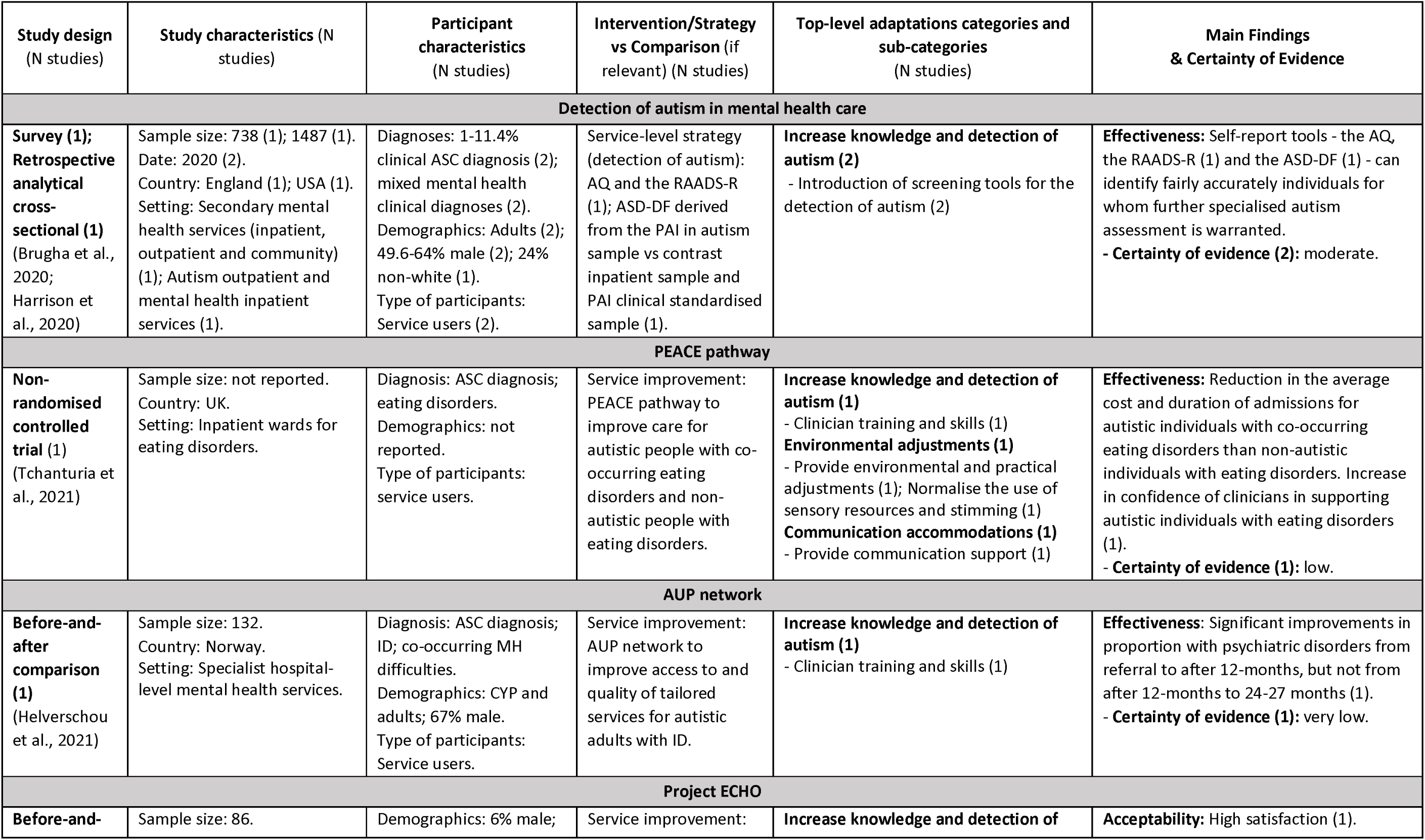
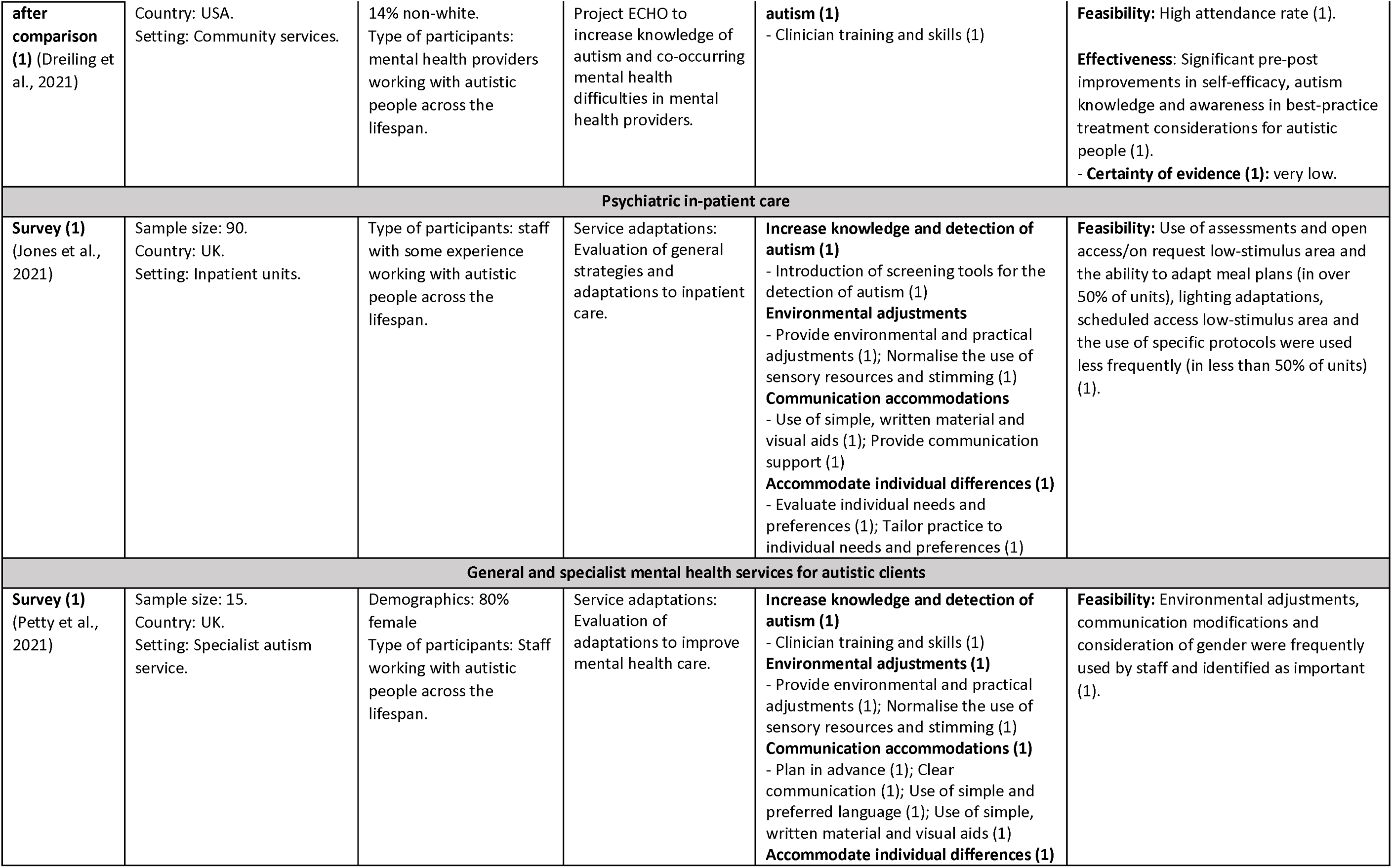
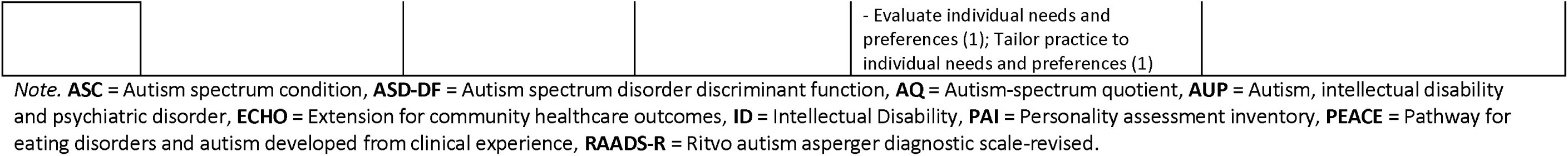
Main findings of mental health service adaptations

#### Service evaluations

Three studies examined clinical pathways, models and networks to improve mental health care for autistic people. The Pathway for Eating Disorders and Autism developed from Clinical Experience (PEACE) aimed to introduce autism specific-training, create an autism-friendly ward and support sensory difficulties and communication (48). The pathway resulted in more reductions in the cost and average duration of hospital admissions in autistic individuals with eating disorders than non-autistic individuals with eating disorders. Evaluation of the PEACE pathway suggested that confidence of clinicians in supporting autistic people with co-occurring eating disorder increased following its implementation.

Project Extension for Community Healthcare Outcomes (ECHO) utilised a tele-mentoring platform to connect primary care providers to increase knowledge of autism and co-occurring mental health difficulties and to appropriately adapt treatments (46). Project ECHO was attended by most mental health providers, and increased knowledge, self-efficacy, and awareness in best-practice treatment considerations for autistic individuals was reported post-ECHO sessions. The project was also viewed positively and was rated highly on satisfaction.

Another study reported significant improvements in the proportion of psychiatric disorders from referral to after 12 months, which were sustained from 12 months to 24-27 months post-implementation of the Autism Intellectual disability and psychiatric disorder (AUP) network (47). The AUP network aimed to improve access and quality of tailored services for autistic adults with ID and increase knowledge of clinicians of how mental health difficulties present in autistic people.

#### Perspectives of staff

Two studies identified service adaptations. One survey reported that the most frequent adaptations within inpatient units involved the use of assessments and open access/on request low-stimulus area, and the ability to adapt meal plans (over 50% of units), whilst lighting adaptations, scheduled access low-stimulus area and the use of specific protocols were used less frequently (less than 50% of units) (44). Another qualitative study also reported a range of adaptations identified by staff, including environmental adjustments, communication modifications and consideration of gender, the majority of which were frequently used and perceived as important (39).

#### Detection of autism

Two studies involved application in routine settings of autism screening instrument with the aim of improving detection within mental health services. One survey found that the autism-spectrum quotient (AQ) and the Ritvo autism–Asperger’s diagnostic scale-revised (RAADS-R) can identify fairly accurately autistic individuals from inpatient, outpatient and community mental health services (41). A retrospective study reported that an autism spectrum disorder discriminant function (ASD-DF) derived from the Personality Assessment Inventory (PAI) can discriminate between a sample of autistic participants from a contrasting inpatient sample (40).

### Bespoke strategies to improve mental health care for autistic people

Most bespoke mental health interventions were evaluated as feasible and acceptable, apart from one study of RTSM (52), which demonstrated limited feasibility and acceptability. Evidence on effectiveness was inconclusive. Table 6 presents findings of evaluations of bespoke mental health interventions designed for autistic people.

**Table 6.** Main findings of bespoke strategies to improve mental health care for autistic people

| Study design (N studies) | Study characteristics (N studies) | Participant characteristics (N studies) | Intervention/Strategy vs Comparison (if relevant) (N studies) | Main Findings & Certainty of Evidence |
| --- | --- | --- | --- | --- |
| **Bespoke individual CBT for anxiety with VR** |  |  |  |  |
| **Before-and-after comparison (1)**<br>(Maskey et al., 2019) | Sample size: 8.<br>Country: UK.<br>Setting: Autism services, autism support network. | Diagnosis: ASC diagnosis; anxiety and/or depression.<br>Demographics: Adults; 5% male.<br>Type of participants: service users. | Bespoke CBT for anxiety in combination with virtual reality. | **Feasibility:** High attendance rate, low drop-out rate (1).<br>**Effectiveness:** 5 of the 8 participants were classified as treatment responders (1).<br>**- Certainty of evidence (1):** very low. |
| **Bespoke individual AAT** |  |  |  |  |
| **RCT pilot (1)**<br>(Wijker et al., 2020) | Sample size: 53.<br>Country: Netherlands.<br>Setting: Psychiatric outpatient autism service. | Diagnosis: ASC diagnosis; stress.<br>Demographics: Adults; 55% male.<br>Type of participants: service users. | Bespoke AAT vs Waiting list. | **Feasibility:** High attendance rate, low drop-out rate (1).<br>**Effectiveness:** No significant differences between groups in perceived stress, general psychopathology symptoms and self-esteem (1).<br>**- Certainty of evidence (1):** low. |
| **Bespoke individual RTSM using a mobile platform** |  |  |  |  |
| **Before-and-after comparison (1)**<br>(Hare et al., 2016) | Sample size: 14.<br>Country: UK.<br>Setting: Unclear. | Diagnosis: High-functioning autism diagnosis.<br>Demographics: Adults; 55% men.<br>Type of participants: Service users | Bespoke RTSM using a mobile platform. | **Acceptability:** Limited acceptability (frustration and interference with daily life) (1).<br>**Feasibility:** Low drop-out rate but engagement rate was also low (1).<br>**Effectiveness:** Significant pre-post improvements in aggregated mean anxiety from baseline to post, but not in self-report anxiety and depression (1).<br>**- Certainty of evidence (1):** very low. |
| **Bespoke group CBT for social anxiety** |  |  |  |  |
| **Before-and-after comparison (1)**<br>(Spain et al., 2017) | Sample size: 18.<br>Country: UK.<br>Setting: Autism outpatient psychological therapies service. | Diagnosis: ASC diagnosis.<br>Demographics: Adults; 100% male; 17% non-white.<br>Type of participants: Service users. | Bespoke CBT for social anxiety. | **Acceptability:** Participants found the intervention helpful; some would have preferred to be given additional practical strategies (1).<br>**Feasibility:** high attendance rate, low drop-out rate (1).<br>**Effectiveness:** Significant pre-post improvements in social anxiety, but not in anxiety, depression or social functioning (1).<br>**- Certainty of evidence (1):** very low. |
| **Bespoke group CBT for anxiety** |  |  |  |  |
| **RCT pilot (1)**<br>(Langdon et al., 2016) | Sample size: 52.<br>Country: UK.<br>Setting: Autism, ID and adult mental health services. | Diagnosis: Asperger's syndrome, high-functioning autism or PDD-NOS diagnosis; anxiety.<br>Demographics: CYP and adults; 46-58% male; 96-100% White.<br>Type of participants: Service users. | Bespoke CBT for anxiety + TAU vs waiting list. | **Acceptability:** Participants found the intervention helpful; some noted the need for longer sessions and the possibility of individual delivery (1).<br>**Feasibility:** high attendance rate, low drop-out rate (1).<br>**Effectiveness:** No significant differences between groups in primary (clinician-assessed anxiety) and secondary outcomes (self-report anxiety, depression, social phobia, functioning) (1).<br>**- Certainty of evidence (1):** low. |
| **Bespoke group CBT for anxiety, stress and depression** |  |  |  |  |
| **Non-randomised controlled trial (1)** (McGillivray et al., 2014) | Sample size: 42.<br>Country: Australia.<br>Setting: Disability service agency. | Diagnosis: Asperger's syndrome or high-functioning autism diagnosis; depressed mood, anxiety, stress and/or negative automatic thoughts.<br>Demographics: CYP and adults; 73-81% male.<br>Type of participants: Service users. | Bespoke CBT for anxiety, stress and depression vs waiting list | **Feasibility:** low drop-out rate, attendance rate is unclear (1).<br>**Effectiveness:** No significant differences between groups in mental health outcomes, but those who scored higher in anxiety, depression and stress benefited the most (1).<br>**- Certainty of evidence (1):** low. |

*Note.* **AAT** = Animal-assisted therapy, **ASC** = Autism spectrum condition, **CBT** = Cognitive behavioural therapy, **CYP** = Children and young people, **PDD-NOS** = Pervasive developmental disorder-not otherwise specified, **RTSM** = Real time stress management, **TAU** = treatment as usual, **VR** = Virtual reality.

#### Pilot randomised controlled trials (RCTs)

Two pilot RCTs evaluated bespoke mental health interventions. One pilot cross-over trial reported no significant differences between group CBT for anxiety and waiting list, in primary (clinician-assessed anxiety) and secondary mental health and social outcomes (self-report anxiety, depression, social phobia, functioning) (32). Additionally, attendance and drop-out rates in the CBT group were acceptable and reports from participants showed that the intervention was found helpful (32). Specifically, CBT participants felt supported by others, found listening to others’ problems helpful, and experienced less anxiety. Participants also reported enjoying the interaction with others during the sessions. However, they noted the need for longer sessions, and it was suggested that individual rather than group-based delivery might be positive. Another pilot trial, comparing individual AAT with a waiting list, reported no significant differences between the two groups in all mental health outcomes, however AAT had acceptable attendance and drop-out rates (34). Findings on effectiveness in both trials were inconclusive as these were not sufficiently powered to show an effect.

#### Non-randomised controlled trials

Only one non-randomised controlled trial evaluated bespoke mental health interventions. No significant differences between group CBT and waiting list were found, but subsequent analysis showed that participants who scored higher on measures of depression, anxiety and stress benefited the most from the intervention (35). The study also reported a low drop-out rate, but attendance rate could not be determined, as it was unclear how many completed the intervention (35).

#### Before-and-after comparison studies

Three before-and-after comparison studies investigated bespoke mental health interventions. Significant improvements in outcomes over time were reported (Table 6). However, causality cannot be inferred as there were no comparison groups, hence these will not be reported in detail.

A study of individual CBT for anxiety in combination with Virtual Reality reported low drop-out rates (54). Similarly, a study of group CBT for social anxiety reported acceptable attendance and drop-out rates (57). Qualitative reports from participants indicated that the intervention helped them meet others and feel more confident in social situations, while some stated that they would have benefited from additional practical strategies. Another study examining the use of RTSM reported an acceptable drop-out rate, but the engagement rate was low (52). Participants receiving RTSM indicated acceptable use of the intervention, but also reported frustration when it interfered with their daily lives (52).

### Predictors of outcome

Four studies explored relevant predictors of treatment outcome, including demographic variables, autism diagnosis, co-occurring mental health difficulties and verbal Intelligence Quotient (IQ), but all were shown to have no effect on change in outcomes (Table S12).

## Discussion

The current systematic review aimed to identify strategies used to improve mental health care for autistic adults and examine their feasibility, acceptability and effectiveness. A total of 29 studies were included. A variety of approaches to adapted and bespoke mental health interventions and service improvements were identified.

Most studies reported on strategies that were adapted mental health interventions. These were largely CBT-based, targeting anxiety or a combination of mental health difficulties, with a few targeting depression and OCD. Adaptations to these interventions included mainly communication accommodations, modifications to their structure and content and tailoring treatment to individual differences. We also identified service adaptations aimed to improve mental health care for autistic people and the identification of autism. Adaptations to services included communication accommodations, increasing knowledge of clinicians and detection of autism, environmental adjustments and accommodating individual differences. Bespoke mental health interventions of CBT for social anxiety, CBT for anxiety in combination with VR, AAT and RTSM were also identified.

Evidence on effectiveness of the strategies was generally of low quality and inconclusive, as most included studies lacked a comparison group and RCTs were not sufficiently powered to detect significant differences between groups. Moreover, there were no trials comparing adapted and non-adapted mental health interventions; consequently, the extent to which adaptations were an improvement on standard mental health interventions could not be determined.

Overall, most bespoke and adapted mental health interventions appeared to be acceptable and feasible. Furthermore, qualitative evidence from participants showed that adaptations such as visualisation (50) in CBT for anxiety and the use of simple language and a concrete and structured approach in guided self-help CBT for depression (38) were perceived as helpful. However, the pacing of sessions of guided self-help CBT for depression and the use of pre-defined visual tools received differing views (38). Therapists reported frequently using adaptations related to communication, content of the intervention and adjustment of treatment to individual needs when delivering CBT and EMDR to autistic people (42,43). However, one study with service users reported limited feasibility of adapted CBT for OCD, which could be attributed to difficulties attending sessions regularly (51). Another study demonstrated limited feasibility and acceptability of bespoke RTSM (52), as the unpredictability of the intervention led to frustration and interference with life.

Autism-tailored service pathways and models were found to be acceptable and feasible, suggesting that the introduction of autism-specific training could develop clinician’s knowledge and skills and subsequently improve care. Initiatives to improve the identification of autism also indicated that the use of AQ, RAADS-R and ASD-DF to identify potentially autistic people could be helpful for screening for autism in mental health settings, and subsequent referral to specialist autism assessment (40,41). However, only two studies investigated the detection of autism, despite the perceived misdiagnosis of autism and the impact this may have on treatment (16).

A variety of adaptations to services and interventions that are acceptable and feasible to implement in mental health care were identified. Many of these adaptations have been prioritised by autistic adults (59), including improving communication, providing environmental adjustments and developing knowledge of autism in clinicians. This is a promising finding, as it demonstrates that autistic people’s priorities are being taken into consideration. However, some adaptations (e.g., communication) were more frequently used than others (e.g., environmental adjustments, clinician knowledge, detection of autism), indicating that there may be scope for further improvement in service provision.

Notably, we identified a lack of comprehensive descriptions of how interventions were adapted and why. Furthermore, no tailored pharmacological interventions or prescription initiatives were found, regardless of the common use of psychotropic medication (60), which is perceived as inappropriate by some autistic people (11), highlighting a potential gap. Other notable gaps include evidence on what works for autistic people with ID, suggesting a selection bias against participants with ID (61), and evidence on supporting autistic people with severe or long-term mental health difficulties including high levels of suicidality and self-harm. These are important areas in the context of the disproportionate detention of autistic people to psychiatric wards (62).

The current review also piloted a novel and lived experience researcher-led approach to identifying neurotypical biases in studies. Only 24% of studies involved autistic individuals, hence co-produced research is lacking. Additionally, selection of outcomes measures and data collection methods in many of the papers demonstrated a lack of adjustments to facilitate wider participation, some interventions appeared to focus on masking autistic traits and language used showed a deficit approach to autism (63,64).

### Strengths and limitations

The current review identified a list of adaptations that have been implemented across the service pathway, which may be helpful for tailoring treatments and services to the needs of autistic people. Another key strength is that the review was co-produced with lived experience researchers, who were involved in all aspects of the review, including protocol development, article screening, data extraction and synthesis, neurotypical bias assessment, interpretation of findings, write-up and dissemination. We used the GRADE framework to assess quality of evidence for effectiveness and integrated it within our narrative synthesis.

A limitation is that most studies included in the review had a small sample and lacked a comparison group preventing us from attributing any improvements in outcomes to the intervention alone, and where there were comparisons, they were not with a non-adapted intervention hindering any conclusions on effects of adaptations. A further limitation is that a high proportion of participants were white and male, neglecting under-represented groups such as black and ethnic minorities and other gender identities. Differences associated with sex/gender, ethnicities and cultures may be implicated in the presentation of autistic traits and may impact how adaptations are received by these groups, thus should be taken into consideration (65–68). Additionally, research on different types of strategies (e.g., adaptations to pharmacological interventions) and research including autistic people with ID and severe or long-term mental health difficulties was notably missing.

### Clinical implications and future directions

The current review highlighted a list of strategies evaluated as acceptable and simple to implement, such as improving communication and providing environmental adjustments, that do not necessarily require a further RCT evaluation. An individually tailored approach to treatment may be particularly helpful and facilitate appropriate mental health care, as autistic people differ in their support needs and the presentation of autistic traits (69,70). Nevertheless, a balance should be struck between tailoring treatment to individual differences and adhering to evidence-based practice, which could possibly be addressed through a neurodivergence-informed approach to therapy and primary research (25).

Future research should investigate a co-produced package of mental health service improvement measures. There is also a need for recruitment strategies that increase participation from under-represented groups and reduce biases (e.g., male bias); and for an increased focus on those with ID and/or severe or long-term mental health difficulties.

## Declaration of Interest

None.

## Funding

This paper presents independent research commissioned and funded by the National Institute for Health Research (NIHR) Policy Research Programme and conducted by the NIHR Mental Health Policy Research Unit (MHPRU). The funder has no role in the study design, data analysis, write-up of the manuscript or the decision to submit for publication.

## Author Contribution

The working group collaboratively conceived and formulated the review questions. TS conducted the searches. TP, AG, AK, TS, DS, SL, RC, JG, HB, UF, KS contributed to the screening and TP, AG, AK, TS, DS, UF, SL, JG, HB, AT, MM, GB, RC extracted data. SL, TP and TS collaboratively drafted the manuscript. TS, TP, SL, PB, RC, KS, VT, SJ, NA, RJ, JP, AT, RP, BC, WM, AS, PC, RRO, SZ contributed to the editing of the manuscript. All authors have approved the final manuscript.

## Data Availability

Data were collected from publicly available research papers which are referenced.

## Data Availability

Data were collected from publicly available research papers which are referenced.

## Footnotes

1 Whilst there are different terms used to refer to people on the autism spectrum, in this review we are using identity-first language (e.g., “autistic person”)

## Notes

### Competing Interest Statement

The authors have declared no competing interest.

