## Supplementary material for "Approaches to improving mental health care for autistic people: a systematic review"

**Table S1** PRISMA checklist

| **Section and topic** | **Item No** | **Checklist item** | **Location where item is reported** |
| --- | --- | --- | --- |
| **ADMINISTRATIVE INFORMATION** | | |  |
| Title: |  |  |  |
| Identification | 1a | Identify the report as a protocol of a systematic review | Page 1 |
| Update | 1b | If the protocol is for an update of a previous systematic review, identify as such | Not applicable |
| Registration | 2 | If registered, provide the name of the registry (such as PROSPERO) and registration number | Page 4 |
| Authors: |  |  |  |
| Contact | 3a | Provide name, institutional affiliation, e-mail address of all protocol authors; provide physical mailing address of corresponding author | Page 1 |
| Contributions | 3b | Describe contributions of protocol authors and identify the guarantor of the review | Pages 1 and 20 |
| Amendments | 4 | If the protocol represents an amendment of a previously completed or published protocol, identify as such and list changes; otherwise, state plan for documenting important protocol amendments | Not applicable |
| Support: |  |  |  |
| Sources | 5a | Indicate sources of financial or other support for the review | Page 20 |
| Sponsor | 5b | Provide name for the review funder and/or sponsor | Page 20 |
| Role of sponsor or funder | 5c | Describe roles of funder(s), sponsor(s), and/or institution(s), if any, in developing the protocol | Page 20 |
| **INTRODUCTION** | | |  |
| Rationale | 6 | Describe the rationale for the review in the context of what is already known | Pages 3 and 4 |
| Objectives | 7 | Provide an explicit statement of the question(s) the review will address with reference to participants, interventions, comparators, and outcomes (PICO) | Page 4 |
| **METHODS** | | |  |
| Eligibility criteria | 8 | Specify the study characteristics (such as PICO, study design, setting, time frame) and report characteristics (such as years considered, language, publication status) to be used as criteria for eligibility for the review | Pages 5-7 |
| Information sources | 9 | Describe all intended information sources (such as electronic databases, contact with study authors, trial registers or other grey literature sources) with planned dates of coverage | Pages 4-6 |
| Search strategy | 10 | Present draft of search strategy to be used for at least one electronic database, including planned limits, such that it could be repeated | Tables S2-S4 in supplementary |
| Study records: |  |  |  |
| Data management | 11a | Describe the mechanism(s) that will be used to manage records and data throughout the review | Page 6 |
| Selection process | 11b | State the process that will be used for selecting studies (such as two independent reviewers) through each phase of the review (that is, screening, eligibility and inclusion in meta-analysis) | Pages 6-8 |
| Data collection process | 11c | Describe planned method of extracting data from reports (such as piloting forms, done independently, in duplicate), any processes for obtaining and confirming data from investigators | Pages 6-8 |
| Data items | 12 | List and define all variables for which data will be sought (such as PICO items, funding sources), any pre-planned data assumptions and simplifications | Pages 6 and 7 |
| Outcomes and prioritization | 13 | List and define all outcomes for which data will be sought, including prioritization of main and additional outcomes, with rationale | Pages 6 and 7 |
| Risk of bias in individual studies | 14 | Describe anticipated methods for assessing risk of bias of individual studies, including whether this will be done at the outcome or study level, or both; state how this information will be used in data synthesis | Page 8 and Table S10 in supplementary |
| Data synthesis | 15a | Describe criteria under which study data will be quantitatively synthesised | Not applicable, narrative synthesis was used |
|  | 15b | If data are appropriate for quantitative synthesis, describe planned summary measures, methods of handling data and methods of combining data from studies, including any planned exploration of consistency (such as I^2^, Kendall’s τ) | Not applicable |
|  | 15c | Describe any proposed additional analyses (such as sensitivity or subgroup analyses, meta-regression) | Not applicable |
|  | 15d | If quantitative synthesis is not appropriate, describe the type of summary planned | Not applicable |
| Meta-bias(es) | 16 | Specify any planned assessment of meta-bias(es) (such as publication bias across studies, selective reporting within studies) | Not applicable |
| Confidence in cumulative evidence | 17 | Describe how the strength of the body of evidence will be assessed (such as GRADE) | Not applicable |

**Table S2.** Ovid MEDLINE(R) search

|  | Query | Hits on 20/07/22 |
| --- | --- | --- |
| 1 | (autis* or Asperger* or ASC or ASD or PDD or pervasive development*).ab,kw,ti. | 84191 |
| 2 (MeSH) | Autistic Disorder/ or Autism Spectrum Disorder/ or Child Development Disorders, Pervasive/ or Asperger Syndrome/ | 44574 |
| 3  (MeSH) | Rett Syndrome/ | 2860 |
| 4 | 1 OR 2 OR 3 | 90125 |
| 5 | (mental health service* or health service* or hospital or GP or ward or inpatient* or "community mental health" or home treatment or crisis resolution or "child and adolescent mental health service" or CAMHS or CBT or "cognitive behavio* therap*" or DBT or "dialectical behavio* therap*" or family therapy or interpersonal therap* or psychodynamic or treatment* or intervention* or occupational therapy or mindfulness or psychological intervention* or psychological therap* or behavio* therap* or psychotherap* or "acceptance and commitment therap*" or ACT).ab,kw,ti. | 7210419 |
| 6 | (mental health or mental illness* or mental disorder* or mental condition* or anxi* or affect or depress* or behavio* problems or eating disorder*).ab,kw,ti. | 919484 |
| 7  (MeSH) | mental disorders/ or anxiety disorders/ or "bipolar and related disorders"/ or "disruptive, impulse control, and conduct disorders"/ or dissociative disorders/ or elimination disorders/ or "feeding and eating disorders"/ or mood disorders/ or neurocognitive disorders/ or neurotic disorders/ or personality disorders/ or "schizophrenia spectrum and other psychotic disorders"/ or substance-related disorders/ or "trauma and stressor related disorders"/ | 373099 |
| 8 | 6 OR 7 | 1142253 |
| 9 | 4 and 5 and 8 | 5262 |
| 10 | limit 9 to yr="1994 -Current" | 5073 |

**Table S3.** APA PsycINFO search

|  | Query | Hits on 20/07/22 |
| --- | --- | --- |
| 1  (MeSH) | autism spectrum disorders/ or neurodevelopmental disorders/ or autistic traits/ or developmental disabilities/ or rett syndrome/ | 65622 |
| 2 | (autis* or Asperger* or ASC or ASD or PDD or pervasive development*).ab,hw,id,ti. | 65688 |
| 3 | 1 OR 2 | 79011 |
| 4 | ("mental health" or "mental illness*" or "mental disorder*" or "mental condition*" or anxi* or affective or depress* or "behavio* problems" or "eating disorder*" or psychosis or schizophrenia or "psychotic disorder*").ab,hw,id,ti. | 998005 |
| 5  (MeSH) | mental disorders/ or affective disorders/ or anxiety disorders/ or bipolar disorder/ or borderline states/ or chronic mental illness/ or dissociative disorders/ or eating disorders/ or mental disorders due to general medical conditions/ or neurocognitive disorders/ or neurosis/ or personality disorders/ or psychosis/ or serious mental illness/ or sleep wake disorders/ or somatoform disorders/ or "stress and trauma related disorders"/ or "substance related and addictive disorders"/ | 235303 |
| 6 | 4 OR 5 | 1030648 |
| 7 | ("mental health service*" or "health service*" or hospital or GP or ward or inpatient* or "community mental health" or "home treatment" or "crisis resolution" or "child and adolescent mental health service" or CAMHS or CBT or "cognitive behavio* therap*" or DBT or "dialectical behavio* therap*" or "family therap*" or "interpersonal therap*" or psychodynamic or treatment* or intervention* or "occupational therap*" or mindfulness or "psychological intervention*" or "psychological therap*" or "behavio* therap*" or psychotherap* or "acceptance and commitment" or ACT).ab,hw,id,ti. | 1399839 |
| 8 | 3 AND 6 AND 7 | 9268 |
| 9 | limit 8 to yr="1994 -Current" | 8349 |

**Table S4.** CINAHL Plus search

|  | Query | Hits on 20/07/22 |
| --- | --- | --- |
| 1  (MeSH) | (MH "Autistic Disorder") OR (MH "Rett Syndrome") OR (MH "Developmental Disabilities") OR (MH "Asperger Syndrome") | 25,117 |
| 2 | autis* or Asperger* or ASC or ASD or PDD or pervasive development* | 25,938 |
| 3 | 1 OR 2 | 30,774 |
| 4  (MeSH) | (MH "Mental Disorders") OR (MH "Neurotic Disorders") OR (MH "Affective Disorders") OR (MH "Anxiety Disorders") OR (MH "Dissociative Disorders") OR (MH "Factitious Disorders") OR (MH "Somatoform Disorders") OR (MH "Personality Disorders") OR (MH "Psychotic Disorders") OR (MH "Substance Use Disorders") OR (MH "Psychological Trauma") OR (MH "Adjustment Disorders") OR (MH "Behavioral Symptoms") OR (MH "Behavioral and Mental Disorders") | 149,139 |
| 5 | "mental health" or "mental illness*" or "mental disorder*" or "mental condition*" or anxi* or affect or depress* or "behavio* problems" or "eating disorder*" or psychosis or schizophrenia or "psychotic disorder*" | 264,382 |
| 6 | 4 OR 5 | 362,727 |
| 7 | "mental health service*" or "health service*" or hospital or GP or ward or inpatient* or "community mental health" or "home treatment" or "crisis resolution" or "child and adolescent mental health service" or CAMHS or CBT or "cognitive behavio* therap*" or DBT or "dialectical behavio* therap*" or "family therap*" or "interpersonal therap*" or psychodynamic or treatment* or intervention* or "occupational therap*" or mindfulness or "psychological intervention*" or "psychological therap*" or "behavio* therap*" or psychotherap* or "acceptance and commitment" or ACT | 1,105,132 |
| 8 | 3 AND 6 AND 7 (**Limiters:** 1994-2022; Exclude MEDLINE records) | 2,344 |

**Table S5.** List of all included articles

| 1 | Bemmer ER, Boulton KA, Thomas EE, Larke B, Lah S, Hickie IB, Guastella AJ. Modified CBT for social anxiety and social functioning in young adults with autism spectrum disorder. Molecular Autism. 2021 Dec;12(1):1-5. |
| --- | --- |
| 2 | Blainey SH, Rumball F, Mercer L, Evans LJ, Beck A. An evaluation of the effectiveness of psychological therapy in reducing general psychological distress for adults with autism spectrum conditions and comorbid mental health problems. Clinical psychology & psychotherapy. 2017 Nov;24(6):O1474-84. |
| 3 | Brugha T, Tyrer F, Leaver A, Lewis S, Seaton S, Morgan Z, Tromans S, van Rensburg K. Testing adults by questionnaire for social and communication disorders, including autism spectrum disorders, in an adult mental health service population. International journal of methods in psychiatric research. 2020 Mar;29(1):e1814. |
| 4 | Cooper K, Loades ME, Russell A. Adapting psychological therapies for autism. Research in autism spectrum disorders. 2018 Jan 1;45:43-50. |
| 5 | Dreiling NG, Cook ML, Lamarche E, Klinger LG. Mental health Project ECHO Autism: Increasing access to community mental health services for autistic individuals. Autism. 2022 Feb;26(2):434-45. |
| 6 | Ekman E, Hiltunen AJ. Modified CBT using visualization for autism spectrum disorder (ASD), anxiety and avoidance behavior–a quasi‐experimental open pilot study. Scandinavian journal of psychology. 2015 Dec;56(6):641-8. |
| 7 | Fisher N, van Diest C, Leoni M, Spain D. Using EMDR with autistic individuals: A Delphi survey with EMDR therapists. Autism. 2023 Jan;27(1):43-53. |
| 8 | Flygare O, Andersson E, Ringberg H, Hellstadius AC, Edbacken J, Enander J, Dahl M, Aspvall K, Windh I, Russell A, Mataix-Cols D. Adapted cognitive behavior therapy for obsessive–compulsive disorder with co-occurring autism spectrum disorder: A clinical effectiveness study. Autism. 2020 Jan;24(1):190-9. |
| 9 | Hare DJ, Gracey C, Wood C. Anxiety in high-functioning autism: A pilot study of experience sampling using a mobile platform. Autism. 2016 Aug;20(6):730-43. |
| 10 | Harrison KB, McCredie MN, Reddy MK, Krishnan A, Engstrom A, Posey YS, Morey LC, Loveland KA. Assessing autism spectrum disorder in intellectually able adults with the personality assessment inventory: Normative data and a novel supplemental indicator. Journal of autism and developmental disorders. 2020 Nov;50:3935-43. |
| 11 | Helverschou SB, Bakken TL, Berge H, Bjørgen TG, Botheim H, Hellerud JA, Helseth I, Hove O, Johansen PA, Kildahl AN, Ludvigsen LB. Preliminary Findings From a Nationwide, Multicenter Mental Health Service for Adults and Older Adolescents With Autism Spectrum Disorder and ID. Journal of Policy and Practice in Intellectual Disabilities. 2021 Jun;18(2):162-73. |
| 12 | Horwood J, Cooper K, Harvey H, Davies L, Russell A. The experience of autistic adults accessing adapted cognitive behaviour therapy: ADEPT (Autism Depression Trial) qualitative evaluation. Research in Autism Spectrum Disorders. 2021 Aug 1;86:101802. |
| 13 | Jones K, Gangadharan S, Brigham P, Smith E, Shankar R. Current practice and adaptations being made for people with autism admitted to in-patient psychiatric services across the UK. BJPsych Open. 2021 May;7(3):e102. |
| 14 | Kiep M, Spek AA, Hoeben L. Mindfulness-based therapy in adults with an autism spectrum disorder: Do treatment effects last?. Mindfulness. 2015 Jun;6:637-44. |
| 15 | Langdon PE, Murphy GH, Shepstone L, Wilson EC, Fowler D, Heavens D, Russell A, Rose A, Malovic A, Mullineaux L. The People with Asperger syndrome and anxiety disorders (PAsSA) trial: a pilot multicentre, single-blind randomised trial of group cognitive–behavioural therapy. BJPsych Open. 2016 Mar;2(2):179-86. |
| 16 | Lobregt-van Buuren E, Sizoo B, Mevissen L, de Jongh A. Eye movement desensitization and reprocessing (EMDR) therapy as a feasible and potential effective treatment for adults with autism spectrum disorder (ASD) and a history of adverse events. Journal of autism and developmental disorders. 2019 Jan 15;49:151-64. |
| 17 | Maskey M, Rodgers J, Ingham B, Freeston M, Evans G, Labus M, Parr JR. Using virtual reality environments to augment cognitive behavioral therapy for fears and phobias in autistic adults. Autism in Adulthood. 2019 Jun 1;1(2):134-45. |
| 18 | McGillivray JA, Evert HT. Group cognitive behavioural therapy program shows potential in reducing symptoms of depression and stress among young people with ASD. Journal of autism and developmental disorders. 2014 Aug;44:2041-51. |
| 19 | Oshima F, Murata T, Ohtani T, Seto M, Shimizu E. A preliminary study of schema therapy for young adults with high-functioning autism spectrum disorder: a single-arm, uncontrolled trial. BMC Research Notes. 2021 Dec;14(1):1-8. |
| 20 | Pahnke J, Hirvikoski T, Bjureberg J, Bölte S, Jokinen J, Bohman B, Lundgren T. Acceptance and commitment therapy for autistic adults: An open pilot study in a psychiatric outpatient context. Journal of Contextual Behavioral Science. 2019 Jul 1;13:34-41. |
| 21 | Petty S, Bergenheim ML, Mahoney G, Chamberlain L. Adapting services for autism: Recommendations from a specialist multidisciplinary perspective using freelisting. Current Psychology. 2021 Jul 16:1-2. |
| 22 | Russell AJ, Jassi A, Fullana MA, Mack H, Johnston K, Heyman I, Murphy DG, Mataix‐Cols D. Cognitive behavior therapy for comorbid obsessive‐compulsive disorder in high‐functioning autism spectrum disorders: A randomized controlled trial. Depression and Anxiety. 2013 Aug;30(8):697-708. |
| 23 | Russell A, Gaunt D, Cooper K, Barton S, Horwood J, Kessler D, Metcalfe C, Ensum I, Ingham B, Parr JR, Rai D. The feasibility of low intensity psychological therapy for co-occurring depression in adult Autism: The ADEPT study-a pilot randomised controlled trial. Autism. 2020 Aug 1;24(6):1360-72. |
| 24 | Sizoo BB, Kuiper E. Cognitive behavioural therapy and mindfulness based stress reduction may be equally effective in reducing anxiety and depression in adults with autism spectrum disorders. Research in developmental disabilities. 2017 May 1;64:47-55. |
| 25 | Spain D, Blainey SH, Vaillancourt K. Group cognitive behaviour therapy (CBT) for social interaction anxiety in adults with autism spectrum disorders (ASD). Research in Autism Spectrum Disorders. 2017 Sep 1;41:20-30. |
| 26 | Spek AA, Van Ham NC, Nyklíček I. Mindfulness-based therapy in adults with an autism spectrum disorder: a randomized controlled trial. Research in developmental disabilities. 2013 Jan 1;34(1):246-53. |
| 27 | Tchanturia K, Dandil Y, Li Z, Smith K, Leslie M, Byford S. A novel approach for autism spectrum condition patients with eating disorders: Analysis of treatment cost‐savings. European Eating Disorders Review. 2021 May;29(3):514-8. |
| 28 | Wijker C, Leontjevas R, Spek A, Enders-Slegers MJ. Effects of dog assisted therapy for adults with autism spectrum disorder: An exploratory randomized controlled trial. Journal of autism and developmental disorders. 2020 Jun;50:2153-63. |
| 29 | Wise JM, Cepeda SL, Ordaz DL, McBride NM, Cavitt MA, Howie FR, Scalli L, Ehrenreich-May J, Wood JJ, Lewin AB, Storch EA. Open trial of modular cognitive-behavioral therapy in the treatment of anxiety among late adolescents with autism spectrum disorder. Child Psychiatry & Human Development. 2019 Feb 15;50:27-34. |

**Table S6.** Study design and study population characteristics

| **Author (Year)** | **Country** | **Aim** | **Study design** | **Setting** | **Baseline N** | **Participants** | **Strategy** | **Comparison** |
| --- | --- | --- | --- | --- | --- | --- | --- | --- |
| Bemmer et al. (2021) | Australia | Evaluate the benefit, tolerability and acceptability of adapted CBT for social anxiety | Pre-post | Research clinic within primary health care network and headspace clinical services | 84 | 100% ASC diagnosis  CYP and adults; mean age = 22.77 (SD = 5.31), range 16-38; 60% male | Adapted CBT for social anxiety  Group; face-to-face; 2.5 hours x 8 weekly sessions. Facilitated by staff with prior experience working with autistic people. 60-minute debrief following each session. | N/A |
| Blainey et al. (2017) | England | Assess the effectiveness of routinely offered psychological therapy in reducing psychological distress | Service evaluation | Autism psychological therapies services | 81 | 100% ASC diagnosis; co-occurring difficulties: 50.6% anxiety disorders, 13.6% depression, 22% co-occurring anxiety and mood disorders, 7% psychotic experiences, 5% features of personality disorder  Adult; mean age = 30 (SD = 10.64); 74.1% male; 40.7% non-White British | Adapted CBT  Individual; face-to-face; ≥ 20 hours delivered by CBT therapists. | N/A |
| Brugha et al. (2020) | England | Validate AQ and RAADS-R in mental health services | Cross-sectional two-phase survey | Inpatient, outpatient and community mental health services | 738 | 1 % had a clinical ASC diagnosis; 4.8% met cut-off criteria for ASC; co-occurring difficulties: 22.6% mood disorder, 8.7% psychotic disorders, 8.3% personality disorder, 5.8% somatoform disorders, 9.9% mental and behavioural disorders due to alcohol/substance misuse, 0.7% ADHD, 1.6% other  Adults; 42.3% <40 years, 29.8% 40-49 years and 27.9% 50+ years; 49.6% male | Detection of autism  Examine whether self-report tools (AQ and RAADS-R) can discriminate between autistic and non-autistic people. | N/A |
| Cooper at al. (2018) | UK | Investigate therapists’ knowledge and experience of working within a CBT framework with autistic people | Cross-sectional survey | IAPT and secondary mental health services | 50 | Therapists working with autistic people across the lifespan. | Adapted CBT  Delivered by psychological therapists. | N/A |
| Dreiling et al. (2022) | USA | Develop and implement Project ECHO for community mental health providers | Non-randomised service implementation | Community services | 86 | Mental health providers working with autistic people across the lifespan.  mean age = 42.22 (SD = 10.6), range 25-66; 6% male; 14% non-white | Project ECHO (tele-mentoring platform) to connect primary care providers to increase knowledge of autism and co-occurring mental health difficulties, and appropriately adapt treatments.  90 minutes x 10 bi-weekly virtual sessions over a period of 6 months. Panel consisted of two senior psychologists and 1 clinician and 1 parent advocate/professional autism resource specialist. | N/A |
| Ekman et al. (2015) | Sweden | Investigate benefits of adapted CBT for anxiety using visualisation and communication | Quasi-experimental open pilot | Psychiatric clinic, treatment centre for youth and private clinic | 18 | 100% ASC diagnosis; 100% diagnosis of anxiety and avoidance behaviour: 33.3% social phobia, 27.8% OCD, 5.6% eating disorder, 16.7% combination of difficulties  CYP and adults; mean age for teens = 14.9 (SD = 1.5), range 13-17, 22.2% male. Mean age for adults = 29.8 (4.4), range 23-36, 38.9% male | Adapted CBT for anxiety  Individual; face-to-face; 45-60 min x 15 sessions every second week or at client's convenience. Delivered by three CBT therapists with prior experience working with autistic people. | N/A |
| Fisher et al. (2023) | Netherlands; UK | Develop therapist consensus about adaptations to EMDR that are important when working with autistic individuals | Delphi Survey (3 rounds) | Psychological therapies, community mental health, intellectual disability, forensic and tertiary services, independent practice, education, military, voluntary organisations | 103^c^ | EMDR therapists working with autistic people across the lifespan. | Adapted EMDR  Delivered by trained therapists. | N/A |
| Flygare et al. (2020) | Sweden | Evaluate adapted CBT for OCD in autistic adults | Non-randomised clinical effectiveness | Specialist outpatient OCD clinic | 19 | 100% Asperger’s syndrome diagnosis; 100% OCD diagnosis; co-occurring difficulties: 26.3% depression, 15.8% GAD, 26.3% ADHD, tic disorder 26.3%, 10.5% panic disorder, 10.5% SAD, 5.3% PTSD, 5.3% substance dependence, 42.1% bulimia nervosa  Adult; mean age = 23.84 (SD = 5.90); 42% male | Adapted CBT for OCD  Individual; 20 weekly sessions. Face-to-face in the OCD clinic. Delivered by clinical psychologists with prior experience working with autistic people. | N/A |
| Hare et al. (2016) | UK | Act as a ‘proof of principle’ study with regard to the use of personal digital assistants to deliver RTSM for everyday stress | Case series | Unclear | 14 | 100% high-functioning autism diagnosis  Adults; 55% men | Bespoke RTSM using a mobile platform  Individual; ESM approach; delivered when high anxiety scores were reported to questionnaires which were delivered 10 times during the day. | N/A |
| Harrison at al. (2020) | USA | Introduce and provide validation for a novel discriminant function for identification of ASD-like symptomatology | Retrospective analytical cross-sectional | Autism outpatient clinic and inpatient psychiatric hospital | 1487 | 11.4% ASC diagnosis (n = 169)  Adults; mean age = 33.33 (SD = 12.04), range 18-66; 64% male, 24% non-White | Detection of autism  Identify an ASD-DF derived from self-report screening tool PAI, which can discriminate autism in a clinical population. | Contrast inpatient sample (n = 72), PAI clinical standardised sample (n = 1246) |
| Helverschou et al. (2021) | Norway | Describe patterns of psychiatric and behaviour problems in autistic people with ID, treated by the AUP network | Service evaluation/implementation | Specialist hospital-level mental health services | 132 | 100% ASC diagnosis; 100% ID diagnosis; co-occurring difficulties: 32.6% psychosis, 50.8% depression, 44.7% anxiety, 15.9% OCD  CYP and adults; mean age = 28.6 (SD = 10.6), range 16-66; 67% male | AUP network for professionals to improve access to and quality of tailored services for autistic adults with ID and increase knowledge of how psychiatric disorders present in autism.  Face-to-face across 8 clinics. Network meetings occur yearly over 6 days, and seminars occur every other year for 2 days. Delivered by professionals. | N/A |
| Horwood et al et al. (2021) | UK | Investigate the acceptability of adapted guided self-help CBT for depression and TAU | Qualitative (part of a pilot RCT) | Autism services | 26 | 100% ASC diagnosis; 100% depression  Adults; mean age = 40, range 21-58; 81% male; 100% white British  Staff (n = 5) | Adapted guided self-help CBT for depression (n = 14)  Individual; largely face-to-face; 30-45 min x 9 weekly sessions. Delivered by low-intensity therapists | TAU (n = 7) |
| Jones et al. (2021) | UK | Explore skills and adaptations to inpatient units | Cross-sectional survey – service evaluation | Inpatient units | 90 | Staff, some had experience working with autistic people across the lifespan. | Evaluation of strategies and adaptations to inpatient care | N/A |
| Kiep et al. (2015) | Netherlands | Examine the effects of MBS on psychological and physical wellbeing of autistic individuals | Non-randomised | Autism centre | 58 | 100% ASC diagnosis; 100% symptoms of anxiety, depression and/or rumination.  Adults; 68% males with mean age = 42.1 (SD = 10.5), 32% females with mean age= 37.9 (SD = 14) | Adapted MBT-AS    Group; face-to-face; 9 weekly sessions, with each session lasting 2.5 hours + 40-60 minutes at-home meditation on 6 out of 7 days of the week. Delivered by trained psychologist. | N/A |
| Langdon et al. (2016) | UK | Examine whether bespoke CBT for anxiety is feasible and likely to be efficacious | Pilot single-blind RCT | Autism, intellectual disability and adult mental health services | 52 | 100% Asperger’s syndrome, high-functioning autism or PDD-NOS diagnosis; 100% anxiety symptoms  CYP and adults; Treatment: mean age 33.1 (SD = 14.6), range 20-64; 46% male; 96% White British. Control: mean age 38.7 (SD = 14.3), range 17-65; 58% male; 100% White British | Bespoke CBT for anxiety + TAU (n = 26)  Group; face-to-face; 1h x 24 weekly sessions (participants received 3 initial sessions of 1:1 CBT, followed by 21 group CBT sessions). Delivered by clinical psychologist or CBT therapist | Waiting list (n = 26) |
| Lobregt-van Buuren et al. (2019) | Netherlands | Investigate whether EMDR has the potential to be a treatment for autistic adults and whether EMDR adjunct to TAU is feasible | Non-randomised add-on design | Outpatient/community services | 27 | 100% Autism, Asperger’s syndrome or PDD-NOS diagnosis; 100% a clear link between PTSD symptoms and adverse events; co-occurring difficulties: 66.6% PTSD, 42.9% depression, 23.8% ADHD, 14.3% personality disorder, 9.5% other  Adults; M age = 34.48 (SD = 11.73); 62% male | Adapted EMDR + TAU  Individual; face-to-face; 75 min x up to 8 weekly or bi-weekly sessions. Delivered by trained therapists. | 6-8 weeks TAU (participants were their own controls) |
| Maskey et al. (2019) | UK | Investigate the feasibility and acceptability of CBT for anxiety in combination with virtual reality | Pilot study | Autism services, autism support network | 8 | 100% ASC diagnosis; 100% specific or social phobias  Adults; mean age = 29.8, range 18.8-57; 5% male | Bespoke CBT for anxiety in combination with virtual reality  Individual; face-to-face; 20-30 min x 2 weekly visits (in total 4 sessions). Delivered by a psychologist. | N/A |
| McGillivray et al. (2014) | Australia | Test CBT aimed to reduce negative and anxious thinking patterns and symptoms of stress, anxiety and depression | Quasi-experimental | Disability service agency | 42 | 100% Asperger’s syndrome or high-functioning autism diagnosis; 100% symptoms of depressed mood, anxiety, stress and/or negative automatic thoughts  CYP and adults; treatment: mean age = 20.27 (SD = 4.39), 73% male; waiting list: mean age = 20.50 (SD = 3.4), 81% male aged 15–25 years | Bespoke CBT for anxiety, stress and depression (n = 26)  Group; face-to-face; 2h x 9 weekly sessions. Delivered by a psychologist. | Waiting list (n = 16) |
| Oshima et al. (2021) | Japan | Examine feasibility and acceptability of schema therapy and effects on quality of life and social adjustment | Single-arm preliminary study | University hospital | 12 | 100% ASC diagnosis; 8.3% ADHD, 33.3% OCD, 41.7% depression^a^  Adults; mean age =26.8 (SD = 6.39), range 20-39; 50% male^a^ | Adapted schema therapy  Individual; face-to-face; 50 min x 25 weekly sessions, plus a follow-up session after 12 weeks. | N/A |
| Pahnke et al. (2019) | Sweden | Evaluate preliminary feasibility and potential utility of ACT | Open pilot study | Psychiatric clinic | 10 | 100% Asperger’s syndrome diagnosis; co-occurring difficulties: 10% dysthymia, 40% depression, 20% GAD, 10% OCD, 50% ADHD, 10% Tourette’s syndrome  Adults; mean age = 49 (SD = 12) | Adapted ACT  Group; face-to-face; 150 min x 12 weekly sessions. Delivered by a clinical psychologist with prior experience working with autistic people and a graduate student. | N/A |
| Petty et al. (2021) | UK | Inform service development within mental health services for autistic clients | Qualitative | Specialist autism service | 15 | Staff working with autistic people across the lifespan  Age range 25-44; 80% female | Evaluation of adaptations to improve mental health care. | N/A |
| Russell et al. (2013) | UK | Evaluate adapted CBT for OCD | Single-blind RCT | Specialist autism, OCD clinics and mental health services | 46 | 100% ASC diagnosis; 100% OCD diagnosis  CYP and adults; Treatment: mean age = 28.6 (SD = 11.3), range 14-49; 82.6% male. Control: mean age = 25.2 (SD = 13.5), range 14-65; 69.6% male | Adapted CBT for OCD (n = 23)  Individual; face-to-face; 60 minutes x up to 20 planned sessions. Delivered by psychologists with prior experience in treating OCD. | Adapted anxiety management matched for duration and amount of therapist contact (n = 23) |
| Russell et al. (2020) | UK | Investigate the feasibility of guided self-help CBT for depression, estimate recruitment and retention rates and identify most appropriate outcome measure for a powered RCT | Pilot single-blind RCT | Autism services | 70 | 100% ASC diagnosis; 100% met cut-off criteria for depression; primary diagnosis:  70% depression, 12.8% GAD, 2.8% mixed anxiety and depression, 2.8% specific phobia, 4.3% panic disorder, 1.4% agoraphobia^b^  Adults; Treatment: mean age = 35.3 (SD = 13.6), 69% male; 6% non-white. Control: mean age = 40.2 (SD = 12.6), 77% males; 6% non-white | Adapted guided self-help CBT for depression (n = 35)  Individual; largely face-to-face; 30-45 min x 9 weekly sessions. Delivered by graduate-level low-intensity therapists. | TAU (n = 35) |
| Sizoo et al. (2017) | Netherlands | Investigate whether adapted MBSR and adapted CBT are equally effective in reducing anxiety and depression | Non-randomised controlled | Outpatient psychiatric clinic | 59 | 100% ASC diagnosis; 100% met cut-off criteria for anxiety and/or depression  Adults; Treatment: mean age 35.1 (SD = 9.22); 70% male. Control: mean age 39.4 (SD = 10.81); 59% male | Adapted MBSR (n = 32) and adapted CBT for anxiety and depression (n = 27)  Both treatments: Group; face-to-face; 90 min x 13 weekly sessions. | MBSR and CBT were compared |
| Spain et al. (2017) | UK | Describe the development and evaluation of CBT for anxiety | Non-randomised single arm | Autism outpatient psychological therapies service | 18 | 100% ASC diagnosis  Adults; mean age = 31 (SD = 7.9), range 22-48; 100% male; 17% non-white British | Bespoke CBT for social anxiety  Group; face-to-face; 2h x 11 weekly sessions. Delivered by two members, either a trainee clinical psychologist, clinical psychologist or nurse consultant. | N/A |
| Spek et al. (2013) | Netherlands | Investigate whether MBT-AS may be beneficial in treating co-occurring affective symptoms | RCT | Autism centre | 42 | 100% ASC diagnosis; 100% symptoms of anxiety, depression and/or rumination  Adults; Treatment group: mean = 40.1 (SD = 11); 67% male. Control group: mean = 44.4 (SD = 11.1); 65% male | Adapted MBT-AS (n = 21)  Group; face-to-face; 2.5h x 9 weekly sessions. Delivered by trained therapists. | Waiting list (n = 21) |
| Tchanturia et al. (2021) | UK | Explore the impact of PEACE pathway on length and cost of hospital admissions | Service evaluation/ implementation | Inpatient wards for eating disorders | Not reported | 100% ASC diagnosis; 100% eating disorders | PEACE pathway to improve care for individuals with eating disorders and co-occurring ASC, through introducing autism-specific training, creating a more ASC-friendly ward and supporting sensory difficulties and communication.  20 face-to-face training events delivered by autism experts and investment in materials necessary to create a more autism-friendly ward environment within eating disorder clinic over a three-year period | Autistic vs non-autistic individuals in an eating disorder clinic |
| Wijker et al. (2020) | Netherlands | Explore the effects of AAT | Exploratory single-blind RCT | Psychiatric outpatient service for autism | 53 | 100% ASC diagnosis; 100% met cut-off criteria for stress  Adults; 36% 18–32 years, 30% 33–46 years, 34% 47–60 years; 55% male | Bespoke AAT (n = 27)  Individual; face-to-face; 60 min x 10 weekly sessions. Delivered by therapists with prior experience working with autistic people. | Waiting list (n = 26) |
| Wise et al. (2019) | USA | Develop and examine the feasibility of adapted CBT for anxiety | Open trial | University-based health clinic specialising in the treatment of anxiety | 7 | 100% ASC diagnosis; 100% panic disorder, GAD, social phobia or OCD  CYP and adults; mean age = 17.14 (SD = 1.68), range 16-20; 57.1% male; 14% non-white | Adapted CBT for anxiety  Individual; face-to-face; 60 minutes x 16 weekly sessions. Offered optional work-readiness program. Delivered by doctoral level students in clinical psychology. | N/A |

*Note*. Where ethnicity, gender, mental health condition, age range and standard deviation (SD) is not listed in the table this means that this was not reported in the paper. **AAT** = animal-assisted therapy, **ACT** = Acceptance and commitment therapy, **ADHD** = Attention deficit hyperactive disorder, **ASC** = Autism spectrum condition, **ASD-DF** = Autism-spectrum disorder – discriminant function, **AQ** = Autism quotient, **AUP** = Autism, intellectual disability and psychiatric disorder, **CBT** = Cognitive behavioural therapy, **CYP** = Children and young people, **ECHO** = Extension for community healthcare outcomes, **EMDR** = Eye movement desensitisation and reprocessing, **ESM** = Experience sampling method, **GAD** = Generalised anxiety disorder, **IAPT** = Improving access to psychological therapies, **ID** = Intellectual disability, **MBSR** = Mindfulness-based stress reduction, **MBT-AS** = Mindfulness-based therapy for autism spectrum disorders, **OCD** = Obsessive compulsive disorder, **PAI** = Personality assessment inventory, **PDDS-NOS** = Pervasive developmental disorder - not otherwise specified, **PEACE** = Pathway for eating disorders and autism developed from clinical experience, **PTSD** = post-traumatic stress disorder, **RAADS-R** = Ritvo autism asperger diagnostic scale-revised, **RTSM** = Real-time stress management, **SAD** = social anxiety disorder, **SD** = Standard deviation, **TAU** = Treatment as usual.

^a^ Participant characteristics for those who dropped-out were not reported (n = 2).

^b^ Primary diagnosis was not reported for 1 participant.

^c^ Round 1 n = 103, round 2 n = 43, round 3 n = 26.

**Table S7.** Mixed Methods Appraisal Tool (MMAT) quality assessment

| **Authors** | **Criterion 1** | **Criterion 2** | **Criterion 3** | **Criterion 4** | **Criterion 5** | **Total** |
| --- | --- | --- | --- | --- | --- | --- |
| Bemmer et al. (2021) | Can’t tell | Yes | Yes | Yes | Yes | 4 |
| Blainey et al. (2017) | Yes | Yes | Yes | No | Can’t tell | 3 |
| Brugha et al. (2020) | Yes | Yes | Yes | No | Yes | 4 |
| Cooper et al. (2018) | Yes | Yes | Can’t tell | Yes | Yes | 4 |
| Dreiling et al. (2022) | Yes | Can’t tell | No | No | Yes | 2 |
| Ekman et al. (2015) | Can’t tell | No | Can’t tell | No | Can’t tell | 0 |
| Fisher et al. (2023) | Yes | Yes | Yes | Yes | Can’t tell | 4 |
| Flygare et al. (2020) | Yes | Yes | No | Yes | No | 3 |
| Hare et al. (2016) | Can’t tell | Yes | No | No | Can’t tell | 1 |
| Harrison et al. (2020) | Yes | Yes | Yes | Can't tell | Yes | 4 |
| Helverschou et al. (2021) | Can’t tell | Yes | Yes | Can’t tell | Yes | 3 |
| Horwood et al. (2021) | Yes | Yes | Yes | Yes | Yes | 5 |
| Jones et al. (2021) | Yes | No | Can’t tell | Can’t tell | Yes | 2 |
| Kiep et al. (2015) | Can’t tell | Yes | Yes | No | Yes | 3 |
| Langdon et al. (2016) | Yes | Yes | Yes | Yes | Yes | 5 |
| Lobregt-van Buuren et al. (2019) | Yes | Yes | Yes | Yes | Yes | 5 |
| Maskey et al. (2019) | Can’t tell | Yes | Yes | No | Yes | 3 |
| McGillivray et al. (2014) | Can’t tell | Yes | Yes | Yes | Yes | 4 |
| Oshima et al. (2021) | Yes | Yes | Yes | Yes | Yes | 5 |
| Pahnke et al. (2019 | Yes | Yes | Yes | No | Yes | 4 |
| Petty et al. 2021) | Yes | Yes | Yes | Yes | Yes | 5 |
| Russell et al. (2013) | Yes | Yes | Yes | Yes | Yes | 5 |
| Russell et al. (2020) | Yes | Yes | No | Yes | Yes | 4 |
| Sizoo et al. (2017) | Can’t tell | Yes | Can’t tell | Yes | Can’t tell | 2 |
| Spain et al. (2017) | Yes | Yes | Yes | Yes | No | 4 |
| Spek et al. (2013) | Yes | Yes | Yes | Yes | Yes | 5 |
| Tchanturia et al. (2021) | Yes | Yes | Yes | Can’t tell | Yes | 4 |
| Wijker et al. (2020) | Yes | Yes | Yes | Yes | Yes | 5 |
| Wise et al. (2019) | No | Yes | Yes | No | Yes | 3 |

**Table S8.** Neurotypical biases

| **Author (Year)** | **Any reported involvement from people with lived experience in the design, conduct, or writing up of the study?** | **For studies with qualitative elements, were adjustments made to the data collection process to facilitate wide participation e.g., allowing non-verbal/non-oral communication for interviews?** | **For studies with quantitative elements, were adjustments made to the data collection tools to facilitate wide participation e.g., adapting Likert scales for greater precision, straightforward language, defining key terms?** | **For studies with quantitative elements, were any of the relevant outcome measures to the review adapted (or reported to have been validated) for people with autism (e.g., measure of autistic quality of life)?** | **For studies with quantitative elements, did the intervention/strategy involve any focus (not just related to the relevant measures to the review) on getting people to mask/change autistic behaviours?** |
| --- | --- | --- | --- | --- | --- |
| Bemmer et al. (2021) | None reported | None reported. | None reported | Yes: some measures were reported as previously validated in research with autistic people: "[Depression Anxiety Stress Scale (DASS-21)] is a self-report measure of depression, anxiety and stress, and assesses symptom severity over the past week, and has recently been validated for use in ASD populations". Other measures were reported as previously used with autistic people: "The[Liebowitz Social Anxiety Scale (LSAS-SR)] is one of the most commonly used measures of social anxiety in adult ASD populations"; "[the Kessler psychological distress scale (K10)] has been used in similar studies to measure overall symptoms of distress, rather than disorder-specific (anxiety/depression) symptoms in adults with ASD"; "The [Social Interaction Anxiety Scale (SIAS) and Social Phobia Scale (SPS)] are partner measures used to assess social anxiety and have previously been used to measure SAD levels in ASD populations". | Yes: The primary outcome measure was Social Responsiveness Scale-2—Adult Self-Report (SRS-2), which assessed social skill functioning and autistic symptoms in adults and measures a reduction in autistic behaviours as a positive outcome. Therefore, reducing autistic traits and potentially getting autistic people to mask was a focus of the study. |
| Brugha et al. (2020) | None reported | Not applicable. | None reported. | Yes: all measures used were reported as previously validated in research with autistic people although to varying degrees of success (e.g., the Ritvo autism–Asperger's diagnostic scale-revised (RAADS-R)). | Not applicable. |
| Blainey et al. (2017) | None reported | Not applicable. | None reported | No: The study reports that the measure Clinical Outcomes in Routine Evaluation-Outcome (CORE-OM) has not been validated specifically within autistic population. | Unclear: The study reports a limitation of CORE-OM in that it can be hard for autistic people to identify and label feelings, which could explain increases in scores as people become more psychologically/emotionally aware upon engaging in therapeutic work. It could be inferred that masking is also a feature here, but it is not clear. The study also highlighted the potential difficulty in interpreting questions, which may have also influenced autistic participants' scores on CORE-OM. |
| Cooper at al. (2018) | None reported | Not applicable. | None reported. Data were collected from providers, no reporting of whether these providers were autistic. | Not reported. Data were collected from providers, no reporting of whether these providers were autistic. | Not applicable. |
| Dreiling et al. (2022) | Yes: The multidisciplinary expert panel (“hub team”) who facilitated the Project ECHO included one regional parent advocate to share a parent perspective with participating providers. In each cohort, the parent advocate was recruited from the cohort’s local region to provide information about local resources. Additionally, the specific didactic topics (in the Project ECHO curriculum) were chosen based on feedback from a series of focus groups with rural families who provided input on the mental health needs of their autistic child. | None reported. Data were collected from providers, no reporting of whether these providers were autistic. | None reported. Data were collected from providers, no reporting of whether these providers were autistic. | Not reported. Data were collected from providers, no reporting of whether these providers were autistic. | Not applicable. |
| Ekman et al. (2015) | None reported | Not applicable. | None reported. | Not reported. | Yes: The study's results showed improvement in psychological, social and occupational functioning ability on the Global Functioning Rating scale (GAF scale). The GAF scale prompts include consideration of 'meaningful social relationships' which leaves it open to neurotypical bias and to an encouragement of masking or changing autistic behaviours because increased social interaction is then seen as a positive. In some cases, increased social interaction may not be what the autistic person wants and may not be meaningful to them but may be a result of masking behaviour. |
| Fisher et al. (2023) | Yes: study reported seeking informal feedback about the scope and aims of the first survey, categories of questions included and barriers to EMDR from one autistic adult. Additionally, the study reported informally discussing the study findings and interpretation with two autistic adults and a parent/carer of an autistic child. Participants also included autistic therapists. | Not reported. Data were collected from providers, and participants included autistic therapists. | None reported. Data were collected from therapists, including autistic therapists | Not reported. Data were collected from therapists, including autistic therapists. | Not applicable. |
| Flygare et al. (2020) | None reported | Not applicable. | None reported. | Yes: Obsessive Compulsive Inventory-Revised (OCI-R) was reported to have good sensitivity and specificity in use with autistic adults | No: There is no evidence to suggest a focus on masking or autism-related outcomes specifically. |
| Hare et al. (2016) | None reported | None reported. | None reported. Study reported that participants had 60 seconds to respond to a beep to complete a questionnaire. | No: The study only reported that Hospital Anxiety and Depression Scale (HADS) has been used previously in research with people with high-functioning autism, but no indication of their psychometric properties for autistic individuals was given. | No: There is no evidence to suggest a focus on masking or autism-related outcomes specifically. |
| Harrison at al. (2020) | None reported | Not applicable. | None reported. | Not reported. | Not applicable. |
| Helverschou et al. (2021) | None reported | Not applicable. | None reported. Measures completed by caregivers. | Yes: the study reported the Psychopathology in Autism Checklist (PAC) to be a specific screening checklist for identification of individuals with autism and intellectual disability in need of psychiatric services, and a psychiatric instrument developed specifically for autistic individuals that has been found to discriminate reliably between psychiatric symptoms and the core autistic characteristics. The study reported that PAC has also been found to distinguish between autistic adults and those with intellectual disabilities with and without psychiatric conditions, and to a certain extent between people with different psychiatric conditions, especially psychosis and obsessive-compulsive disorder. | Yes: The use of the Aberrant Behaviour Checklist (ABC) focuses on behaviour that is challenging to people around the autistic person and measures a reduction in these behaviours as a positive outcome. Therefore, reducing autistic traits and potentially getting autistic people to mask was a focus of the study. Additionally, the significant correlation between PAC and ABC suggests that reducing 'challenging' behaviour may lead to change the response of others around the person, and may thereby reduce feelings linked to depression/anxiety, indicating potential masking. |
| Horwood et al et al. (2021) | Yes: feedback of two autistic adults informed two iterations of the design of the intervention session materials during the development phase. No other mention of involvement of people with lived experience in the design, conduct or writing-up of the study. | None reported. | Not applicable. | Not applicable. | Not applicable. |
| Jones et al. (2021) | None reported | Not reported. Data were collected from providers, no reporting of whether these providers were autistic. | None reported. | Not reported. Data were collected from providers, no reporting of whether these providers were autistic. | Not applicable. |
| Kiep et al. (2015) | None reported | Not applicable. | None reported. | Not reported. All measures were for non-specific populations. | No: There is no evidence to suggest a focus on masking or autism-related outcomes specifically. |
| Langdon et al. (2016) | None reported | None reported. | None reported. | Not reported. | No: There is no evidence to suggest a focus on masking or autism-related outcomes specifically. |
| Lobregt-van Buuren et al. (2019) | None reported | Not applicable. | Yes: The Adapted Anxiety Disorders Interview Schedule-Children (ADIS-C) PTSD version for adults used in the study was described to be implemented in the following way, which indicates an adjustment to the data collection tool: "[The difficulty to spontaneously share relevant information for autistic people] was addressed by making use of the concrete, visualized and structured way in which trauma, adverse events and trauma related symptoms are probed by the Adapted ADIS-C section PTSD (version for adults with mild to borderline intellectual disabilities), such that also in adults with ASD unprocessed memories could be identified. This instrument seemed to be appropriate to investigate trauma history and trauma related symptoms in adults with ASD." | Yes: The study reported using the Adapted ADIS-C section PTSD version for adults that is a semi-structured interview to assess trauma, adverse events and trauma related symptoms in adults with mild to borderline intellectual disabilities. The study reported that psychometric properties of this instrument have been studied in adults with mild to borderline intellectual disabilities, some of whom were also autistic. | No: the focus of the intervention was around processing trauma. The Social Responsiveness Scale-Adult version (SRS-A) was used as an outcome measure looking at changes in autistic traits, however there was not an implication that reducing autistic traits was a goal, focus or positive outcome. |
| Maskey et al. (2019) | None reported | Not applicable. | None reported | Not reported. | No: The focus of the study was on phobias and there was no discussion of reduction in autistic traits as a focus or positive outcome. Outcomes were measured based on participant reports and observed behaviours/reduction in safety behaviours around their phobia. |
| McGillivray et al. (2014) | None reported | Not applicable. | None reported | Not reported. | No: the intervention did not focus on getting people to mask or change autistic behaviours and was focused on what worked for autistic people. The Depression Anxiety Stress Scales (DASS) measure in particular was good because it measured physical responses to anxiety rather than behaviours, reducing the potential of measuring masking behaviours as a positive outcome. |
| Oshima et al. (2021) | None reported | Not applicable. | None reported | Not reported. | No: the intervention did not focus on changing behaviour, but on reducing maladaptive schemas. |
| Pahnke et al. (2019) | None reported | Not applicable. | None reported. | Not reported. | No: The focus of the intervention appeared to be on mindful awareness of sensations and around managing stress in social situations rather than on changing social behaviour. The definition of psychological flexibility here is not the same as 'reduction in black and white thinking' or 'reducing rigidity' and seems to be more about being aware of thoughts and sensations which therefore isn't getting people to mask autistic behaviours. Potentially looking at 'reducing social impairment' could lead to a focus on getting people to mask autistic traits, but as the functional impairment scale was self-rated and only had three items (how much does your disability impact on these situations) rather than going into specific social behaviours the focus isn't on 'did the autistic behaviour reduce', rather it's on 'do I feel like I can cope better?'. |
| Petty et al. (2021) | Yes: the study reported that all study materials were revised by people with lived experience of autism, including the outcome measures, the design of the study and the interpretation of the findings. | Not reported. Data were collected from providers, no reporting of whether these providers were autistic. | Not applicable. | Not applicable. | Not applicable. |
| Russell et al. (2013) | None reported | Not applicable. | Yes: as described - "at the start of each clinical interview, care was taken to ensure that the participant was cognisant of the phenomena to be rated, that the discomfort and anxiety basis for each potential [obsessive-compulsive] symptom was clearly established using visual tools if necessary. Eliciting of symptoms was achieved if needed by enquiring about daily routines in total before gathering further phenomenological information. Communication style and preferences of each individual were also taken into account when administering the [primary outcome measure]." | Not reported. | No: Communication style and preference were taken into account when administering the YBOCS symptom checklist for OCD symptoms, suggesting that the potential for masking behaviours to be seen as a positive outcome was minimised. |
| Russell et al. (2020) | Yes: feedback of two autistic adults informed two iterations of the design of the intervention session materials during the development phase. No other mention of involvement of people with lived experience in the design, conduct or writing-up of the study. | Not applicable. | None reported. | Yes: The study reported that Beck Depression Inventory-II (BDI-II) and OCI-R has been found to have good psychometric properties in a sample of autistic people. Authors also noted measures that to date had no evidence of psychometric properties being investigated within autistic population: Patient Health Questionnaire-9 (PHQ-9), GRID-Hamilton Rating Scale for Depression, Generalised Anxiety Disorder Assessment (GAD-7), Positive and Negative Affect Schedule (PANAS), Work and Social Adjustment Scale (WSAS), EQ-5D-5L, 12-Item Short Form Health Survey (SF-12). | No: none of the elements of intervention or outcome measures are likely to have focus on masking or changing autistic behaviours. |
| Sizoo et al. (2017) | Yes: study reported close collaboration with autistic adults in the review of both intervention protocols, particularly around improving the explanation of aspects of autism. | Not applicable. | None reported. | No: The study reported that, to their knowledge, psychometric properties of The Hospital Anxiety and Depression Scale (HADS) have not been investigated for autistic adults, although the instrument is routinely used in clinical practice for this population. Authors also reported that the Global Mood Scale (GMS) and Rumination Reflection Questionnaire (RRQ) measures have previously been used with autistic adults, but no indication of their psychometric properties for autistic individuals was given. | Yes: the study measured positive outcomes by reduction in anxiety and depressive symptoms, negative affect, as well as autistic symptoms and rumination. Autistic people may have felt they had to mask their autistic behaviours and to change their natural way of thinking as part of the intervention. |
| Spain et al. (2017) | None reported | Not applicable. | None reported. | No: the study reports that measures commonly administered in adult non-clinical and clinical populations, including autistic samples were chosen, and that application of normative Liebowitz Social Anxiety Scale (LSAS) clinically significant thresholds to autistic people requires further scrutiny. Psychometric properties of all relevant outcome measures for autistic individuals were not reported. | Unclear: There are elements which may focus on getting autistic people to mask due to the social skills component of the program. There was a lack of detail around what suggestions were made around social skills and whether these included things such as increasing eye contact or reducing stimming. The outcome measures included measures of reduced social avoidance which may indicate increased masking behaviours. However, that most autistic participants indicated that they would like more friends, it may be that this was indeed a reasonable positive outcome measure. |
| Spek et al. (2013) | None reported | Not applicable. | None reported. | Not reported. | No: The intervention focused on mindfulness rather than changing behaviours. All the outcome measures were self-report and did not specifically measure behaviours that would indicate masking. |
| Tchanturia et al. (2021) | Yes: the study reported that the clinical pathway for autistic patients with co-occurring eating disorder was co-produced with people with lived experience. No other mention of involvement of people with lived experience in the design, conduct or writing-up of the study. | Not applicable. | None reported. | Not reported | No: None of the outcome measures are likely to have measured masking or changing autistic behaviours. |
| Wijker et al. (2020) | None reported | Not applicable. | None reported. | Not reported. | Yes: The intervention measured Social Responsiveness, using the Social Responsiveness Scale for Adults (SRS-A). This scale includes a subscale for 'restrictive and repetitive behaviour' which is a core autistic trait and also covers eye contact and social communication. Interestingly only the proxy report showed a change in social responsiveness following the intervention. The paper suggests this is due to lack of self-awareness on the part of the autistic participants, however it could also be because this perceived change was down to masking. The focus of the therapy on improving social responsiveness raises the question whether the reduction in perceived stress was due to having accommodated the communication needs of the autistic people in their lives and therefore not having to deal with miscommunications as often. |
| Wise et al. (2019) | None reported | Not applicable. | None reported. | Not reported. | Unclear: It is not clear exactly what the CBT protocol entailed specifically or how much of the exposure work was lead by the autistic persons wants. Therefore, it is possible that some of the exposure work required the autistic person to learn to mask and that some of the thought challenging was invalidating the autistic person's experiences. Interestingly, the clinician rated scales showed a significant reduction in anxiety while the self-rated scales did not. This was explained with 'lack of insight' on the part of autistic participants, rather than considering the effects of the autistic people masking traits. |

*Note.* **ECHO** = Extension for community healthcare outcomes, **EMDR** = Eye movement desensitisation and reprocessing, **PTSD** = post-traumatic stress disorder

**Table S9.** All service-level and intervention-level adaptations (detailed version) (*N* = 23)

| **Top-level categories** | **Sub-categories** | **Summary** | ***N* studies** | **Setting** | **Adaptations** | **Type of intervention** | **Rationale** |
| --- | --- | --- | --- | --- | --- | --- | --- |
| Increase knowledge and detection of autism | Clinician training and skills | Training to administer measures, tailor treatment to individual needs and increase self-efficacy, knowledge of autism and skills. Use of skills such as normalising experiences and prioritising therapeutic relationship | 5 (CYP and adults N = 4, no age information N = 1) | Inpatient, outpatient and community mental health services, autism, psychological therapies, forensic, disability and tertiary services. | Development of the Extension for Community Healthcare Outcomes (Project ECHO) Autism model (Dreiling et al. 2022) | - | To increase mental health provider self-efficacy, knowledge of autism, and problem-solving skills |
|  |  |  |  |  | Development of the Autism Intellectual Disability and Psychiatry Disorder (AUP) network (Helverschou et al., 2021) | - | To improve professional competence and the quality of specialized mental health services for individuals with autism, intellectual disability, and psychiatric disorders |
|  |  |  |  |  | Development of the Pathway for Eating disorders and Autism developed from Clinical Experience (PEACE pathway) (Tchanturia et al., 2021) | - | To increase autism awareness in eating disorder clinics, train to administer measures and tailor treatment to individual needs |
|  |  |  |  |  | Normalise experiences (Fisher et al., 2023 | EMDR | General rationale: to address or accommodate barriers |
|  |  |  |  |  | Prioritise the therapeutic relationship above everything else (Fisher et al., 2023) | EMDR | General rationale: to address or accommodate barriers |
|  |  |  |  |  | Assure you are well tuned in to the client before starting (Fisher et al., 2023) | EMDR | General rationale: to address or accommodate barriers |
|  |  |  |  |  | Be open to learning from the client and celebrate each person's uniqueness (Fisher et al., 2023) | EMDR | General rationale: to address or accommodate barriers |
|  |  |  |  |  | Be aware of the possibility of sensory overload (Fisher et al., 2023) | EMDR | General rationale: to address or accommodate barriers |
|  |  |  |  |  | Maintain the specialist skillset of staff (having skilled or trained staff including staff being skilled to communicate clearly and understand the needs of people with autism) (Petty et al., 2021) | - | Not reported |
|  |  |  |  |  | Maintain awareness of gender differences (being aware that autism can present in many ways; female presentations of autism and masking, with elaborations suggesting this is not explicitly tied to client sex) (Petty et al., 2021) | - | Not reported |
|  |  |  |  |  | Maintain awareness of gendered socialisation (being aware that people are socialised in different ways regarding gender; that gender can influence one’s life experiences) (Petty et al., 2021) | - | Not reported |
|  |  |  |  |  | Know how someone identifies (knowing, asking or checking the gender someone identifies with, including awareness of people identifying in many ways in terms of gender) (Petty et al., 2021) | - | Not reported |
|  |  |  |  |  | Do not make assumptions (not making assumptions, having no expectations or being open-minded around gender, gender identity, sexuality or ways of addressing clients) (Petty et al., 2021) | - | Not reported |
|  |  |  |  |  | Respond to the person in front of you. Some may take well to rating scales and questions about cognitions, others may not at all (Fisher et al., 2023) | EMDR | General rationale: to address or accommodate barriers |
|  | Introduction of screening tools for the detection of autism | Use of assessments such as the ASD-DF, the AQ and the RAADS-R | 3 (Adults N = 2, CYP and adults N = 1) | Inpatient, outpatient and community autism and mental health services. | Use of ASD-DF (Harrison et al. 2020) | - | To aid in the identification of individuals in need of specialized autism assessment |
|  |  |  |  |  | Use of self-report questionnaires, the AQ and the RAADS-R (Brugha et al. 2020) | - | To facilitate referral to ASD services for diagnostic assessment |
|  |  |  |  |  | Assessment of autism (Jones et al., 2021) | - | Not reported |
| Environmental adjustments | Provide environmental and practical adjustments | Provide adjustments to minimise sensory distractions such as offering sessions at the same time and place, low stimulus area and adjustments to noise, decor, odor, lighting and meals. | 4 (CYP and adults N = 3, no age information N = 1) | Inpatient and community mental health services, autism, psychological therapies, forensic, disability and tertiary services. | Change the environment (e.g., reduce bright lights or distracting noises, provide fiddle toys) (Fisher et al., 2023) | EMDR | To reduce sensory demands |
|  |  |  |  |  | Open-access low-stimulus area (Jones et al., 2021) | - | Not reported |
|  |  |  |  |  | Low-stimulus area on request (Jones et al., 2021) | - | Not reported |
|  |  |  |  |  | Scheduled access low-stimulus area (Jones et al., 2021) | - | Not reported |
|  |  |  |  |  | Lighting adaptations (Jones et al., 2021) | - | Not reported |
|  |  |  |  |  | Redecorating the ward to create a neutral color scheme (Tchanturia et al., 2021) | - | To create a more ASC-friendly ward environment |
|  |  |  |  |  | Check the suitability of the sensory environment (considering environment suitability; included checking the sensory environment and removing irritable or overwhelming things where possible) (Petty et al., 2021) | - | Not reported |
|  |  |  |  |  | Check suitability of lighting (considering lighting suitability, brightness or harshness) (Petty et al., 2021) | - | Not reported |
|  |  |  |  |  | Provide a sensory friendly environment (reducing sensory input to reduce overwhelm) (Petty et al., 2021) | - | Not reported |
|  |  |  |  |  | Reduce noise (included choosing quieter rooms, avoiding flapping blinds or tapping a pen) (Petty et al., 2021) | - | Not reported |
|  |  |  |  |  | Provide adjustable lighting (included trying to offer natural lighting or a range of lamp/lighting options) (Petty et al., 2021) | - | Not reported |
|  |  |  |  |  | Reduce scents (cooking or food smells were considered; strong disinfectants or air fresheners were avoided) (Petty et al., 2021) | - | Not reported |
|  |  |  |  |  | Neutralise decor (a neutral colour scheme avoided bright colours for the walls, furniture, carpets, clothing or accessories) (Petty et al., 2021) | - | Not reported |
|  |  |  |  |  | Reduce the number of items in the environment (practitioners maintained a minimal amount of objects within each room) (Petty et al., 2021) | - | Not reported |
|  |  |  |  |  | Avoid patterns (patterns in the environment, including on walls, carpets or clothes, were avoided) (Petty et al., 2021) | - | Not reported |
|  |  |  |  |  | Control outside noise (lawn mowers, traffic, maintenance works, simultaneous appointments and open windows were each described as noises to manage) (Petty et al., 2021) | - | Not reported |
|  |  |  |  |  | Offer space (choice was given to clients where possible of the size and layout of the room, especially for psychological therapy sessions; choice was given on seating arrangement) (Petty et al., 2021) | - | Not reported |
|  |  |  |  |  | Neutralise all sensory demands (a plain and neutral sensory environment was described, including reflecting on possible sensory demands) (Petty et al., 2021) | - | Not reported |
|  |  |  |  |  | Keep to plain design (keeping things plain or neutral included minimal decoration and uncluttered rooms or walls) (Petty et al., 2021) | - | Not reported |
|  |  |  |  |  | Ensure suitable noise levels (controlling noise included minimising sounds outside the building, using quieter rooms, having quiet waiting rooms or minimising noise from phones, clocks or equipment inside the building) (Petty et al., 2021) | - | Not reported |
|  |  |  |  |  | Consider the room seating arrangement (considering room seating arrangements, including where to sit, letting the client choose their seat or checking they are comfortable with seating closeness) (Petty et al., 2021) | - | Not reported |
|  |  |  |  |  | Utilise a protected building or space (having a designated building or space for autism services ensured design and environment decisions could be maintained) (Petty et al., 2021) | - | Not reported |
|  |  |  |  |  | Use signs up to modify the environment (signs encouraged clients to adjusts their environment, for example to shut the blinds or turn music off) (Petty et al., 2021) | - | Not reported |
|  |  |  |  |  | Ability to adapt meal plans to sensory requirements (Jones et al., 2021) | - | Not reported |
|  |  |  |  |  | Always offer sessions at the same time and place (Fisher et al., 2023) | EMDR | General rationale: to address or accommodate barriers |
|  | Normalise the use of sensory resources and stimming | Provide sensory resources such as ear defenders, weighted blankets, stress ball, relaxing music and sensory box. Encourage use of stimming behaviour. | 4 (CYP and adults N = 3, no age information N = 1) | Inpatient and community mental health services, psychological therapies, forensic, disability and tertiary services. | Ear defenders (Jones et al., 2021) | - | Not reported |
|  |  |  |  |  | Weighted blankets (Jones et al., 2021) | - | Not reported |
|  |  |  |  |  | Stress ball (Jones et al., 2021) | - | Not reported |
|  |  |  |  |  | Relaxing music (Jones et al., 2021) | - | Not reported |
|  |  |  |  |  | Develop a sensory box with items such as weighted blankets and sensory toys (Tchanturia et al., 2021) | - | To create a more ASC-friendly ward environment |
|  |  |  |  |  | Provide sensory resources (sensory or fidget toys were available) (Petty et al., 2021) | - | Not reported |
|  |  |  |  |  | Encourage them to use stimming behaviour as self-soothing if it works (Fisher et al., 2023) | EMDR | General rationale: to address or accommodate barriers |
| Communication accommodations | Plan in advance | Share a plan in advance, ensure the client is prepared, find out about the client in advance, use of appointment reminders, pay extra attention to planning and discuss issues. | 5 (Adults N = 3, CYP and adults N = 2) | Outpatient and community mental health services, autism, psychological therapies, forensic, disability and tertiary services. | Share a plan in advance for each session (Fisher et al., 2023) | EMDR | So that the client knows what to expect |
|  |  |  |  |  | Special attention was paid to planning the home practice program e.g., the exercises to do were always noted down by participants, and if necessary, planning issues were discussed individually (Spek et al., 2013) | MBT-AS | Accounting for impairment in executive functioning |
|  |  |  |  |  | Find out about the client in advance (finding out about the client in advance; included current wellbeing and priorities and checking notes about their gender preferences) (Petty et al., 2021) | - | Not reported |
|  |  |  |  |  | Find out about the client in advance from significant people (finding out about the client in advance from family, teachers, carers or other significant people) (Petty et al., 2021) | - | Not reported |
|  |  |  |  |  | Find out about the client in advance from case notes (finding out about the client in advance specifically by reading case notes or diagnostic reports) (Petty et al., 2021) | - | Not reported |
|  |  |  |  |  | Ensure the client is prepared for what will happen (ensuring the client is prepared for what is going to happen or what questions will be asked during the session) (Petty et al., 2021) | - | Not reported |
|  |  |  |  |  | Ensure the client is prepared about the purpose of the appointment (including descriptions about why they are there and ensuring consent) (Petty et al., 2021) | - | Not reported |
|  |  |  |  |  | Extra attention was paid to homework planning (Sizoo et al., 2017) | CBT, MBSR | Bearing in mind issues with potential executive functioning problems |
|  |  |  |  |  | Appointment reminders (Horwood et al., 2021; Russell et al., 2020) | GSH CBT | To accommodate the needs of individual participants |
|  | Clear communication | Provide clear instructions and guidance, repetition, be more directive, monitor, adapt and slow the pace of communication. | 5 (Adults N = 4, CYP and adults N = 1) | Specialist, outpatient and community mental health services, autism, psychological therapies, forensic, disability and tertiary services. | Use more directive interweaves than usual (Fisher et al., 2023) | EMDR | General rationale: to address or accommodate barriers |
|  |  |  |  |  | Give explicit permission to ask question (Fisher et al., 2023) | EMDR | General rationale: to address or accommodate barriers |
|  |  |  |  |  | Clarification of homework assignments (Pahnke et al., 2019) | ACT | Not reported |
|  |  |  |  |  | Be more directive in style (i.e., less Socratic, with fewer open-ended questions) (Fisher et al., 2023) | EMDR | General rationale: to address or accommodate barriers |
|  |  |  |  |  | Spell things out in black and white and be more directive than usual during history taking (Fisher et al., 2023) | EMDR | General rationale: to address or accommodate barriers |
|  |  |  |  |  | Use more prompts and suggestions to find positive cognition (Fisher et al., 2023) | EMDR | General rationale: to address or accommodate barriers |
|  |  |  |  |  | Asking questions about specific reactions like heightened pulse rather than open-ended questions (Flygare et al., 2020) | CBT | When participants struggled to report experiences during and after exposure exercises, therapists carefully reviewed thoughts, emotions, and physical reactions to help the patient identify OCD-relevant thoughts and emotions |
|  |  |  |  |  | Be very clear with clients what the preparation phase is about and why it is necessary (Fisher et al., 2023) | EMDR | General rationale: to address or accommodate barriers |
|  |  |  |  |  | Offer clear guidance on what to do after the session and what they might experience after the session (Fisher et al., 2023) | EMDR | General rationale: to address or accommodate barriers |
|  |  |  |  |  | Adapt communication (being aware of communication with clients meant communicating clearly informed by an understanding of autism) (Petty et al., 2021) | - | Not reported |
|  |  |  |  |  | Monitor own communication (descriptions of being aware of one’s own communication, including communication styles and skills, pragmatic communication (Petty et al., 2021) | - | Not reported |
|  |  |  |  |  | Communicate clearly (using clear, direct, firm, concrete or verbally explicit communication, language, or requests, adapting communication to client’s language profile or avoiding jargon) (Petty et al., 2021) | - | Not reported |
|  |  |  |  |  | Slow the pace of communication (communicating more slowly, including thinking about talking pace or giving clients more time to process language) (Petty et al., 2021) | - | Not reported |
|  |  |  |  |  | Be prepared to adjust communication (included having information available in different formats or being able to explain things in a different way) (Petty et al., 2021) | - | Not reported |
|  |  |  |  |  | Check for understanding (checking that the client understands or has understood) (Petty et al., 2021) | - | Not reported |
|  |  |  |  |  | Repeat their feedback to them (Fisher et al., 2023) | EMDR | To aid processing |
|  |  |  |  |  | Repetition was introduced (Sizoo et al., 2017) | CBT, MBSR | General rationale: to better explain aspects of autism |
|  |  |  |  |  | Repetition (Blainey et al., 2017) | CBT | To aid generalisation of skills learned |
|  | Use of simple and preferred language | Avoid use of metaphors, abstract language, awareness of the language, use of plain and preferred language. | 10 (Adults N = 6, CYP and adults N = 4) | Improving Access to Psychological Therapies (IAPT),  outpatient and community mental health services, autism, psychological therapies, forensic, disability and tertiary services. | Avoid metaphors in therapy (Cooper et al., 2018) | CBT | Not reported |
|  |  |  |  |  | Metaphors, words or sentences that are ambiguous or that require imagination skills were avoided (Kiep et al., 2015) | MBT-AS | General rationale: to suit the specific needs of adults with ASD |
|  |  |  |  |  | Minimise the use of abstract language/use of concrete examples (e.g., traffic light) (Flygare et al., 2020) | CBT | To communicate anxiety levels |
|  |  |  |  |  | Avoid metaphor (Fisher et al., 2023) | EMDR | General rationale: to address or accommodate barriers |
|  |  |  |  |  | Be clear in communication (communicating clearly; included descriptions of being black and white, using simple and concise language or giving salient summary points) (Petty et al., 2021) | - | Not reported |
|  |  |  |  |  | Avoid ambiguity (avoiding ambiguity or ambiguous expressions in communication, including non-literal or figurative language) (Petty et al., 2021) | - | Not reported |
|  |  |  |  |  | Avoid idioms (avoiding idioms in communication. This include giving examples of idioms with explanation of avoiding such language) (Petty et al., 2021) | - | Not reported |
|  |  |  |  |  | Use a literal description. Ask the person to explain what we would see if looking at a photo or a still of a movie (Fisher et al., 2023) | EMDR | General rationale: to address or accommodate barriers |
|  |  |  |  |  | Text clarification such as avoiding metaphors (Sizoo et al., 2017) | CBT, MBSR | Some people with ASD are inclined to literal interpretations |
|  |  |  |  |  | The use of metaphors, words or sentences that are ambiguous or that require imagination skills were avoided (Spek et al., 2013) | MBT-AS | Individuals with autism  have the tendency to  take language literally |
|  |  |  |  |  | Concrete/special interest-related analogies were used (Russell et al., 2013) | CBT | To convey psychological concepts |
|  |  |  |  |  | Use plain English more than with other clients (Cooper et al., 2018) | CBT | Not reported |
|  |  |  |  |  | Use very clear language. Do not assume that they have necessarily understood what you intended to say (Fisher et al., 2023) | EMDR | General rationale: to address or accommodate barriers |
|  |  |  |  |  | Instructions were made as clear as possible (Sizoo et al., 2017) | CBT, MBSR | General rationale: to better explain aspects of autism |
|  |  |  |  |  | The Dutch version of the standard EMDR procedure for children was used (Lobregt-van Buuren et al., 2019) | EMDR | The concrete language used in this protocol is suitable for people with ASD |
|  |  |  |  |  | Take time to understand the language they use around thoughts and emotions, and mirror this (Fisher et al., 2023) | EMDR | General rationale: to address or accommodate barriers |
|  |  |  |  |  | Use their own language to describe emotions (Fisher et al., 2023) | EMDR | General rationale: to address or accommodate barriers |
|  |  |  |  |  | Be aware of how they communicate their level of arousal through behaviour and use this information to evaluate how they are coping during sessions (Fisher et al., 2023) | EMDR | General rationale: to address or accommodate barriers |
|  |  |  |  |  | Be particularly mindful of language (Fisher et al., 2023) | EMDR | People may be very sensitive to failure and ‘getting it wrong’ |
|  |  |  |  |  | The intervention was being adapted from a version used with adolescents and young adults (Pahnke et al., 2019) | ACT | So that adaptation of examples to be recognizable to adults |
|  | Use of simple, written material and visual aids | Use of written information and external cues such as use of a whiteboard, color-coded worksheets, timers, agendas and calendars. Use of visual aids such as drawings, pictures, videos and leaflets. | 11 (Adults N = 4, CYP and adults N = 7) | Improving Access to Psychological Therapies (IAPT),  inpatient, outpatient and community mental health services, autism, psychological therapies, forensic, disability and tertiary services, academic medical centre. | Inclusion of written information and handouts (Blainey et al., 2017) | CBT | To bridge sessions |
|  |  |  |  |  | More written and visual information (Cooper et al., 2018) | CBT | Not reported |
|  |  |  |  |  | Visual guidance (Flygare et al., 2020) | CBT | Not reported |
|  |  |  |  |  | Use of color-coded worksheets (Pahnke 2019) | ACT | To facilitate and provide more structure for the participants |
|  |  |  |  |  | Use visual aids (e.g., drawing, pictures, videos) (Fisher et al., 2023) | EMDR | General rationale: to address or accommodate barriers |
|  |  |  |  |  | Create a visual timeline (Fisher et al., 2023) | EMDR | General rationale: to address or accommodate barriers |
|  |  |  |  |  | Include props (e.g., charts about feelings and an emotion wheel) (Fisher et al., 2023) | EMDR | To help them identify emotions |
|  |  |  |  |  | In some cases, response prevention was scheduled and initiated using external cues (e.g., timers, reminders and calendars) (Flygare et al., 2020) | CBT | To overcome executive dysfunctions in planning and response initiation, often seen in ASD. |
|  |  |  |  |  | Use visual or simplified version of ratings scales (Fisher et al., 2023) | EMDR | General rationale: to address or accommodate barriers |
|  |  |  |  |  | Use an image of a place rather than imaginal calm place (Fisher et al., 2023) | EMDR | General rationale: to address or accommodate barriers |
|  |  |  |  |  | Visual images were used (Russell et al., 2020)/Visual cues accompanied the written information (Horwood et al., 2021) | GSH CBT | To supplement written accounts of psychological principles/To increase accessibility |
|  |  |  |  |  | Visual tools were used (Russell et al., 2013) | CBT | To convey psychological concepts |
|  |  |  |  |  | Visual signage or orientation tools (Jones et al., 2021) | - | Not reported |
|  |  |  |  |  | Visual help/cue cards (Jones et al., 2021) | - | Not reported |
|  |  |  |  |  | Increased use of visual aids (Wise et al., 2019) | CBT | Not reported |
|  |  |  |  |  | Adapt written correspondence (written correspondence included clear and specific written communication or references to letters with information, checked by experts by experience) (Petty et al., 2021) | - | Not reported |
|  |  |  |  |  | Use agendas (utilising or giving the option of an agenda, sometimes specified as a written or visual agenda, or using visual strategies to explain session structure) (Petty et al., 2021) | - | Not reported |
|  |  |  |  |  | Make information available for clients about the service (available information included leaflets, information packs or documents about the service and what to expect from a visit, photographs showing clients where they are going or who they will meet) (Petty et al., 2021) | - | Not reported |
|  |  |  |  |  | Mentalise and materialize the mental states and pictures on the whiteboard while speaking (Ekman et al., 2015) | CBT | To illustrate and systematize for the client |
|  | Provide communication support | Host wellbeing groups and use of communication passports and social stories. | 2 (CYP and adults N = 1, no age information N = 1) | Inpatient mental health services. | Hosting wellbeing groups for autistic patients and for those without autistic traits together with members of the multi-disciplinary eating disorder clinic team (Tchanturia et al., 2021) | - | To support sensory difficulties and enhance social communication |
|  |  |  |  |  | Communication passports (Jones et al., 2021) | - | Not reported |
|  |  |  |  |  | Social stories (Jones et al., 2021) | - | Not reported |
| Accommodate individual differences | Evaluate individual needs and preferences | Evaluate preferences, sensitivities, needs, likes and dislikes, coping strategies and daily habits. | 5 (Adults N = 2, CYP and adults N = 3) | Inpatient, outpatient and community mental health services, autism, psychological therapies, forensic, disability and tertiary services | Assessment of likes and dislikes (Jones et al., 2021) | - | Not reported |
|  |  |  |  |  | Bespoke sensory assessment (Jones et al., 2021) | - | Not reported |
|  |  |  |  |  | Assess sensory preferences and sensitivities (Fisher et al., 2023) | EMDR | General rationale: to address or accommodate barriers |
|  |  |  |  |  | Assessment of coping strategies (Jones et al., 2021) | - | Not reported |
|  |  |  |  |  | For some, an in-depth assessment of daily habits (e.g., sleep, food intake, physical exercise and other daily routines), in the beginning of treatment (Flygare et al., 2020) | CBT | To ensure that treatment gains would not be compromised by a lack of sleep or food intake prior to exposure and response prevention |
|  |  |  |  |  | Opportunity for the coach to learn about individualised needs for autism specific adaptation (Russell et al., 2020)/Focused orientation for the coach as to how best to adapt for autistic adults on an individualised basis (Horwood et al., 2021) | GSH CBT | General rationale: tailored to meet the needs of autistic people |
|  |  |  |  |  | Find out if the client has sensory needs (understanding if the client has sensory needs) (Petty et al., 2021) | - | Not reported |
|  |  |  |  |  | Agree etiquette for making eye contact (reducing, avoiding or not expecting eye contact; thinking or asking clients about their eye contact preferences) (Petty et al., 2021) | - | Not reported |
|  |  |  |  |  | Use a preference notifications system (using a computer system where gender and associated preferences can be added as visible notifications for all staff) (Petty et al., 2021) | - | Not reported |
|  |  |  |  |  | Know pronoun or name preferences (descriptions included knowing, asking, checking or using the preferred pronouns, names or terminology) (Petty et al., 2021) | - | Not reported |
|  |  |  |  |  | Check suitability of clinician gender (ensuring a client is comfortable with the clinician’s gender; offering a chaperone or a different therapist) | - | Not reported |
|  |  |  |  |  | Presence of a standardised protocol for people with autism (specific protocol for admission, assessment and management of people with autism) (Jones et al., 2021) | - | Not reported |
|  | Encourage individual’s hobbies and interests | Include and ask about individual’s special interests and hobbies in therapy. | 4 (Adults N = 1, CYP and adults N = 3) | Improving Access to Psychological Therapies (IAPT),  outpatient and community mental health services, psychological therapies, forensic, disability and tertiary services. | Discussing individual hobbies and interests as part of therapy (Cooper et al., 2018) | CBT | Not reported |
|  |  |  |  |  | Include an individual’s special interests (Flygare et al., 2020) | CBT | To improve adherence and communication and as a reward-based behaviour |
|  |  |  |  |  | Ask about and include special interests throughout the therapy (Fisher et al., 2023) | EMDR | General rationale: to address or accommodate barriers |
|  |  |  |  |  | Use their special interests and how they feel when engaged in it as a resource (Fisher et al., 2023) | EMDR | General rationale: to address or accommodate barriers |
|  |  |  |  |  | Incorporation of specific interests into treatment (Wise et al., 2019) | CBT | General rationale: to address or accommodate barriers |
|  | Tailor practice to individual needs and preferences | Tailor care plans and practice to individual differences such as incorporating approaches targeted at neurodevelopmental comorbidities, being flexible with the treatment manual and the session timings and ensuring that resources are appropriate for the person’s gender. | 6 (Adults N = 3, CYP and adults N = 3) | Inpatient, outpatient and community mental health services, autism, psychological therapies, forensic, disability and tertiary services. | For those who presented with neurodevelopmental co-morbidities (e.g., ADHD) specific approaches targeted at these conditions were incorporated (Blainey et al., 2017) | CBT | To manage these conditions |
|  |  |  |  |  | Emotional literacy and executive function difference were supported throughout (Russell et al., 2020)/Scaffolds to support planning and scheduling of new activities were included (Horwood et al., 2021) | GSH CBT | General rationale: tailored to meet the needs of autistic people |
|  |  |  |  |  | Care plans based on individual needs specific to people with autism (Jones et al., 2021) | - | Not reported |
|  |  |  |  |  | Try different types of bilateral stimulation (e.g., eye movements, tapping, auditory sounds) (Fisher et al., 2023) | EMDR | General rationale: to address or accommodate barriers |
|  |  |  |  |  | Vary the way you work (e.g., on the floor, walking, use play, engage with their hobbies and interests) (Fisher et al., 2023) | EMDR | General rationale: to address or accommodate barriers |
|  |  |  |  |  | Offer alternatives, prompts and suggestions for cognitions (Fisher et al., 2023) | EMDR | General rationale: to address or accommodate barriers |
|  |  |  |  |  | Ask for all the elements but if they cannot provide information, go with whatever is given (Fisher et al., 2023) | EMDR | General rationale: to address or accommodate barriers |
|  |  |  |  |  | Proceed without an image if they struggle with finding an image (Fisher et al., 2023) | EMDR | General rationale: to address or accommodate barriers |
|  |  |  |  |  | Skip negative cognition altogether if it causes problems (Fisher et al., 2023) | EMDR | General rationale: to address or accommodate barriers |
|  |  |  |  |  | Use any sensory modality as a target, not necessarily an image (Fisher et al., 2023) | EMDR | General rationale: to address or accommodate barriers |
|  |  |  |  |  | Allow the positive cognition to emerge during processing rather than identifying it beforehand (Fisher et al., 2023) | EMDR | General rationale: to address or accommodate barriers |
|  |  |  |  |  | Be flexible and creative (Fisher et al., 2023) | EMDR | General rationale: to address or accommodate barriers |
|  |  |  |  |  | Don't emphasise keeping logs between sessions if difficult for the client (Fisher et al., 2023) | EMDR | General rationale: to address or accommodate barriers |
|  |  |  |  |  | Do not expect generalisation (Fisher et al., 2023) | EMDR | General rationale: to address or accommodate barriers |
|  |  |  |  |  | Use more physical movement (Fisher et al., 2023) | EMDR | General rationale: to address or accommodate barriers |
|  |  |  |  |  | Use softeners for the positive cognition (e.g., instead of I am strong, use I am starting to believe that I am strong) (Fisher et al., 2023) | EMDR | General rationale: to address or accommodate barriers |
|  |  |  |  |  | Use fantasy figures as resources (e.g., superheroes) (Fisher et al., 2023) | EMDR | General rationale: to address or accommodate barriers |
|  |  |  |  |  | Consider small traumas as well as big traumas as possible targets (Fisher et al., 2023) | EMDR | General rationale: to address or accommodate barriers |
|  |  |  |  |  | Use flash (Fisher et al., 2023) | EMDR | General rationale: to address or accommodate barriers |
|  |  |  |  |  | Use a present-day target first (Fisher et al., 2023) | EMDR | General rationale: to address or accommodate barriers |
|  |  |  |  |  | Make time for the person to debrief about their week (Fisher et al., 2023) | EMDR | General rationale: to address or accommodate barriers |
|  |  |  |  |  | Let the client choose and control length of sets (Fisher et al., 2023) | EMDR | General rationale: to address or accommodate barriers |
|  |  |  |  |  | Think in terms of a 'positive engaging focus' rather than necessarily a 'calm place' (Fisher et al., 2023) | EMDR | General rationale: to address or accommodate barriers |
|  |  |  |  |  | Expect to add to history taking throughout the therapy as new information emerges (Fisher et al., 2023) | EMDR | General rationale: to address or accommodate barriers |
|  |  |  |  |  | Be ready to reformulate throughout the therapy and to shift the focus. Whilst you might start with symptoms, later work could focus on identity, the impact of neurodiversity and adapting to diagnosis (Fisher et al., 2023) | EMDR | General rationale: to address or accommodate barriers |
|  |  |  |  |  | Keep it simple, even with things that are very complex, and adapt to the person’s level of understanding (Fisher et al., 2023) | EMDR | General rationale: to address or accommodate barriers |
|  |  |  |  |  | Be creative with the calm place (e.g., use drawings, emojis, pictures, media clips, animals, fiddle toys, smells) (Fisher et al., 2023) | EMDR | General rationale: to address or accommodate barriers |
|  |  |  |  |  | Focus first on strengths and interests and then move onto problems and history (Fisher et al., 2023) | EMDR | General rationale: to address or accommodate barriers |
|  |  |  |  |  | End with a relaxing and positive activity (Fisher et al., 2023) | EMDR | General rationale: to address or accommodate barriers |
|  |  |  |  |  | Include your own thoughts as part of the debrief (Fisher et al., 2023) | EMDR | General rationale: to address or accommodate barriers |
|  |  |  |  |  | Use their background and history to identify resource possibilities (Fisher et al., 2023) | EMDR | General rationale: to address or accommodate barriers |
|  |  |  |  |  | Focus on quality of life and functioning (Fisher et al., 2023) | EMDR | General rationale: to address or accommodate barriers |
|  |  |  |  |  | Focus on quality of life and functioning to assess progress (Fisher et al., 2023) | EMDR | General rationale: to address or accommodate barriers |
|  |  |  |  |  | Offer a flexible and individualised approach (adapting to each individual, including using techniques known to work them) (Petty et al., 2021) | - | Not reported |
|  |  |  |  |  | Adapt questioning for female representation (going beyond standard questions in an assessment) (Petty et al., 2021) | - | Not reported |
|  |  |  |  |  | Offer gender appropriate resources (ensuring that any resources used are appropriate for the gender a client identifies with) (Petty et al., 2021) | - | Not reported |
|  |  |  |  |  | Offer flexible session timings (being flexible with appointments included breaking appointments or having shorter sessions) (Petty et al., 2021) | - | Not reported |
|  |  |  |  |  | Don't insist on or encourage eye contact (Fisher et al., 2023) | EMDR | General rationale: to address or accommodate barriers |
|  |  |  |  |  | Offer your own observations of what has changed (Fisher et al., 2023) | EMDR | General rationale: to address or accommodate barriers |
|  |  |  |  |  | Flexibility around timings and frequencies of sessions (Horwood et al., 2021; Russell et al., 2020) | GSH CBT | General rationale: tailored to meet the needs of autistic people |
| Intervention structure       adaptations | Format of intervention | Reduce or increase the number and duration of sessions and exercises, additional support by therapists. | 6 (Adults N = 5, CYP and adults N = 1) | Improving Access to Psychological Therapies (IAPT), outpatient mental health services, psychological therapies services, autism services, | Increased hours of treatment (Blainey et al., 2017) | CBT | Not reported |
|  |  |  |  |  | The eight-week protocol was extended by one week (Kiep et al., 2015) | MBT-AS | Due to the relatively slow information processing |
|  |  |  |  |  | The three-minute breathing exercise was changed into a five-minute breathing exercise (Kiep et al., 2015) | MBT-AS | Due to the relatively slow information processing |
|  |  |  |  |  | Shorter or longer sessions (Cooper et al., 2018) | CBT | Not reported |
|  |  |  |  |  | The eight-week protocol was extended by one week (Spek et al., 2013) | MBT-AS | Due to the relatively slow information processing  in adults with ASD |
|  |  |  |  |  | The three-minute breathing exercise was changed into a five-minute breathing exercise (Spek et al., 2013) | MBT-AS | Due to the relatively slow information processing  in adults with ASD |
|  |  |  |  |  | Longer introductory session (Horwood et al., 2021; Russell et al., 2020) | GSH CBT | To build working alliance |
|  |  |  |  |  | Individual support on a voluntary basis from the group leaders during 30 min after each treatment group session (Pahnke et al., 2019) | ACT | Not reported |
|  | Family/caregiver/other involvement | Involve important people such as family members, partners, teachers throughout therapy. | 6 (Adults N = 2, CYP and adults N = 4) | Improving Access to Psychological Therapies (IAPT),  outpatient and community mental health services, psychological therapies, forensic, disability and tertiary services. | Involvement of family members or other carers where appropriate (Blainey et al., 2017) | CBT | Not reported |
|  |  |  |  |  | Involving a family member or partner in sessions (Cooper et al., 2018) | CBT | Not reported |
|  |  |  |  |  | Family members, teachers and other important people around participants were involved, educated, and coached (Flygare et al., 2020) | CBT | To provide optimal conditions for exposure exercises |
|  |  |  |  |  | Use storytelling, perhaps including information from others (Fisher et al., 2023) (storytelling in EMDR involves the use of caregivers) | EMDR | General rationale: to address or accommodate barriers |
|  |  |  |  |  | Parents were often encouraged to be a part of therapy when possible (Wise et al., 2019) | CBT | Parental involvement is commonly suggested as beneficial when working with youth with ASD |
|  |  |  |  |  | Obtain information from other people as well as the person themselves (Fisher et al., 2023) | EMDR | General rationale: to address or accommodate barriers |
| Intervention content      adaptations | Simplified and structured content | Remove or simplify psychoeducation and cognitive elements and ensure that there is structure. | 6 (Adults N = 3, CYP and adults N = 3) | Improving Access to Psychological Therapies (IAPT), specialist and  outpatient mental health services, autism services, research clinic embedded in clinical services. | Cognitive work (e.g., identifying and challenging negative beliefs) was simplified and used to support the behavioural components (role plays, exposure tasks and out-of-session practice tasks) that formed the core interventions in the program. (Bemmer et al., 2021) | CBT | General rationale: considered the needs of adults with SAD and co-morbid ASD who have difficulty implementing typical cognitive interventions due to limited introspection and a poorer understanding of social rules and norms |
|  |  |  |  |  | A more structured and concrete approach to therapeutic work (Cooper et al. 2018) | CBT | Not reported |
|  |  |  |  |  | The cognitive elements of the original intervention were omitted (e.g., exercises examining the content of ones thoughts were omitted) (Kiep et al., 2015) | MBT-AS | Because of deficits in information processing that characterise ASD |
|  |  |  |  |  | Sessions had a consistent structure (Russell et al., 2020; Horwood et al., 2021) | GSH CBT | General rationale: tailored to meet the needs of autistic people |
|  |  |  |  |  | The cognitive elements of the original intervention were omitted (Spek et al., 2013) | MBT-AS | Because of the information processing deficits that characterise autism |
|  |  |  |  |  | A structured and therapist-directed approach to sessional and homework content was taken (Russell et al., 2013) | CBT | Not reported |
|  |  |  |  |  | The materials had a consistent structure and format (Horwood et al., 2021; Russell et al., 2020) | GSH CBT | General rationale: tailored to meet the needs of autistic people |
|  | Taking it slow | Taking a slow/progressive approach to treatment, regular breaks. | 2 (Adults N = 1, CYP and adults N = 1) | Outpatient and community mental health services, psychological therapies, forensic, disability and tertiary services. | Slow down every phase (Fisher et al., 2023) | EMDR | General rationale: to address or accommodate barriers |
|  |  |  |  |  | Take graduated/progressive approach towards full trauma processing (Fisher et al., 2023) | EMDR | General rationale: to address or accommodate barriers |
|  |  |  |  |  | Take longer to close down and leave extra time for a debrief (Fisher et al., 2023) | EMDR | General rationale: to address or accommodate barriers |
|  |  |  |  |  | Use shorter sets and a more frequent return to target (Fisher et al., 2023) | EMDR | General rationale: to address or accommodate barriers |
|  |  |  |  |  | Use a progressive approach to processing, starting with the 'tip of the finger' (Fisher et al., 2023) | EMDR | General rationale: to address or accommodate barriers |
|  |  |  |  |  | Slower pace was set (Sizoo et al., 2017) | CBT, MBSR | General rationale: to better explain aspects of autism |
|  |  |  |  |  | Start with short sets and build up tolerance from there (Fisher et al., 2023) | EMDR | General rationale: to address or accommodate barriers |
|  | Consider the role of autism | Consider the role of autism, develop an understanding of autism such as its characteristics and impact on daily life | 2 (Adults N = 1, CYP and adults N = 1) | Outpatient and community mental health services, psychological therapies, forensic, disability and tertiary services. | Consider the role of autism within the conceptualisation (Fisher et al., 2023) | EMDR | General rationale: to address or accommodate barriers |
|  |  |  |  |  | Patients received feedback about "what is ASD?" and "What are the characteristics of my ASD?" based on assessment tests (Oshima et al., 2021) | Schema therapy | To develop an understanding of ASD |
|  |  |  |  |  | Learn what secondary disabilities of ASD are (Oshima et al., 2021) | Schema therapy | Not reported |
|  |  |  |  |  | Understand how early maladaptive schemas and schema modes created by friction between their autistic traits, and their environment are creating difficulties in their daily lives/ understand the characteristics of ASD (Oshima et al., 2021) | Schema therapy | To transform the schema modes that cause difficulties in their lives into an adaptive mode that takes into account these characteristics |
|  | Integration of emotion-focused strategies | Provide psychoeducation on emotions, arousal and feeling physiologically overwhelmed and exercises to access emotions. | 6 (Adults N = 2, CYP and adults N = 4) |  | Extended psychoeducative material (on stress) (Pahnke et al., 2019) | ACT | To help obtain knowledge of the treatment themes |
|  |  |  |  |  | Include exercises to facilitate accessing emotions (Fisher et al., 2023) | EMDR | General rationale: to address or accommodate barriers |
|  |  |  |  |  | Psychoeducation about emotions (Cooper et al., 2018) | CBT | Not reported |
|  |  |  |  |  | If required, educational sessions about understanding and rating anxiety were provided (Russell et al., 2013) | CBT | Not reported |
|  |  |  |  |  | Provide extra psychoeducation around trauma, arousal and feeling physiologically overwhelmed (Fisher et al., 2023) | EMDR | General rationale: to address or accommodate barriers |
|  |  |  |  |  | Psychoeducation on anxiety and ASD for both the adolescent and parent when involved in therapy (Wise et al., 2019) | CBT | General rationale: to address or accommodate barriers |
|  |  |  |  |  | Extra sessions devoted to psychoeducation skills training in anxiety identification and communication (Flygare et al., 2020) | CBT | To meet the needs of individuals with ASD |
|  | Integration of cognitive-behavioural approaches | Provide cognitive and behavioural strategies including building a positive self-image, coping strategies and making links between behaviour, thoughts and feelings. | 6 (Adults N = 1, CYP and adults N = 5) | Improving Access to Psychological Therapies (IAPT), specialist, outpatient and community mental health services, psychological therapies, forensic, disability and tertiary services, research clinic embedded in clinical services. | Behavioural strategies to introduce change (Cooper et al., 2018) | CBT | Not reported |
|  |  |  |  |  | Behavioural interventions were integrated within treatment sessions, and as a focus of weekly homework (Bemmer et al., 2021) | CBT | To facilitate engagement and promote positive treatment outcomes |
|  |  |  |  |  | Cognitive strategies to introduce change (Cooper et al., 2018) | CBT | Not reported |
|  |  |  |  |  | Facilitate learning about links between situations, behaviours and feelings and use of this learning to schedule activities promoting positive feelings (Russell et al., 2020)/Explicit learning tasks (Horwood et al., 2021) | GSH CBT | To ensure the foundation skills for BA were in place i.e. making links between situations, feelings and behaviours and noticing and rating positive feelings |
|  |  |  |  |  | Focus on building a positive self-image and coping strategies rather than pathologising and eliminating symptoms (Fisher et al., 2023) | EMDR | General rationale: to address or accommodate barriers |
|  |  |  |  |  | Install a positive self-view as a resource (Fisher et al., 2023) | EMDR | General rationale: to address or accommodate barriers |
|  |  |  |  |  | Do more cognitive work if necessary to identify a positive cognition (Fisher et al., 2023) | EMDR | General rationale: to address or accommodate barriers |
|  |  |  |  |  | Emphasis on a reward system (Wise et al., 2019) | CBT | To help motivate the adolescent to engage in exposure therapy |
|  |  |  |  |  | Ensuring the building blocks for treatment (i.e., understanding and differentiating emotions, particularly anxiety, and making links between thoughts, feelings and behaviours) were in place (Russell et al., 2013) | CBT | Not reported |
|  | Integration of social skills training | Integration of social skills training such as entering and maintaining conversations and managing disagreements | 3 (Adults N = 1, CYP and adults N = 2) | Research clinic embedded in clinical services, academic medical center and psychological therapies services. | Social skills training was offered in some cases, which is part of broader skills training (including emotion recognition and regulation, problem-solving approaches) (Blainey et al., 2017) | CBT | Not reported |
|  |  |  |  |  | Inclusion of structured frameworks for teaching of social skills such as entering and maintaining conversations and managing disagreements (Bemmer et al., 2021) | CBT | To make the anxiety-based interventions more effective for adults with ASD |
|  |  |  |  |  | Treatment components (e.g., independence and social skills) (Wise et al., 2019) | CBT | To address ASD-specific deficits |

*Note.* **ACT** = Acceptance and Commitment Therapy, **ASD-DF =** Autism Spectrum Disorder-Discriminant Function, **AQ =** Autism-spectrum Quotient (**CBT** = Cognitive Behavioural Therapy, **EMDR** = Eye Movement Desensitisation and Reprocessing, **GSH** = Guided Self-Help, **MBSR** = Mindfulness-Based Stress Reduction, **MBT-AS** = Mindfulness-Based Therapy for Autism Spectrum disorders, **RAADS-R** = Ritvo autism-Asperger's diagnostic scale-revised.

**Table S10.** GRADE Assessment for effectiveness outcomes

| **N studies** | **Study quality** | **Concerns about certainty** | **Inconsistency** | **Concerns about certainty** | **Indirectness** | **Concerns about certainty** | **Imprecision** | **Concerns about certainty** | **Publication bias** | **Concerns about certainty** | **Certainty** |
| --- | --- | --- | --- | --- | --- | --- | --- | --- | --- | --- | --- |
| **Adapted individual CBT for anxiety** | | | | | | | | | | | |
| 2 | 1/2 studies was of low and 1/2 was of moderate methodological quality | Borderline | 1/2 report significant improvement in self-reported anxiety and clinician-rated global functioning from pre- to post-treatment. 1/2 report significant improvement in clinician-rated anxiety and severity of psychopathology, and no significant changes in clinician-rated depression and self-reported anxiety from pre- to post-treatment. Therefore, there is minor inconsistency across the results of these studies. | Borderline | 2/2 studies used established rating scales. 2/2 measured effect of time. | Serious concerns | 2/2 had a sample size under 100 individuals. There were only two contributing study | Serious concerns | Both significant and non-significant findings were reported. There were just two contributing studies. | Borderline | low certainty ⊕⊕OO |
| **Adapted individual CBT for OCD** | | | | | | | | | | | |
| 2 | 1/2 was of moderate methodological quality, 1/2 was of high methodological quality. | No concern | 1/1 report significant improvement in clinician-rated and self-reported OCD and clinician-rated depressive symptoms from pre- to post-treatment, but no significant change in global functioning and quality of life. 1/1 report no significant difference in clinician-rated OCD symptoms between treatment groups, except significant difference between groups in proportion of participants clinician-rated as improved; and no significant differences between pre-, post-, and 1-month follow-up in the secondary mental health and social outcomes in neither treatment group, except for informant-rated OCD symptoms that significant improved over time only in one treatment group. Therefore, there is minor inconsistency across the results of these studies. | Borderline | 2/2 studies used established rating scales. 1/2 analysed group by time interaction. 2/2 measured effect of time. | Borderline | 2/2 had a sample size under 100 individuals. There were only two contributing study | Serious concerns | Both significant and non-significant findings were reported. There were just two contributing studies. | Borderline | moderate certainty ⊕⊕⊕O |
| **Adapted EMDR** | | | | | | | | | | | |
| 1 | 1/1 was of high methodological quality | No concern | Impossible to assess inconsistency as there is just one contributing study. | No concern | 1/1 study used established rating scales. Effect of time analysed, and participants acted as their own control. | Borderline | 1/1 had a sample size under 100 individuals. There was only one contributing study | Serious concerns | Only one study found, potential publication bias | Serious concerns | low certainty ⊕⊕OO |
| **Adapted Schema therapy** | | | | | | | | | | | |
| 1 | 1/1 was of high methodological quality | No concern | Impossible to assess inconsistency as there is just one contributing study. | No concern | 1/1 used established rating scales. 1/1 measured effect of time. | Serious concerns | 1/1 had a sample size under 100 individuals. There was only one contributing study | Serious concerns | Only one study found, potential publication bias | Serious concerns | very low certainty ⊕OOO |
| **Adapted individual CBT for general psychological distress** | | | | | | | | | | | |
| 1 | 1/1 was of moderate methodological quality | No concern | Impossible to assess inconsistency as there is just one contributing study. | No concern | 1/1 used established rating scales. 1/1 measured effect of time. | Serious concerns | 1/1 had a sample size under 100 individuals. There was only one contributing study | Serious concerns | Only one study found, potential publication bias | Serious concerns | very low certainty ⊕OOO |
| **Adapted group mindfulness-based therapy** | | | | | | | | | | | |
| 3 | 1/3 was of low methodological quality, 1/3 was of moderate methodological quality, 1/3 was of high methodological quality | Borderline | 1/3 reported no significant difference between treatment groups in anxiety and depression scores, global mood and rumination. 1/3 reported significant difference between two treatment groups in depressive and anxiety symptoms, positive affect and rumination. 1/3 reported significant improvement in psychological symptoms such as general psychopathology, rumination, and positive affect from pre- to post-treatment. Therefore, there is minor inconsistency across the results of these studies. | Borderline | 3/3 studies used established rating scales. 1/3 analysed group by time interaction, no randomisation. 1/3 analysed group by time interaction. 1/3 measured effect of time. | Borderline | 3/3 had a sample size under 100 individuals. There were 3 contributing studies. | Serious concerns | No publication bias is suspected, as both significant and non-significant findings were found. | No concern | moderate certainty ⊕⊕⊕O |
| **Adapted group CBT for anxiety and depression** | | | | | | | | | | | |
| 1 | 1/1 of low methodological quality | Serious concerns | Impossible to assess inconsistency as there is just one contributing study. | No concern | 1/1 study used established rating scales. Group by time interaction was analysed, but no randomisation was done | Borderline | 1/1 had a sample size under 100 individuals. There was only one contributing study | Serious concerns | Only one study found, potential publication bias | Serious concerns | very low certainty ⊕OOO |
| **Adapted group CBT for social anxiety** | | | | | | | | | | | |
| 1 | 1/1 of high methodological quality | No concern | Impossible to assess inconsistency as there is just one contributing study. | No concern | 1/1 used established rating scales. 1/1 measured effect of time. | Serious concerns | 1/1 had a sample size under 100 individuals. There was only one contributing study | Serious concerns | Only one study found, potential publication bias | Serious concerns | very low certainty ⊕OOO |
| **Adapted group acceptance and commitment therapy** | | | | | | | | | | | |
| 1 | 1/1 of high methodological quality | No concern | Impossible to assess inconsistency as there is just one contributing study. | No concern | 1/1 used established rating scales. 1/1 measured effect of time. | Serious concerns | 1/1 had a sample size under 100 individuals. There was only one contributing study | Serious concerns | Only one study found, potential publication bias | Serious concerns | very low certainty ⊕OOO |
| **PEACE Pathway** | | | | | | | | | | | |
| 1 | 1/1 of high methodological quality | No concern | Impossible to assess inconsistency as there is just one contributing study. | No concern | 1/1 study used established data collection methods to assess service use, i.e., data on hospital admissions from clinical records. However, no statistical analysis was reported, and no randomisation was done. | Borderline | Sample size of individuals was not reported. | Serious concerns | Only one study found, potential publication bias | Serious concerns | low certainty ⊕⊕OO |
| **AUP network** | | | | | | | | | | | |
| 1 | 1/1 of moderate methodological quality | No concern | Impossible to assess inconsistency as there is just one contributing study. | No concern | 1/1 used established rating scales. 1/1 measured effect of time. | Serious concerns | 1/1 had a sample size above 100 individuals. There was only one contributing study | Serious concerns | Only one study found, potential publication bias | Serious concerns | very low certainty ⊕OOO |
| **Project ECHO** | | | | | | | | | | | |
| 1 | 1/1 of low methodological quality | Serious concerns | Impossible to assess inconsistency as there is just one contributing study. | No concern | 1/1 measured effect of time and used less direct proxies for effectiveness | Serious concerns | 1/1 had a sample size under 100 individuals. There was only one contributing study | Serious concerns | Only one study found, potential publication bias | Serious concerns | very low certainty ⊕OOO |
| **Detection of autism strategies** | | | | | | | | | | | |
| 2 | 2/2 studies were of high methodological quality | No concern | 2/2 studies report on tools that can identify individuals for whom specialised autism assessment is needed, thus the results are sufficiently consistent. | No concern | 2/2 studies used established rating scales to assess identification of autism | No concern | 2/2 studies had a sample size above 100 individuals. There were just two contributing studies | Borderline | Only significant findings were reported. There are just two contributing studies. | Serious concerns | moderate certainty ⊕⊕⊕O |
| **Bespoke individual CBT for anxiety with VR** | | | | | | | | | | | |
| 1 | 1/1 of moderate methodological quality | No concern | Impossible to assess inconsistency as there is just one contributing study. | No concern | 1/1 used established rating scales. 1/1 measured effect of time. | Serious concerns | 1/1 had a sample size under 100 individuals. There was only one contributing study | Serious concerns | Only one study found, potential publication bias | Serious concerns | very low certainty ⊕OOO |
| **Bespoke individual Animal-assisted therapy** | | | | | | | | | | | |
| 1 | 1/1 of high methodological quality | No concern | Impossible to assess inconsistency as there is just one contributing study. | No concern | 1/1 studies used established rating scales. 1/1 analysed group by time interaction. | No concern | 1/1 had a sample size under 100 individuals. There was only one contributing study | Serious concerns | Only one study found, potential publication bias | Serious concerns | low certainty ⊕⊕OO |
| **Bespoke group CBT for social anxiety** | | | | | | | | | | | |
| 1 | 1/1 of high methodological quality | No concern | Impossible to assess inconsistency as there is just one contributing study. | No concern | 1/1 used established rating scales. 1/1 measured effect of time. | Serious concerns | 1/1 had a sample size under 100 individuals. There was only one contributing study | Serious concerns | Only one study found, potential publication bias | Serious concerns | very low certainty ⊕OOO |
| **Bespoke group CBT for anxiety** | | | | | | | | | | | |
| 1 | 1/1 of high methodological quality | No concern | Impossible to assess inconsistency as there is just one contributing study. | No concern | 1/1 studies used established rating scales. 1/1 analysed group by time interaction. | No concern | 1/1 had a sample size under 100 individuals. There was only one contributing study | Serious concerns | Only one study found, potential publication bias | Serious concerns | low certainty ⊕⊕OO |
| **Bespoke group CBT for anxiety, stress and depression** | | | | | | | | | | | |
| 1 | 1/1 of high methodological quality | No concern | Impossible to assess inconsistency as there is just one contributing study. | No concern | 1/1 study used established rating scales. Group by time interaction was analysed, but no randomisation was done | Borderline | 1/1 had a sample size under 100 individuals. There was only one contributing study | Serious concerns | Only one study found, potential publication bias | Serious concerns | low certainty ⊕⊕OO |
| **Bespoke individual RTSM using a mobile platform** | | | | | | | | | | | |
| 1 | 1/1 of low methodological quality | Serious concerns | Impossible to assess inconsistency as there is just one contributing study. | No concern | 1/1 used established rating scales. 1/1 measured effect of time. | Serious concerns | 1/1 had a sample size under 100 individuals. There was only one contributing study | Serious concerns | Only one study found, potential publication bias | Serious concerns | very low certainty ⊕OOO |

**Method:** The Grading of Recommendations Assessment, Development and Evaluation (GRADE) system (Guyatt et al., 2008), adapted for narrative synthesis according to (Murad et al., 2017) and according to methodological aspects of research contributing to the research question about effectiveness of strategies to improve mental health care for autistic people. The certainty of evidence for each outcome was independently assessed by two people, after which they met to address any inconsistencies. Each GRADE domain could obtain ‘no concerns’, ‘borderline’ or ‘serious concerns’ rating and the overall certainty for each outcome started as high and was lowered for any ‘serious concerns’. The GRADE domains were rated accordingly:

- **Study quality** – ‘no concerns’ were noted if ≤ 33% of the contributing studies were of low quality, ‘borderline’ if 34-67% were of low quality, and ‘serious concerns’ if > 67% were of low quality based on quality ratings (see Table S7).
- **Inconsistency** – consistency of the direction of change and the magnitude of effects across the research evidence was evaluated. ‘No concerns’ were noted when most studies reported associations/effects in the same direction, or where there was only one contributing study and therefore the inconsistency was impossible to tell. ‘Serious concerns’ were noted when there was evidence of opposite directions of change (e.g., significant improvement and significant worsening), and ‘borderline’ concerns were notes when there was evidence of significant improvement/worsening and no significant change.
- **Indirectness** – a judgement was made on the degree of similarity of the research evidence with the research question of interest, reflecting on how directly the available evidence answered the specific research questions set out in the review. For example, measures of effect of time instead of group by time interactions and lack of randomisation contributed to down-ratings for this domain.
- **Imprecision** – a judgement was made based on the total number of contributing studies and their sample size. The sample size threshold used for the relevant analysis was 100. ‘Serious concerns’ were noted when there was only one contributing study, even if its sample size was above 100 individuals.
- **Publication bias** – we considered if studies contributing to an outcome reported significant and non-significant results, or if publication bias was likely due to missing evidence. ‘Serious concerns’ were assigned if there were only one contributing study per outcome, as this may have indicated a shift in research publication priorities.

**Table S11.** Full results by study

| **Authors** | **Strategy vs comparison** | **Outcomes/measures** | **Adaptation categories and sub-categories** | **Acceptability/Feasibility findings** | **Effectiveness findings** |
| --- | --- | --- | --- | --- | --- |
| Bemmer et al. (2021) | Adapted CBT for social anxiety | **Primary outcome measures**  Liebowitz Social Anxiety Scale (LSAS-SR) - self report measure of anxiety and avoidance of social situation.  **Secondary outcome measures**  Depression Anxiety Stress Scale (DASS-21) - self-report measure of symptom severity of depression, anxiety and stress.  Kessler Psychological Distress Scale (K10) - self-report measure of psychological distress.  Social Interaction Anxiety Scale (SIAS)- self-report measure of social anxiety.  Social Phobia Scale (SPS) - self-report measure of social anxiety.  Tolerability measures - self-report at the mid-point of treatment, assessing expectations of, and engagement with the intervention, and potential barriers. It consisted of six free response, and two Likert-scale questions. | **Intervention content**  - Simplified and structured content  - Integration of cognitive-behavioural approaches  - Integration of social skills training | **Feasibility**  - *Drop-out rate:* 6/84 participants dropped out (8%).  **Acceptability**  - *Experience of care at mid-point (quantitative):* 96% of 28 participants agreed or strongly agreed that they are enjoying the group.  - *Experience of care at mid-point (qualitative):* Participants indicated that making friends, being able to talk and ask questions without judgement, feeling understood by others and having practical help and support were working well within the group. Participants indicated that making phone calls, feeling like their anxiety was hindering their learning, and finding the groups either too long or too short were difficulties with the group. Participants generally reported positive engagement with other group members, though some reported difficulties with a group member who was perceived as too talkative and disruptive. | **Effect of time (no comparison group)**  **Primary outcomes**  Mental health outcome using the LSAS-SR:  - Significant improvements in anxiety and avoidance of social situations (total score) from pre (M = 79.78, SD = 27.36), to post (M = 70.17, SD = 31.04), M change = 9.61 (SD = 20.41), p < .001.  **Secondary outcomes**  Mental health outcomes using DASS-21; K10; SIAS; & SPS:  - Significant improvements in social anxiety related to initiating and maintaining conversations, depression, anxiety and stress from pre- to post-intervention, but not in social anxiety related to fears of being observed or evaluated in daily activities and psychological distress. |
| Blainey et al. (2017) | Adapted CBT | **Primary and secondary outcome measures not specified.**  Clinical Outcomes in Routine Evaluation-Outcome (CORE-OM) - self-report measure of global distress. | **Communication accommodations**  - Clear communication  - Use of simple, written material and visual aids  **Accommodate individual differences**  - Tailor practice to individual needs and preferences  **Intervention structure**  - Format of intervention  - Family/caregiver/other involvement  **Intervention content**  - Integration of social skills training | **Feasibility**  **-** *Intervention attendance rate:* 80% (n=62) of individuals completed all sessions of therapy that were offered. All participants attended ≥ 3 sessions. | **Effect of time (no comparison group)**  Mental health outcome using CORE-OM:  - Significant improvements in *global distress* (total) from pre (M = 1.79, SD = 0.79), to post (M = 1.48, SD = 0.79), M change = -0.31, p < .001 CI [0.17, 0.44], d = 0.39 (small effect size)  - 36.9% reliably improved (reliable change defined as an increase or decrease in clinical score of 5.00 or more), 38.5% did not experience a large enough change to be classified as reliable and 24.6% deteriorated from first to last session. 18.5% of the individuals who met criteria for caseness on the CORE‐OM at first session experienced clinical change (i.e., clinical to non-clinical) in global distress post-therapy. |
| Brugha et al. (2020) | Detection of autism | The autism-spectrum quotient (AQ).  Ritvo autism–Asperger's diagnostic scale-revised (RAADS-R).  A subsample was selected for a second assessment with the autism diagnostic observation schedule (ADOS Module 4). | **Increase knowledge and detection of autism**  - Introduction of screening tools for the detection of autism | Not applicable. | **Effectiveness in detecting autism in mental health services**  Autism detection using AQ; & RAADS-R:  - "Fair" diagnostic accuracy was observed for the AQ, based on the higher cut-off threshold for autism (ADOS threshold 10+). Optimal sensitivity and specificity was at a cut-off of 31, with sensitivity 0.79 (95% CI [0.54, 0.94]) and specificity 0.77 (95% [0.65, 0.86]).  - "Fair" diagnostic accuracy was observed for the RAADS-R, based on the higher cut-off threshold for autism (ADOS threshold 10+). Optimal sensitivity and specificity was at a cut-off of 120–126, with sensitivity 0.75 (95% CI [0.48, 0.93]) and specificity 0.71 (95% CI [0.60, 0.81]). |
| Cooper at al. (2018) | Not applicable. | **Primary and secondary outcomes not specified.**  Short survey developed for this study, which sought information such as knowledge/use of adaptations of CBT as outlined in the National Institute for Health and Care Excellence (NICE) guidance. | **Communication accommodations**  - Use of simple and preferred language  - Use of simple, written material and visual aids  **Accommodate individual differences**  - Encourage individual's hobbies and interests  **Intervention structure**  - Format of intervention  - Family/caregiver/other involvement  **Intervention content**  - Simplified and structured content  - Integration of emotion-focused strategies  - Integration of cognitive-behavioural approaches | **Feasibility**  *% of therapists endorsing the following adaptations:*  - Behavioural strategies to introduce change (n = 37, 74%); Using plain English more than with other clients (n = 35, 70%); A more structured and concrete approach to therapeutic work (n = 35, 70%); Psychoeducation about emotions (n = 34, 68%); More written and visual information than I usually use (n = 30, 60%); Discussing individual hobbies and interests as part of therapy (n = 29, 58%); Involving a family member or partner in sessions (n = 24, 48%); Avoiding metaphors in therapy (n = 20, 40%); Shorter sessions (n = 14, 28%); Cognitive strategies to introduce change (n = 14, 28%); Other (n = 5, 10%); Longer sessions (n = 1, 2%)  **Acceptability**  - *Satisfaction with care:* Most respondents favoured a cognitive behavioural approach with autistic clients, rating it 7.17/10 for helpfulness, and 74% having ever used this approach. 16% of responders used systemic approaches, giving it an average rating of 6.75/10 for helpfulness. 14% of respondents used eclectic approaches, giving it an average of 6.29/10 for helpfulness. 4% of responders used psychodynamic approaches giving it a 3.1/10 rating for helpfulness. | Not applicable. |
| Dreiling et al. (2022) | Extension for community healthcare outcomes project (Project ECHO) | **Primary and secondary outcome measures not specified.**  Primary Care Autism Self-Efficacy (PCASE) Survey - self-report measuring mental health provider level of confidence in effectively enacting the treatment strategy described.  Knowledge test - self-report measure of mental health provider autism knowledge.  Problem-solving scenarios - self-report measure to assess changes in provider’s clinical problem-solving skills using evidence-based strategies that were discussed during ECHO Autism sessions.  Satisfaction survey - self-report. Participants were also invited to share their thoughts and suggestions for improvement in free-response sections. | **Increase knowledge and detection of autism**  - Clinician training and skills | **Feasibility**  - *Attendance rate:* 88.2% (n = 45) of participants attended at least 80% of ECHO sessions (average attendance per provider was 8.78/10 sessions). Providers who attended less than 60% of ECHO sessions (n=4), who attended more than 60% of sessions but failed to complete post-assessment measures (n=1), or who failed to meet both attendance and pre-post questionnaire completion (n=30) were excluded from the analysis, yielding a final sample size of 51.  **Acceptability**  - *Satisfaction (quantitative)* (M = 1.32, range 1-2) was rated highly (5-point scale, with “1” indicating the highest degree of satisfaction).  - *Satisfaction (qualitative):* responses were positive, suggestions for improvement included ideas for additional didactic topics (e.g., gender and ASD) and recommendations for future ECHO Autism cohorts made up of previous participants to gain advanced-level training including additional opportunities for case presentations and feedback. | **Effect of time (no comparison group).**  **Outcomes on the way to improving care:**  Clinicians’ self-efficacy using PCASE Survey; autism knowledge using the Knowledge test; problem-solving skills using Problem-solving scenarios:  - Significant improvements in self-efficacy from pre (M = 64.90, SD = 13.36), to post (M = 85.29, SD = 11.10), p < 0.001.  - Significant improvements in autism knowledge from pre (M = 11.06, SD = 2.77), to post (M = 14.31, SD = 2.56), p < 0.001.  - Significant improvements in awareness in best-practice treatment considerations for autistic individuals from pre (M = 0.92, SD = 0.23), to post (M = 1.34, SD = 0.18), p < 0.001. |
| Ekman et al. (2015) | Adapted CBT for anxiety | **Primary and secondary outcome measures not specified.**  Level of anxiety - self-report measured on a scale from 0-3 for anxiety.  Global Function Rating Scale (GAF scale) - clinician-rated. | **Communication accommodations**  - Use of simple, written material and visual aids | **Acceptability**  - *Satisfaction with care:* Most clients reported finding the visualisation helpful. Help from visualisation to remember the conversation with the therapist: Yes: 14/18; Not known: 1/18; No: 3/18; Chi square test p value = 0.001. Visualisation useful in homework: Yes: 13/18; Not known: 2/18; No: 3/18; Chi square test p value = 0.005. | **Effect of time (no comparison group).**  Mental health outcome using Level of anxiety:  **-** Anxiety related to behaviour excess: a statistically significant level of symptom reduction (p<0.001). In post hoc tests (Bonferroni) this was explained by a significant decrease in anxiety from both pre-measurements to post-treatment (p<0.05).  **-** Anxiety related to behavioural avoidance: a statistically significant level of symptom reduction (p<0.0001). In post hoc tests (Bonferroni), this was explained by a significant decrease in anxiety from both pre-measurements to post-treatment (p<0.005).  **-** Anxiety related to cognitive excess: a significant main term (p<0.001). In post-hoc tests (Bonferroni), this was explained by a significant decrease in anxiety from both pre-measurements to midpoint (p<0.05) and post-treatment (p<0.05).  **-** Anxiety related to cognitive avoidance: no significant differences.  Social outcomes using GAF:  - Significant improvement in clients’ psychological, social and occupational ability to function from pre-treatment (Mean = 55.72, SE = 2.19) to post-treatment (Mean= 73.17, SE = 2.71), p < 0.0001. |
| Fisher at al. (2022) | Adapted EMDR | **Primary and secondary outcome measures not specified.**  Round 2 survey comprised of statements generated by thematically analysing responses to Round 1. Statements rated on a 5-point scale (1 = I always do this, 2 = I often do this, 3 = I sometimes do this, depending on the client, 4 = I never do this, 5 = I think this should not be done at all). | **Increase knowledge and detection of autism**  - Clinician training and skills  **Environmental adjustments**  - Provide environmental and practical adjustments  - Normalise the use of sensory resources and stimming  **Communication accommodations**  - Plan in advance  - Clear communication  - Use of simple and preferred language  - Use of simple, written material and visual aids  **Accommodate individual differences**  - Evaluate individual needs and preferences  - Encourage individual's hobbies and interests  - Tailor practice to individual needs and preferences  **Intervention structure**  - Family/caregiver/other involvement  **Intervention content**  - Taking it slow  - Consider the role of autism  - Integration of emotion-focused strategies  - Integration of cognitive-behavioural approaches | **Feasibility**  *Elements of EMDR that ≥ 80% of therapists often or always incorporate in therapy with autistic clients*: 88% normalise experiences, 86% use very clear language, 91% be flexible and creative, 84% be aware of how they communicate their level of arousal through behaviour and use this information to evaluate how they are coping during sessions, 91% take time to understand the language they use around thoughts and emotions, and mirror this, 91% respond to the person in front of you, 95% be open to learning from the client and celebrate each person's uniqueness, 86% consider the role of autism within the conceptualisation, 98% consider non-life threatening traumas and life threatening traumas as possible targets during history taking, 88% use their background and history to identify resource possibilities, 84% try a range of different types of bilateral stimulation, 80% be particularly mindful of language in the re-evaluation phase as people may be very sensitive to failure and 'getting it wrong', 83% use their own language to describe emotions, 80% make time for the person to debrief about their week.  *Elements of EMDR that ≥ 80% of therapists sometimes incorporate in therapy:* 93% assess sensory preferences and sensitivities, 100% be more directive in style, 86% use visual aids, 93% avoid metaphors, 98% ask about and include special interests throughout, 100% always offer sessions at the same time and place, 100% take graduated/ progressive approach towards full trauma processing, 81% use flash, 93% focus on quality of life and functioning, 98% share a plan in advance for each session to manage expectations, 95% change the environment to reduce sensory demands, 100% provide extra psychoeducation around trauma, arousal and feeling physiologically overwhelmed, 98% slow down every phase, 88% use storytelling, 100% focus on building a positive self-image and coping strategies rather than pathologizing and eliminating symptoms, 100% prioritise the therapeutic relationship above everything else, 95% don't insist on or encourage eye contact, 81% use visual or simplified version of ratings scales, 100% be ready to reformulate throughout the therapy and to shift the focus, 93% give explicit permission to ask questions, 86% obtain information from other people as well as the person themselves, 100% expect to add to history taking throughout the therapy as new information emerges, 93% focus first on strengths and interests and then move onto problems and history, 86% vary the way you work during history taking, 88% create a visual timeline during history taking, 95% spell things out in black and white and be more directive than usual, 98% use an image of a place rather than imaginal calm place, 95% think in terms of a 'positive engaging focus' rather than necessarily a 'calm place', 93% be creative with the calm place, 81% encourage them to use stimming behaviour as self-soothing if it works, 95% use fantasy figures as resources in preparation stage, 93% use their special interests and how they feel when engaged in it as a resource, 81% include exercises to facilitate accessing emotions in preparation stage, 81% include props to help them identify emotions , 84% install a positive self-view as a resource, 91% be very clear with clients what the preparation phase is about and why it is necessary, 100% ask for all the elements but if they cannot provide information, go with whatever is given in assessment phase, 100% assure you are well tuned in to the client before starting, 80% use a progressive approach to processing, starting with the 'tip of the finger', 88% use any sensory modality as a target, not necessarily an image, 90% use a present-day target first, 88% use a literal description. Ask the person to explain what we would see if looking at a photo or a still of a movie, 95% proceed without an image if they struggle with finding an image, 98% offer alternatives, prompts and suggestions for cognitions, 93% skip negative cognition altogether if it causes problems, 88% allow the positive cognition to emerge during processing rather than identifying it beforehand, 95% use more prompts and suggestions to find positive cognition, 93% use softeners for the positive cognition, 100% be aware of the possibility of sensory overload, 98% use more directive interweaves than usual, 98% let the client choose and control length of sets in desensitisation phase, 93% use shorter sets and a more frequent return to target, 90% do not expect generalisation, 85% repeat their feedback to them to aid processing, 80% use more physical movement, 90% start with short sets and build up tolerance from there, 85% do more cognitive work at this stage if necessary to identify a positive cognition, 98% take longer to close down and leave extra time for a debrief, 95% end with a relaxing and positive activity, 100% offer clear guidance on what to do after the session and what they might experience after the session , 100% include your own thoughts as part of the debrief, 100% focus on quality of life and functioning to assess progress, 100% offer your own observations of what has changed, 95% don't emphasise keeping logs between sessions if difficult for the client. | Not applicable. |
| Flygare et al. (2020) | Adapted CBT for OCD | **Primary outcome measures**  Yale-Brown Obsessive Compulsive Scale (Y-BOCS) - clinician-rated measure assessing severity of OCD symptoms.  **Secondary outcome measures**  Clinical Global Impression Scale for severity and improvement (CGI-S and CGI-I) - clinician-administered.  Global Assessment of Functioning (GAF) - clinician-administered.  Obsessive Compulsive Inventory-Revised (OCI-R) - self-report of OCD symptoms.  Montgomery Asberg Depression Rating Scale-Self report (MADRS-S)- self-report of depressive symptoms.  EuroQol 5-dimensions (EQ-5D) - self-report of quality of life.  Number of cancelled appointments and adherence to homework assignments. | **Communication accommodations**  - Clear communication  - Use of simple and preferred language  - Use of simple, written material and visual aids  **Accommodate individual differences**  - Evaluate individual needs and preferences  - Encourage individual's hobbies and interests  **Intervention structure**  - Family/caregiver/other involvement  **Intervention content**  - Integration of emotion-focused strategies | **Feasibility**  *- Intervention attendance rate:* Attended sessions was low (M = 15.93, SD = 2.79), range 10-19. Missed or cancelled sessions M = 4 (SD = 2.79), range 1-10 weeks. Duration of treatment was spread out (M = 33 weeks, SD = 9.47), range 22-56 weeks  - *Intervention drop-out rate*: 3 participants (1 at session 8, 2 after mid-treatment), 6 participants lost to follow-up (1 at mid-treatment, 5 at 3-month follow-up). Thus, post-treatment, 4/19 participants dropped out - thus dropout rate is low.  - *Homework compliance:* rated as good (53%), adequate (12%), or poor (35%) by the therapists. | **Effect of time (no comparison group)**  **Primary outcomes**  Mental health outcome using Y-BOCS:  - Significant improvements in clinician-rated *obsessive-compulsive symptoms* from pre (M = 24.68, SD = 1.21) to post (M = 16.96, SD = 1.28), M change = -7.72 [95% CI -2.30, -0.64], p < .001, and from pre to 3-month follow up (M = 18.43, SD = 1.48), M change = -6.25 [95% CI -2.05, -0.33], p < .001, but not from post to 3-month follow-up, p = .389.  **Secondary outcomes**  Mental health outcomes using OCI-R; MADRS-S; CGI-S; & CGI-I:  - Significant improvements in self-reported obsessive-compulsive symptoms and depressive symptoms from pre- to post-treatment. Significant improvements in obsessive-compulsive symptoms from post to 3-month follow-up, but not in depressive symptoms.  - Significant improvements in obsessive-compulsive symptom severity (CGI-S) at post (proportional OR = 0.09, 95% CI 0.01 to 0.43), SE = .84, p = .005) and at 3-month follow-up (pOR = 0.15, 95% CI 0.02 to 0.87), SE = 0.93, p = .041. CGI-I data was not extractable.  - 16% classified as responders (35% or more reduction in Y-BOCS score and CGI-I of 1 or 2) at post and 3-month follow-up, 21% in remission (12 points or less on the Y-BOCS) at post and 5% at 3-month follow-up.  Social outcomes using GAF:  - No statistically significant improvements in global functioning (p=0.275)  Quality of life using EQ-5D:  - No statistically significant improvements in quality of life (p = 0.832) |
| Hare et al. (2016) | Bespoke RTSM | **Primary and secondary outcome measures not specified.**  Hospital Anxiety and Depression Scale (HADS) - self-report measure of anxious and depressed feelings.  Subjective ratings of anxiety - self-report of daily experience of anxiety. | Not applicable. | **Feasibility**  - *Intervention attendance rate*: gradual decrease in the number of interventions used over the RTSM phase. 6 participants did not engage in any techniques on at least 1/3 days and 4 participants did not use a technique on the final day, with 1 participant not employing any techniques at all. Participants completed more than half of the questionnaires during baseline and RTSM (64.7% and 54.3%, respectively).  - *Intervention drop-out*: 14 participants started the study, with four dropping out within 3 days due to technical problems and one lost contact. The final sample was 9.  **Acceptability**  - *Experience of care:* Participants commented that responding to questions was straightforward and similar to a mobile phone and caused little if any disruption. There was some frustration when a trial interrupted an activity. Most participants reported some degree of interference with their daily lives. Some participants reporting frustration due to the unpredictability of the PDA beeps and/or waiting for beeps. | **Effect of time (no comparison group)**  Mental health outcomes using Subjective ratings of anxiety; & HADS:  - During the RTSM, elevated anxiety levels (i.e., >3) were reported on 51 (33.8%) occasions, with participants employing various techniques of which 58.8% (n=30) were successful. Most (N=33, 66.0%) post-technique anxiety ratings (x=2.84; SD=0.77) remained elevated (i.e., >3), and participants attempted a second technique on only seven occasions (18.2%), after which anxiety ratings remained elevated on five occasions (x=3.00; SD=0.82).  - No significant improvements in aggregated mean anxiety from first and last days of Baseline and RTSM phases (baseline first day M = 2.71, SD = 1.14 vs last day M = 2.92, SD = 0.99, p = 0.345; RTSM first day M = 2.46, SD = 0.75 vs last day M = 2.09 SD = 0.62, p = 0.0075). Significant improvements in aggregated mean anxiety by phase from start of baseline (M = 2.92, SD = 0.96) to end of RTSM phase (M = 2.26, SD = 0.56), p = 0.012. Significant improvements in subjective anxiety from pre (M = 3.31, SD = 0.28) to post-technique (M = 2.91, SD = 0.51), p = .018.  - No significant improvements in anxiety and depression from baseline to post-RTSM phase (HADS anxiety: M = 10.12, SD=2.70, p=0.21; HADS depression: M = 4.25, SD=3.28, p=0.75). |
| Harrison at al. (2020) | Detection of autism | **Primary and secondary outcome measures not specified.**  Personality Assessment Inventory (PAI) - self-report assessing a variety of personality and psychopathology domains. | **Increase knowledge and detection of autism**  - Introduction of screening tools for the detection of autism | Not applicable. | **Effectiveness in detecting autism in mental health services**  Autism detection using PAI:  - An Autism-Spectrum Disorder-Discriminant Function (ASD-DF) was derived consisting of 18 variables. Classification results of the discriminant analysis revealed that the ASD-DF significantly differentiated the ASD sample from the combined randomly selected (two-thirds) clinical/inpatient contrast sample (Canonical r=0.58; Wilks’ lambda=0.67, p <.001). Cut-off of 0.8407 yielded results that maximized the combination of sensitivity 87.6% and specificity 86.9%.  - Cross-validation showed that the ASD-DF continued to be quite effective at discriminating between the ASD sample and the remaining one-third combined inpatient and standardised samples.  - **ASD diagnosis** significantly predicted the ASD-DF in the combined samples after covarying out the effect of gender (p<0.001), with a large effect size (Partial eta2=0.31). **Sex** was determined to be a significant covariate, (p<0.01), but the effect size was quite small (Partial eta2=0.006). Likewise, the difference in ASD-DF scores between men (1.89) and women (1.95) was not significant in the ASD sample, p=0.759 |
| Helverschou et al. (2021) | Autism, Intellectual Disability and Psychiatric Disorder (AUP) Network | **Primary and secondary outcome measures not specified.**  Psychopathology in Autism Checklist (PAC): caregiver-report of mental health. | **Increase knowledge and detection of autism**  - Clinician training and skills | Not applicable. | **Effect of time (no comparison group)**  Psychiatric assessment using PAC:  - Significant improvements in proportion with *psychiatric disorders* from referral to after 12 months p < .001, but not from after 12 months to 24-27 months. |
| Horwood et al et al. (2021) | Adapted guided self-help CBT for depression vs TAU | Topic guides for trial participants and coach interviews were developed and modified throughout the study. The topic guides covered several areas including acceptability. | **Communication accommodations**  - Plan in advance  - Use of simple, written material and visual aids  **Accommodate individual differences**  - Evaluate individual needs and preferences  - Tailor practice to individual needs and preferences  **Intervention structure**  - Format of intervention  **Intervention content**  - Simplified and structured content  - Integration of cognitive-behavioural approaches | (This publication is linked with Russell et al., 2020)  **Acceptability**  Acceptability of guided self-help (Themes):  - *Experience of guided self-help arm:* All coaches valued the comprehensive training and supervision, particularly the underlying communication approaches and the emphasis on behavioural activation rather than cognition, which they felt was ideally suited for autistic adults.  - *Session content, pacing and structure:* Most participants viewed the session content and pacing of the session positively. However, the pacing for some was a little slow in the early sessions and for some it was difficult to take in all the information in the allocated time. All participants valued the guided element and appreciated that the coaches were relaxed and interested. Participants appreciated coaches using plain language to explain key concepts, checking understanding and encouraging questions. Both participants and coaches valued the concrete and structured approach of the sessions. Participants also liked the format, design and clarity of the printed materials. Most participants commented that the homework tasks were acceptable and feasible.  - *Guided self-help use of visual information:* Participants found the use of visual tools to record daily activities, rate feelings and notice links between situations and feelings generally helpful. However, for some, a pre-defined template was not ideal and individually tailored one was preferred. Additionally, noticing and rating mood was not easy for some even with an individualised chart.  - *Treatment goals:* most participants noted that their primary goal was to improve their low mood and that this had been met by the end of treatment. A minority did not have a clear goal. Most coaches found setting treatment goals challenging. Reviewing goals at the start and end was viewed as helpful for noticing change by both participant groups. | Not applicable. |
| Jones et al. (2021) | Not applicable. | Survey covering a range of domains including adaptations within the settings. | **Increase knowledge and detection of autism**  - Introduction of screening tools for the detection of autism  **Environmental adjustments**  - Provide environmental and practical adjustments  - Normalise the use of sensory resources and stimming  **Communication accommodations**  - Use of simple, written material and visual aids  - Provide communication support  **Accommodate individual differences**  - Evaluate individual needs and preferences  - Tailor practice to individual needs and preferences | **Feasibility**  *Prevalence of adaptations across participating clinics:* 81% of in-patient units (64/79 respondents) have specific assessments on ‘likes and dislikes’ of patients with autism; 82% of in-patient units (65/79 respondents) have assessments of coping strategies; Care plans based on individual needs specific to people with autism were available in 71% of units (53/75 of respondents); Only two-third of units (66%, 52/79 respondents) provided communication passports; 62% (49/79 respondents) a bespoke sensory assessment; The presence of a standardised protocol for people with autism (specific protocol for admission, assessment and management of people with autism) was available only in a fifth of the respondent’s units (21%, 17/79 respondents); Of all units 63% provided visual signage or orientation tools; 76% were able to provide visual timetables; 74% units were able to provide visual help/cue cards; 60% units were able to provide social stories; One of seven units (14%) reported being unable to provide any extra adaptations beyond communication support for people with autism; Open access low-stimulus area (52% of units providing this, 41/79 respondents); On request low-stimulus area (42%, 33/79); Scheduled access low-stimulus area (15%, 12/79); Lighting adaptations (23%, 18/79); Ability to adapt meal plans to sensory requirements (51%, 40/79); Noise adaptations (14%, 11/79); Other adaptations (4%, 3/79); No adaptations provided (15%, 12/79).  - The survey looked at the assessments in place for in-patient services to support people with autism in a person-centred manner as per current good practice. 90% (71/79 respondents) of units reported offering autism assessment.  - Other adaptations (i.e tools/strategies designed for specifically supporting autistic people) mentioned in the free text as made available in the respondent's in-patient units included ear defenders, weighted blankets, stress ball and relaxing music (no proportions provided in paper). | Not applicable. |
| Kiep et al. (2015) | Adapted MBT-AS | **Primary and secondary outcome measures not specified.**  Symptom Checklist-90-revised (SCL-90-R) - self-report symptom inventory.  The Rumination-Reflection Questionnaire (RRG) - self-report.  The Dutch Mood Scale (GMS): self-report measuring positive affect. | **Communication accommodations**  - Use of simple and preferred language  **Intervention structure**  - Format of intervention  **Intervention content**  - Simplified and structured content | **Feasibility:**  - *Intervention attendance rate:* 8 of the participants dropped out of the study before completion, thus data was collected on 50/58 participants.  - *Intervention drop-out rates:* 8/58 participants dropped out of the study before completion | **Effect of time (no comparison group).** **(The data of 20/50 subjects used in this study have been used in earlier research of Spek et al., 2013)**  Mental health outcomes using SCL-90-R, GMS, and RRG: A significant main effect for time was found on the following variables: *somatization* (p=0.002), *inadequacy in thinking and acting* (p=0.000), *depression* (p=0.000), *agoraphobia* (p=0.008), *distrust and interpersonal sensitivity* (p=0.002), *sleeping problems* (p=0.000), *general psychological and physical well-being* (p=0.000), and *rumination* (p=0.000).  The difference between the first and second (right after completing treatment) evaluations are significant for all dependent variables (SCL-90 subscales, rumination RRQ subscale, positive affect GMS subscale; p value<0.01), whereas there are no significant differences on any of the scales between the second and third (9 weeks after completing treatment) evaluations (lowest p value=0.187). This indicates significant positive effects of MBT-AS right after completing treatment, which remains stable over a 9-week period after completing therapy. |
| Langdon et al. (2016) | Bespoke CBT for anxiety vs waiting list | **Primary Outcome Measure**  Hamilton Rating Scale for Anxiety (HAM-A): clinician-rated scale of anxiety symptoms.  **Secondary Outcome Measures**  Social Phobia Inventory: self-report measure of behavioural, physiological and cognitive symptoms associated with social phobia.  Liebowitz Social Anxiety Scale: self-report measure of fear and avoidance throughout 24 listed situations likely to elicit social anxiety.  Social and Emotional Functioning Interview (Informant and Subject Versions): semi-structured clinician-rated assessment of everyday social and psychiatric functioning.  Social Interaction Anxiety Scale: self-report measure of anxiety as experienced in social situations associated with social anxiety and social phobia in accordance to the DSM-IV criteria.  Fear Questionnaire: self-report assessing individual perception of fears and phobias.  Hamilton Rating Scale for Depression (HAM-D): clinician-rated interview assessing depression symptom severity.  **Interviews about experiences of care:** Following the completion of the trial, participants were interviewed, and asked to rate nine questions on a 5-point Likert Scale about their experience of receiving therapy. Participants were also asked (1)‘What were you hoping for by taking part in this research study?’, (2)‘What was best about the group?’, (3)‘What was worst about the group?’, (4)‘What advice would you give for the next group?’ and (5)‘Were there any difficulties you feel that the group did not address?’ | Not applicable. | **Feasibility**  - *Intervention attendance rate:* Mean=13.3 treatment sessions, SD=7.17  - *Intervention drop-out rate:* During the trial, seven participants were lost (5 from intervention arm and 2 from control arm), representing an attrition rate of 13%.  **Acceptability**  - *Experience of receiving care (quantitative):* 53% of the participants agreed or strongly agreed that the individual sessions that were initially offered helped prepare them for the group sessions. 59% of the participants agreed or strongly agreed that they now knew how to reduce their feelings of anxiety following treatment. However, 38% of participants thought there was insufficient time during sessions and 41% thought there were too few sessions. 79% of participants agreed or strongly agreed that they found listening to the problems of others helpful, while nearly 80% agreed or strongly agreed that they felt supported by other group members. 56% agreed or strongly agreed that therapy reduced their anxiety, while 44% were neutral, disagreed, or strongly disagreed on this. 73% of participants agreed or strongly agreed that they would recommend therapy to others, and 73% agreed or strongly agreed that therapy was helpful.  - *Experience of care (qualitative):*  1) ‘Motivation to take part’. Participants described taking part in the trial in order to access help for their mental health problems, while others had hoped that they might form new relationships with other people with ASDs.  2) ‘Positive experiences’. Participants described that they enjoyed 'interacting with the others'  3) ‘Negative experiences’. Many participants were clear that they wanted to have had longer sessions. Others spoke about issues around the dynamics of being in a group, with one participant stating, ‘the group could be easily hijacked’. Several spoke about needing more continuity and greater focus on making sure the sessions flowed more effectively, while there were a few participants who commented that they found taking part in a group very difficult and thought the whole experience was negative.  4) ‘Further adaptations’. Participants indicated they may benefit from more individual sessions, and the suggestion to alternate between blocks of both group and individual sessions might improve treatment efficacy. This would also help to ensure that clients are afforded sufficient time to address their difficulties. Participants asked for more innovative homework options, using technology.  5) ‘Pragmatic issues’. Participants told us that there were sometimes issues with public transport, travelling, the timings of the group, heating in the rooms and difficulties with parking, all of which they did not like." | **Primary outcome**  Mental health outcome using HAM-A:  HAM-A mean scores significantly improved over time, regardless of arm, and regardless of baseline scores, p<0.001. Controlling for baseline scores, there was no significant difference between the treatment and wait list arms at either follow-up 1 (after initial 24 weeks of treatment) or 2 (after further 24 weeks of treatment) on the HAM-A.  **Secondary outcomes**  Mental health outcomes using Social Phobia Inventory; Liebowitz Social Anxiety Scale; Social Interaction Anxiety Scale; Fear Questionnaire; & HAM-D:  There was a significant improvement over time, regardless of arm, and baseline scores, on the HAM-D, P=0.008; Fear Questionnaire Total Phobia Score, P=0.019; Liebowitz Avoidance, P=0.003; Liebowitz Fear/Anxiety, P<0.002. There was a significant improvement over time, regardless of arm, and baseline scores on the Social Interaction Anxiety Scale, P<0.001; Social Phobia Inventory, P=0.007  Controlling for baseline scores, there was no significant difference between the treatment and wait list arms on any of the secondary outcomes at follow-up 1 (after initial 24 weeks of treatment) or 2 (after further 24 weeks of treatment).  Social outcomes using Social and Emotional Functioning Interview (Informant and Subject Versions)  There was a significant improvement over time, regardless of arm, and baseline scores on the Social/Emotional Functioning Interview–Informant, P<0.001; and Social/Emotional Functioning Interview–Subject Versions, P<0.001.  Controlling for baseline scores, there was no significant difference between the treatment and wait list arms on any of the secondary outcomes at follow-up 1 (after initial 24 weeks of treatment) or 2 (after further 24 weeks of treatment). |
| Lobregt-van Buuren et al. (2019) | Adapted EMDR | **Primary and secondary outcome measures not specified.**  Impact of Event Scale Revised (IES-R) - self-report measure of PTSD symptoms.  Adapted Anxiety Disorders Interview Schedule-Children (ADIS-C) section PTSD version for adults (semi-structured interview) - assesses trauma, adverse events and trauma related symptoms in adults with mild to borderline intellectual disabilities and to establish a PTSD diagnosis according to DSM-IV-TR and DSM-5.  Brief Symptom Inventory (BSI) - self-report measure of psychological distress and symptoms of psychopathology. | **Communication accommodations**  - Use of simple and preferred language | **Feasibility**  - *Intervention drop-out rate:* 5 (18.5%) participants dropped out due to: 4 dropped out during the waiting period for EMDR as a result of perceiving EMDR on top of TAU too time-consuming anticipating problems at work and travelling time (n=1), physical health problems (n=1), travelling time to EMDR therapist (n=1), no willingness to fill out questionnaires (n=1), 1 dropped out over the course of EMDR therapy due to suicidal ideation in response to increased problems at home (n=1). 1 additional participant who completed EMDR therapy was excluded from analysis due to their measurements being unusable.  **Acceptability**  - *Satisfaction with care:* All participants described in the follow-up session that they had found the EMDR sessions stressful. 86% of the participants indicated that they would choose EMDR therapy again. | Mental health outcomes using IES-R; Adapted ADIS-C section PTSD version for adults & BSI  - A significant multivariate Time effect was found, p < 0.001, resulting from significant changes in mean scores on: the thermometer card of the Adapted ADIS-C section PTSD, p < 0.001, the IES-R, p < 0.001, and the BSI, p < 0.001.  - Mean score of the thermometer card of the Adapted ADIS-C section PTSD did not vary significantly between T1 (baseline) and T2 (after 6-8 weeks TAU), but the mean score decreased significantly at T3 (after max 8 EMDR sessions + TAU) and T4 (after further 6-8 weeks of TAU only) showing a large effect size (d = 1.81) on T3 and a moderate effect size (d = 0.62) on T4, p<0.05.  - Mean IES-R score did not differ significantly between T1 and T2, but it decreased significantly at T3 (d = 1.16, p<0.05), and remained stable until T4.  - Mean BSI score did not change significantly between T1 and T2, but decreased significantly at T3 (d = 0.93, p<0.05), and remained stable until T4. |
| Maskey et al. (2019) | Bespoke CBT for anxiety in combination with virtual reality | **Primary and secondary outcomes not specified.**  Beck Anxiety Inventory (BAI): self-report measuring the severity of anxiety.  Generalized Anxiety Disorder 7 (GAD-7): self-report measuring severity of generalized anxiety disorder.  Patient Health Questionnaire-9 (PHQ-9): self-report measuring symptoms of depression.  WHOQOL-BREF: self-report measuring 4 quality of life domains: physical health (7 items), psychological health (6 items), social relationships (3 items), and environment (8 items).  Target Behaviours: were used to identify symptom change over time for the phobia targeted in the treatment having identified a specific anxiety target, questions such as ‘‘how often?’’ and ‘‘how distressed?’’ are asked in a standard interview format to the participant and their supporter. This enabled a researcher to write a vignette describing the anxiety and its impact at the three timepoints. | Not applicable | **Feasibility**  - *Intervention attendance rate:* 100% (n=8), each participant completed all sessions showing that the intervention is feasible and acceptable.  - Intervention drop-out: there were no dropouts. | **Effect of time (no comparison group).**  The study did not statistically analyse differences over time on BAI, GAD-7, PHQ-9 and WHOQOL-BREF scores.  Mental health outcomes using Target Behaviours:  - 5 of the 8 participants were classified as treatment responders (mean score of 3.0 or less as rated by the four-person rating panel) on the change in Target Situation Rating at 6 weeks and at 6 months post-intervention. Of these 5, 4 had a score of 1.0 or 1.25 at 6 months postintervention, indicating that the participant was able to function normally without any impact from the phobia. The 5 responders showed a pattern of increasing improvement with time as indicated by the improvement in target behaviour scores from 6 weeks to 6 months follow-up, indicating a strengthening of the treatment effect over time. Three of the participants were non-responders to treatment, each scoring 4.0 (equivocally improved and indicating no worsening of symptoms). |
| McGillivray et al. (2014) | Bespoke CBT for anxiety, stress and depression vs waiting list | **Primary outcomes and secondary outcomes not specified.** **Depression Anxiety Stress Scales (DASS) - self-report measure of depression, anxiety and stress.**  Automatic thoughts questionnaire (ATQ) - self-report measure of frequency of cognitive self-statements associated with depressed mood.  Anxious self-statements questionnaire (ASSQ) - self-report measure of frequency of cognitive self-statements associated with anxiety.  Assessment questionnaire (self-report) asking about any treatments for anxiety or depression they had received during the past 3 months. | Not applicable | **Feasibility**  - *Intervention attendance rate:* 6/16 waitlist control group completed the programme, but it is unclear how many in the treatment group completed treatment.  - *Intervention drop-out rate:* All participants (n=42: CBT group = 26, waiting list group = 16) completed pre- and post-group assessments. Participants in the waitlist group were then invited to take part in the intervention program. 6 of these participants subsequently completed the program, making the total of 32 people who finished the intervention program. Of the remaining participants from the waitlist group, 4 started the program but dropped out and 6 reported that they were no longer interested in participating due to changed personal circumstances. Participants were invited to return for a follow-up group session to undertake repeat assessments at 3 and 9 months after completion of the program. 27 participants completed the 3- and 9-month follow-up indicating a small attrition rate. | Mental health outcomes using DASS; ATQ; ASSQ:  - DASS overall: There was no significant effect for Group X Time interaction (p>0.05). Participants improved over time regardless of whether they were in the treatment group or on the waitlist.  - ASSQ: no significant effect for group X time interaction (p>0.05)  - ATQ: no significant effect for group X time interaction (p>0.05)  - A subsequent analysis was undertaken with only those people who scored above the normal range on the depression (>9), anxiety (>7) and stress (>14) subscales of the DASS, the ATQ >60 and the ASSQ >64. For the depression DASS subscale, a significant effect for Group X Time interaction (p<0.05) was found for these participants. Significant effect for Group x Time interaction (p<0.05) was seen for the DASS stress subscale, but not for anxiety DASS subscale (p>0.05).  - Gor people who scored over 60 on the ATQ, there was no significant effect for Group X Time interaction (p>0.05).  - For those who scored over 64 on the ASSQ, there was no significant effect for Group X Time interaction (p>0.05)  - In order to determine whether the reduction of scores in the DASS depression and stress subscales was sustained over time, repeated measures analysis was conducted. Participants who were symptomatic and who had completed the treatment program and post group assessments were included. A significant main effect for time was found for the DASS depression subscale, p<0.01; and the DASS stress subscale, p<0.01. Follow up comparisons found that significant differences occurred between pre-group and post-group assessments, with no differences evident between scores at the 3- and 9-month follow-ups. Scores at the 9-month follow-up remained significantly lower than those obtained at the pre-program assessment.  Service use using a self-report assessment questionnaire:  - Most participants (78%) reported no change in their receipt of additional mental health treatments during the course of the intervention phase. During the study period, additional treatment for depression and for anxiety had been initiated for two (6.3%) and three participants (9.4%) respectively, while additional treatment for depression and for anxiety had been discontinued for five (15.6 %) and four participants (12.5 %) respectively. Treatment outside of the group CBT intervention was thus relatively stable over the study period. |
| Oshima et al. (2021) | Adapted schema therapy | **Primary outcomes**  Global assessment functioning (GAF) scale - an interview rating of social functioning.  The World Health Organization quality of life assessment brief (WHO QOL-BREF) - self-report measure of subjective feeling of social adaptiveness and quality of life.  **Secondary outcomes**  Beck Depression Inventory (BDI-II).  State-Trait Anxiety Inventory (STAI).  Liebowitz Social Anxiety Scale (LSAS).  Obsessive-Compulsive Inventory (OCI). | **Intervention content**  Consider the role of autism | **Feasibility**  - *Intervention drop-out rate:* 2/12 participants dropped out of the intervention (n = 1 due to hospital admission, n = 1 due to moving out of the city). | **Effect of time (no comparison group).**  **Primary outcomes**  Social outcomes using GAF:  - Global functioning measured on GAF changed significantly over time (p < 0.01). A post hoc analysis showed significant differences between pre- and post-treatment (p < 0.001), and between pre-treatment and follow-up (12 weeks after completion of treatment) (p < 0.001, d = 3.35).  Quality of life using WHO QOL-BREF:  - Quality of life also changed significantly over time (p < 0.01), however, the results did not remain significant between each time point after correction (p > 0.05).  **Secondary outcomes**  Mental health outcomes using BDI-II; STAI; LSAS; & OCI:  - BDI-II: Depression scores changed significantly over time (p < 0.01), but this did not remain significant after post-hoc correction (p > 0.05).  - OCI: Obsessive compulsive symptoms did not change significantly over time (p > 0.05).  - STAI (state): State anxiety scores changed significantly over time (p < 0.05), but this did not remain significant after post-hoc correction (p > 0.05). STAI (trait): Trait anxiety scores changed significantly over time (p < 0.05), but this did not remain significant after post-hoc correction (p > 0.05).  - LSAS total: scores did not change significantly over time (p > 0.05). LSAS (fear/anxiety): scores changed significantly over time (p < 0.05), but this did not remain significant after post-hoc correction (p > 0.05). LSAS (escape): scores did not change significantly over time (F(1.27, 11.46) = 0.59, p > 0.05). |
| Pahnke et al. (2019) | Adapted acceptance and commitment therapy | **Primary and secondary outcome measures not specified.**  ASD-adapted version of the Treatment Credibility Scale - self-report measure of treatment credibility.  Perceived Stress Scale (PSS-14) - self-report measure of subjective stress.  Satisfaction with Life Scale (SWLS) - self-report of quality of life.  Beck Depression Inventory-II (BDI-II) - self-report measure of depressive symptoms.  Beck Anxiety Inventory (BAI) - self-report measure of anxiety symptoms.  Sheehan Disability Scale (SDS) - self-report measure of functional impairment as related to familial, social and vocational aspects of life. | **Communication accommodations**  - Clear communication  - Use of simple and preferred language  - Use of simple, written material and visual aids  **Intervention structure**  - Format of intervention  **Intervention content**  - Integration of emotion-focused strategies | **Feasibility**  - *Intervention attendance rate:* 9/10 completed the treatment, the mean number of sessions attended was 11 sessions (SD = 3, range: 9-12)  - *Intervention drop-out rate:* 1/10 participants dropped out, post-treatment after two sessions  - *Homework compliance:* Overall, participants were successful in completing homework assignments, as well as carrying out mindfulness and exercises at home.  **Acceptability**  - *Treatment credibility:* The treatment credibility total score on the TCS was high (M = 7.7, SD = 0.8). | **Effect of time (no comparison group).** Mental health outcomes using PSS-14; BDI-II; & BAI:  - PSS-14: significant improvements in perceived stress from pre (M = 35.1, SD = 5.4) to post (M = 29.0, SD = 7.7, p < 0.05.) No significant change from pre to 3-month follow-up (M = 31.5, SD = 8.3, p > 0.05)  - BDI-II: significant improvement in depressive symptoms from pre (M=21.6, SD=14.3) to 3-month follow-up (M=14.4, SD=11.6, p<0.05). No significant change from pre to post (M=15.3, SD=10.7, p > 0.05)  - BAI: No significant change from pre (M= 24.2, SD=16.4) to post (M=14.5, SD=9.5) or from pre to 3-month follow-up (M=18.4, SD=11), p>0.05.  Quality of life outcome using SWLS:  - Satisfaction with Life Scale: non-significant change in satisfaction with life between pre (M = 13.2, SD = 5.1) to post (M = 15.5, SD = 5.7, t = -1.54, p > 0.05). Significant increase in satisfaction with life between pre and 3-month follow-up (M = 17.0, SD = 4.8, t = -2.79, p < 0.05)  Social outcome using SDS:  - SDS (work): No significant change in disability at work/school between pre (M = 6.7, SD = 2.8) and post (M = 7.1, SD = 1.7, t = -0.51, p > 0.05). No significant change in disability at work/school between pre and 3-month follow-up (M = 6.2, SD = 2.5, t = 0.75, p > 0.05).  - SDS (social): Significant reduction in disability in social life/leisure activities between pre (M = 7.6, SD = 3.0) and post (M = 6.2, SD = 3.2, t = 2.69, p < 0.05). Significant reduction in disability in social life/leisure activities between pre and 3-month follow-up (M = 6.2, SD = 3.2, t = 2.45, p < 0.05).  - SDS (family): No significant change in disability in family life/home responsibilities between pre (M = 6.6, SD = 2.0) and post (M = 5.9, SD = 2.0, t = 0.96, p > 0.05). No significant change in disability in family life/home responsibilities between pre and 3-months follow-up (M = 5.9, SD = 2.0, t = 9,64, p > 0.05). |
| Petty et al. (2021) | Not applicable. | Freelisting interview guide was developed, which included questions in relation to 1) adaptations made prior to or within a client appointment 2) service-level adaptations 3) adaptations made in the physical environment and 4) adaptations relating to a person's gender. | **Increase knowledge and detection of autism**  - Clinician training and skills  **Environmental adjustments**  - Provide environmental and practical adjustments  - Normalise the use of sensory resources and stimming  **Communication accommodations**  - Plan in advance  - Clear communication  - Use of simple and preferred language  - Use of simple, written material and visual aids  **Accommodate individual differences**  - Evaluate individual needs and preferences  - Tailor practice to individual needs and preferences | **Feasibility:**  *Adaptations before attending an appointment (Salient items are considered to be consensus responses to the question):*  - Check the suitability of the sensory environment - listed by 60% of staff, mean rank = 6.2, salience = .334 (salient)  - Find out about the client in advance - listed by 33% of staff, mean rank = 1.6, salience = .306 (salient)  - Be clear in communication - listed by 33% of staff, mean rank = 4.6, salience = .206 (salient)  - Check suitability of lighting - listed by 33% of staff, mean rank = 7.6, salience = .179 (salient)  - Ensure the client is prepared for what will happen - listed by 20% of staff, mean rank = 3.0, salience = .168 (salient)  - Find out if the client has sensory needs - listed by 27% of staff, mean rank = 6.3, salience = .141 (not salient)  - Be prepared to adjust communication - listed by 20% of staff, mean rank = 4.3, salience = .141 (not salient)  - Find out about the client in advance from significant people - listed by 20% of staff, mean rank = 5.3, salience = .139 (not salient)  - Find out about the client in advance from case notes - listed by 13% of staff, mean rank = 1.5, salience = .129 (not salient)  - Ensure the client is prepared about the purpose of the appointment - listed by 13% of staff, mean rank = 3.0, salience = .117 (not salient)  *Service-level adaptations:*  - Provide a sensory friendly environment - listed by 53% of staff, mean rank = 2.6, salience = .448 (salient)  - Make information available for clients about the service - listed by 46% of staff, mean rank = 4.1, salience = .346 (salient)  - Ensure suitable noise levels - listed by 53% of staff, mean rank = 4.4, salience = .334 (salient)  - Adapt communication - listed by 40% of staff, mean rank = 4.7, salience = .244 (salient)  - Keep to plain design - listed by 33% of staff, mean rank = 3.4, salience = .230 (salient)  - Offer flexible session timings - listed by 33% of staff, mean rank = 3.2, salience = .203 (salient)  - Maintain the specialist skillset of staff - listed by 27% of staff, mean rank = 2.8, salience = .181 (salient)  - Adapt written correspondence - listed by 20% of staff, mean rank = 2.0, salience = .181 (salient)  - Utilise a protected building or space - listed by 20% of staff, mean rank = 2.0, salience = .167 (salient)  - Use signs up to modify the environment - listed by 20% of staff, mean rank = 3.3, salience = .134 (not salient)  *Adaptations within a client appointment:*  - Communicate clearly - listed by 60% of staff, mean rank = 2.0, salience = .536 (salient)  - Avoid ambiguity - listed by 27% of staff, mean rank = 2.5, salience = .231 (salient)  - Offer a flexible and individualised approach - listed by 33% of staff, mean rank = 5.0, salience = .204 (salient)  - Check for understanding - listed by 27% of staff, mean rank = 4.8, salience = .180 (not salient)  - Agree etiquette for making eye contact - listed by 27% of staff, mean rank = 3.5, salience = .177 (not salient)  - Slow the pace of communication - listed by 20% of staff, mean rank = 2.7, salience = .169 (not salient)  - Avoid idioms - listed by 20% of staff, mean rank, mean rank = 3.3, salience = .159 (not salient)  - Monitor own communication - listed by 20% of staff, mean rank = 3.0, salience = .156 (not salient)  - Consider the room seating arrangement - listed by 20% of staff, mean rank = 3.3, salience = 1.52 (not salient)  - Use agendas - listed by 27%, mean rank = 6.5, salience = .151 (not salient)  *Gender:*  - Know how someone identifies - listed by 60% of staff, rank = 1.9, salience = .444 (salient)  - Maintain awareness of gender differences - listed by 60% of staff, mean rank = 2.4, salience = 4.06 (salient)  - Do not make assumptions - listed by 33% of staff, mean rank = 2.6, salience = .243 (salient)  - Know pronoun or name preferences -listed by 40% of staff, mean rank = 2.7, salience = .230 (salient)  - Use a preference notifications system - listed by 13% of staff, mean rank = 1.5, salience = .126 (not salient)  - Maintain awareness of gendered socialisation - listed by 13% of staff, mean rank = 2.5, salience = .100 (not salient)  - Check suitability of clinician gender - listed by 13% of staff, mean rank = 3.0, salience = .083 (not salient)  - Adapt questioning for female representation - listed by 13% of staff, mean rank = 3.0, salience = .067 (not salient)  - Offer gender appropriate resources - listed by 7% of staff, mean rank = 1.0, salience = .067 (not salient)  *Adaptations to the physical environment:*  - Reduce noise - listed by 80% of staff, mean rank = 3.8, salience .559 (salient)  - Provide adjustable lighting - listed by 60% of staff, mean rank = 3.6, salience = .442 (salient)  - Neutralise decor - listed by 47% of staff, mean rank = 4.4, salience = .306 (salient)  - Offer space - listed by 40% of staff, mean rank = 2.7, salience = .279 (salient)  - Reduce scents - listed by 33% of staff, mean rank = 5.4, salience = .200 (not salient)  - Neutralise all sensory demands - listed by 27% of staff, mean rank = 4.0, salience = .172 (not salient)  - Control outside noise - listed by 33% of staff, mean rank = 6.2, salience = .162 (not salient)  - Avoid patterns - listed by 20% of staff, mean rank = 3.3, salience = .144 (not salient)  - Reduce the number of items in the environment - listed by 20% of staff, mean rank = 4.3, salience = .138 (not salient)  - Provide sensory resources - listed by 27% of staff, mean rank = 6.0, salience = .129 (not salient) | Not applicable. |
| Russell et al. (2013) | Adapted CBT for OCD vs adapted anxiety management (AM) | **Primary outcome** (clinician-administered)  Yale-Brown Obsessive Compulsive Severity Scale (YBOCS).  **Secondary outcomes**  Clinician-administered:  Clinical Global Impression Rating Scales (CGI).  CGI improvement (CGIi).  Dimensional YBOCS (D-YBOCS): semi structured interview to ascertain the presence and severity of six symptom dimensions of OCD.  Self-report adult measures:  Obsessive Compulsive Inventory-Revised (OCI-R).  Beck Depression Inventory (BDI).  Beck Anxiety Inventory (BAI).  Liebowitz Social Anxiety Scale (LSAS).  Work and Social Adjustment Scale (WSAS). Self-report scales for youth ages 14-18:  Obsessive Compulsive Inventory-Revised (OCI-R).  Beck Depression Inventory-Youth (BDI-Youth).  Work and Social Adjustment Scale (WSAS).  Spence Children's Anxiety Scale (SCAS).  All participants:  Treatment satisfaction: a self-rated 8-point visual analogue scale.  Children's Obsessive-Compulsive Inventory-Parent Version (PR-CHOCI-R): parental/carer report. | **Communication accommodations**  - Use of simple and preferred language  - Use of simple, written material and visual aids  **Intervention content**  - Simplified and structured content  - Integration of emotion-focused strategies  - Integration of cognitive-behavioural approaches | **Feasibility**  *- Intervention drop-out rate:* 20/23 people in each arm completed treatment  *- Intervention attendance rate:* The mean number of treatment sessions was marginally greater in the CBT (M = 17.43, SD = 4.3) group than in the control group (M = 14.43, SD = 5.3, t = -2.022, p = 0.03; 95% confidence interval: -5.98 to -0.006). The mean number of weeks between pre-treatment end-of-treatment ratings were: AM group=23.74 (SD=10.37); CBT for OCD=27.06 (SD=10.27). The most usual length of treatment in weeks was 25.  **Acceptability**  - *Treatment satisfaction:* There were no differences between the two treatment groups in their reports of treatment satisfaction. Anxiety management group mean satisfaction score = 5.60 (SD = 2.131); CBT group mean satisfaction score = 4.9 (SD = 2.3), p = 0.425.  *- Cross-over cases:* Nine (39%) participants in the anxiety management comparison group, compared with three (13%) participants in the CBT group, asked to "crossover" or try the other treatment either at or after the 1-month follow-up point (χ2 = 4.05, p = 0.044). 8/9 participants originally randomised to anxiety management who "crossed over" to CBT completed the second treatment and attended for symptom ratings | **Primary Outcome**  Mental health primary outcome using YBOCS  - There was no significant difference between treatment groups on YBOCS total severity scores (primary outcome measure) at the end of treatment when controlling for pre-treatment YBOCS severity ratings (p = 0.295)  - There were significant changes in YBOCS total severity ratings from pre-treatment to end of treatment in both the CBT group (p = 0.001) and the anxiety management control group (p<0.0001). The YBOCS total severity scores effect size was large and clinically meaningful in the CBT group (1.15) and medium in the anxiety management control group (0.6)  - There were more treatment responders (i.e., had a >25% reduction in YBOCS total severity ratings) in the CBT group as compared to the AM group (9/20 (45%) versus 5/20 (20%), respectively). However, this difference in response rate was not statistically significant (χ2 = 1.72, p = 0.160). When a more stringent rating of treatment response (a CGI “much or very much improved” combined with a >35% reduction in YBOCS total severity ratings) was considered, 6/20 (30%) of the CBT group achieved treatment response compared with 2/20 (10%) of the anxiety management control group. Again, the groups did not differ significantly in the proportion of treatment responders. Slightly more participants in the CBT group were classified as remitted cases (i.e., with a YBOCS total severity rating of ≤12 1 week after treatment ended) as compared with the anxiety management control group (5/20 (20%) versus 3/20 (15%)) but this difference was not statistically significant.  - Cohen’s d effect sizes were 0.4 for the YBOCS total severity rating, 0.4 for YBOCS obsessions severity, 0.2 for YBOCS compulsions, and 0.3 for CGI, all indicating a small advantage for CBT over the anxiety management control group after treatment.  -In the CBT group, changes in YBOCS total severity scores compared to pre-treatment remained significant at 1-month follow-up (p = 0.005), 3-month follow-up (n = 10, p = 0.008), 6-month follow-up (n = 12, p = 0.007), and 12-month follow-up (n = 11; p = 0.011).  **Secondary outcome**  Mental health secondary outcomes (clinician-administered) using CGI; CGIi; & D-YBOCS:  - Although clinician ratings of CGI changed significantly between pre- and post-treatment (p < 0.001), this did not vary by treatment group (p = 0.140).  - The treatment groups differ significantly in terms of the proportion of participants in each group rated as "much" or "very much improved" on the CGIi scale (CBT group n = 11; anxiety management control group n = 5 (χ2 = 3.886 (df = 1), p = 0.050).  - D-YBOCS results not reported  Mental health secondary outcomes (self-report adult) using OCI-R; BDI; BAI; & LSAS:  - Neither of the groups showed significant differences between pre-, post-, and 1-month follow-up mean scores on any of the self-ratings.  Mental health secondary outcome (self-report youth ages 14-18) using OCI-R; BDI-Youth; & SCAS:  - Neither of the groups showed significant differences between pre-, post-, and 1-month follow-up mean scores on any of the self-ratings.  Mental health secondary outcomes (parent/carer-report) using PR-CHOCI-R:  - Informant ratings differed significantly between pre- and post-treatment (improvement) only for the anxiety management comparison group (p<0.05)  Social outcome using WSAS:  - Neither of the groups showed significant differences in social functioning (WSAS) between pre-, post-, and 1-month follow-up mean scores |
| Russell et al. (2020) | Adapted guided self-help CBT for depression vs TAU | **Primary and secondary outcome measures not specified.**  Self-report questionnaire on statutory health and voluntary service use  Health care consultations (obtained from GP and secondary care records). | **Communication accommodations**  - Plan in advance  - Use of simple, written material and visual aids  **Accommodate individual differences**  - Evaluate individual needs and preferences  - Tailor practice to individual needs and preferences  **Intervention structure**  - Format of intervention  **Intervention content**  - Simplified and structured content  - Integration of cognitive-behavioural approaches | **(This publication is linked with Horwood et al., 2021)**  **Feasibility**  - *Intervention attendance rates:* 32/35 (91%) participants in the GSH group attended at least one session; 30/35 (86%) participants in the GSH group completed the pre-specified 'dose' of treatment (5 sessions); 25/35 (71%) participants in the GSH group attended all nine sessions; One participant in the GSH group was unable to attend any sessions due to ill health; By 10 weeks, 10/32 (31%) had completed the intervention, by 16 weeks, 23/32 (73%) had completed the intervention, by 24 weeks, 26/32 (84%) had completed the intervention; Mean number of treatment sessions was 7.6 (SD: 2.9, n = 35)  - *Intervention drop-out rate:* 9% (n = 3/35) of participants allocated to GSH withdrew from the trial, compared to 17% (n=6/35) of TAU participants. The timing of withdrawals in the GSH group was shortly after randomisation (n = 2) and before the final follow-up 24-weeks post-randomisation (n = 1). | The study was not powered to detect differences between the treatment groups, thus did not statistically analyse differences between the two groups on any of the outcome measures.  Service use during the study using self-report questionnaire  During the study, 11%-49% of participants attended an NHS outpatient or community mental health team clinic for mental health problems and/or visited their GP (26-46%). Participants did not report any overnight stays in an NHS hospital, or visits to a private hospital or clinic due to mental health problems, and rarely attended an Accident and Emergency department, an out-of-hours clinic or NHS walk-in centre. 23% of participants had counselling or talking therapy during the trial outside of the trial intervention. Participants used a variety of different types of help for mental health problems including support groups and received help around or outside of the home. Many participants attended a social group (24%) or drop-in service (26%) for autistic adults. Participants rarely had a home visit from any other professional (for any medical condition). |
| Sizoo et al. (2017) | Adapted mindfulness-based stress reduction and adapted CBT for anxiety and depression | **Primary and secondary outcome measures not specified.**  The Hospital Anxiety and Depression Scale (HADS) - self-reported scale. comprising of anxiety (HADS-A) and depression (HADS-D) subscales.  Global Mood Scale (GMS) - self-reported scale, gives an indication of the positive (GMS-P) and negative (GMS-N) affect resulting from general health and psychological problems.  Rumination Reflection Questionnaire (RRQ) - self-report. | **Communication accommodations**  - Plan in advance  - Clear communication  - Use of simple and preferred language  **Intervention content**  - Taking it slow | Not applicable. | Mental health outcome using HADS; GMS & RRQ:  - There was no significant group x time interaction effect for either *anxiety* or *depression* scores (p > 0.05).  - There was no significant group x time interaction effect for *positive (GMS-P) and negative (GMS-N) general mood scores* (p > 0.05).  - There was no significant group x time interaction effect for rumination (P>0.05).  Assessment points: Pre-treatment (T1), post-treatment (13 weeks, T2), and 3-months after treatment completion (T3) |
| Spain et al. (2017) | Bespoke CBT for anxiety | **Primary outcome measure:**  Liebowitz Social Anxiety Scale (LSAS) - self-report assessing anxiety about and avoidance of a range of social situations.  **Secondary outcome measures:**  The Hospital Anxiety and Depression Scale (HADS) - self-report measuring symptoms of anxiety and depression.  The Work and Social Adjustment Scale (WSAS) - self-report measuring severity of impairment.  Non-validated feedback questionnaire, designed by the group facilitators - self-report assessing satisfaction with, and acceptability of, the environment, session content, amount of time spent on each topic, strategies used and the duration and number of sessions on a Likert scale, or could be open-ended to encourage participant feedback. | Not applicable. | **Feasibility:**  *-Intervention drop-out rate:* 2/18 (11.1%) participants dropped out of the intervention, both after one session  - *Intervention attendance rate:* 16/18 (88.9%) participants completed the groups  **Acceptability:**  - *Experience of care (qualitative):* Most participants stated that they found it helpful to meet others in a similar situation. Some participants described feeling more confident in social situations (e.g., trying out new ways of conversing and incorporating a broader range of topics, as well as feeling better able to cope with and manage anxious thoughts and feelings). Participants identified as having gained an increased ability to identify different types and aspects of relationships and enhanced understanding of modes of non-verbal communication and assertiveness. Some participants stated that they would have preferred to be given additional practical strategies (e.g., for specific situations), or for the group to have incorporated additional opportunities for skills rehearsal | **Effect of time (no comparison group).** **Primary outcome measure**  Mental health outcome using LSAS:  - *Total score on the LSAS* were significantly lower post-intervention (M = 61, SD = 28.0) compared to pre-intervention (M = 80, SD = 30.7; p = 0.01; Cohen's d = 0.65).  - *LSAS fear/anxiety subscale*: There was no significant change in scores on the LSAS fear/anxiety subscale post-intervention (M = 37, SD = 12.8) compared to pre-intervention (M = 42, SD = 14.4; p = 0.34; Cohen's d = 0.39).  - *LSAS avoidance subscale*: There was no significant change in scores on the LSAS avoidance subscale post-intervention (M = 30, SD = 17.1) compared to pre-intervention (M = 38, SD = 17.2; p = 0.13; Cohen's d = 0.43).  **Secondary outcome measures**  Mental health outcome using HADS:  - There was no significant change in *HADS-A* scores post-intervention (M = 9, SD = 3.4) compared to pre-intervention (M = 10, SD = 4.8; p = 0.15; Cohen's d = 0.1).  - There was no significant change in *HADS-D* scores post-intervention (M = 8, SD = 4.0) compared to pre-intervention (M = 8, SD = 4.9; p = 0.42, Cohen's d = 0.04).  Social outcome using WSAS:  There was no significant change in *WSAS total score* (measure of social functioning) post-intervention (M = 18, SD = 7.9) compared to pre-intervention (M = 20, SD = 9.4; p = 0.32; Cohen's d = 0.20). |
| Spek et al. (2013) | Adapted MBT-AS vs waiting list | **Primary and secondary outcome measures not specified.** **The Symptom Checklist-90-revised (SCL-90-R): self-report measure of psychological distress (scores for anxiety and depression were used in this study).**  The Rumination-Reflection Questionnaire (RRG): self-report measuring rumination tendencies.  The Dutch Mood Scale (GMS): self-report measuring positive general affect. | **Communication accommodations**  - Plan in advance  - Use of simple and preferred language  **Intervention structure**  - Format of intervention  **Intervention content**  - Simplified and structured content | **Feasibility**  - *Intervention drop-out rates:* 1/21 participants dropped out of the intervention arm (due to serious illness)  - *Intervention attendance rate:* 20/21 participants did not miss more than one treatment session (exact rate not reported) | Mental health outcome using SCL-90-R; RRG & GMS:  - *SCL-90-R (depression subscale):* There was a significant group x time interaction effect for depressive symptoms measured on the SCL-90-R depression subscale (F(1, 39) = 6.15, p = .02, partial η2 = 0.14; p < 0.05). Depressive symptoms decreased more in the MBT-AS group than the control group; the difference between the groups had a medium-large effect size (Cohen's d = 0.78).  - SCL-90-R (anxiety subscale): There was a significant group x time interaction effect for symptoms of anxiety (F(1, 39) = 5.50, p = .02, partial η2 = 0.12; p < 0.05). Symptoms of anxiety decreased more in the MBT-AS group than in the control group; the difference between the groups had a medium-large effect size (Cohen's d = 0.76).  - Positive affect (GMS): There was a significant group x time interaction effect for positive affect (F(1, 39) = 6.32, p = 0.02, partial η2 = 0.14, p < 0.05). Positive affect increased more in the MBT-AS group than in the control group; the effect size was medium-large (Cohen's d = 0.79).  - Rumination-Reflection Questionnaire (RRQ): There was a significant group x time interaction effect for rumination (F(1, 39) = 15.73, p < .001, partial η2 = 0.29; p < 0.001). Rumination decreased more in the MBT-AS group than in the control group; the difference between the groups had a large effect size (Cohen's d = 1.25). |
| Tchanturia et al. (2021) | Pathway for eating disorders and autism developed from clinical experience (PEACE pathway) | **Primary and secondary outcome measures not specified.** **Hospital admissions from clinical records for 6 years before the new pathway was introduced (January 1, 2012–December 31, 2017) and 2 years after the new pathway was introduced (January 1, 2018–December 31, 2019) for patients with autism.**  Average cost of hospital admissions for inpatient eating disorder clinics by applying the national average unit cost per night for patients admitted to adult specialist ED services for the financial year 2018/19 (£532.32), taken from the National Schedule of NHS Costs. | **Increase knowledge and detection of autism**  - Clinician training and skills  **Environmental adjustments**  - Provide environmental and practical adjustments  - Normalise the use of sensory resources and stimming  **Communication accommodations**  - Provide communication support | Not applicable. | Service use using Hospital admissions from clinical records  There was a clear reduction in the *average duration of admissions* for those with co-morbid ASC since the PEACE pathway started. Mean length of admissions for patients admitted during the 6 years before the implementation of the PEACE pathway was higher for ASC patients with EDs (133 days; 19 weeks) than those without comorbid ASC (109 days; 16 weeks). This pattern reversed in the 2 years following the implementation of the PEACE pathway (comorbid ASC 90 days; 13 weeks vs. no ASC 118 days; 17 weeks).  Cost per admission using national average unit cost per night for patients admitted to adult specialist ED services for the financial year 2018/19 (£532.32), taken from the National Schedule of NHS Costs:  The *mean cost per admission* prior to the implementation of the PEACE pathway was almost £13,000 higher for ASC patients with EDs (£70,925 per patient) compared to non-ASC patients (£58,218 per patient). After the implementation of the pathway, the average cost of admissions was almost £15,000 lower for those ASC patients with EDs (£48,087 per patient) compared to those without comorbid ASC (£62,896 per patient). Given a reduction in mean cost per admission of £22,837 per ASC patient with an ED after the implementation of the pathway compared to before implementation, and an average of 12 ASC patients with EDs admitted per year, estimated total savings are in the region of £275,000 per year.  **Outcomes on the way to improving care:**  Confidence of clinicians in supporting people with comorbid autism: Evaluation of 100 training attendees suggested that confidence of clinicians in supporting this comorbidity increased on average from 46 to 68%. This was supported by qualitative evaluations from the clinical team. |
| Wijker et al. (2020) | Bespoke AAT vs waiting list | **Primary and secondary outcome measures not specified.**  Perceived Stress Scale (PSS): self-report measuring self-perceived stress.  Symptom Checklist-90-Revised (SCL-90-R): self-report measuring of psychological and physical symptoms.  Rosenberg Self-Esteem Scale (RSES): self-report measuring self-esteem. | Not applicable | **Feasibility:**  - *Intervention attendance rates:* All participants in the intervention group took part in at least 9/10 intervention sessions. The adherence rate was 98%  - *Intervention drop-out rates:* No participants dropped out of the intervention arm, one participant from the intervention arm was excluded from analysis due to being identified as an outlier, sever personal issues unrelated to the study. | Mental health outcome using PSS; SCL-90-R; & RSES:  *-* There was no significant time x group interaction effect for *Perceived stress* (PSS, estimated effect with potential covariates: -3.3; 95% CI: -6.1 to -0.5; p = 0.020; d=0.53).  - There was no significant time x group interaction effect for *psychological and physical symptoms* scores (SCL-90-R: -14.7; 95% CI: -30.8 to 1.4; p = 0.072).  - There was no significant time x group interaction effect for *self-esteem* (RSES: 0.8, 95% CI −1.3 to 2.9; p=0.440).  - Results were consistent between models in which covariates were adjusted for (age, gender, having a dog at home at baseline and WAIS total IQ) and unadjusted models. Mixed models adjusted for covariates showed small significant intervention effects on the *agoraphobia subscale of the SCL-90-R*. There was no significant effect on the *depression subscale of the SCL-90-R* in the model adjusted for the covariates (d = 0.33; p = 0.055).  For all tested models, there were no improvements in model fit when the interaction term (time-points x intervention condition) was added, indicating no significant difference in intervention effect for post-intervention compared to 20-week post-baseline follow-up. |
| Wise et al. (2019) | Adapted CBT for anxiety | **Primary and secondary outcome measures not specified.** **Hamilton Anxiety Scale (HAM-A) - clinician-rated measure assessing anxiety symptoms.**  Hamilton Depression Scale (HAM-D) - clinician-rated measure of depressive symptoms.  Clinical Global Impression-Severity (CGI-S) - clinician-rated measure of severity of psychopathology.  Clinical Global Impression-Improvement (CGII) - clinician-rated measure of treatment response.  Beck Anxiety Inventory (BAI) - self-report measure assessing the presence and severity of anxiety. | **Communication accommodations**  - Use of simple, written material and visual aids  **Accommodate individual differences**  - Encourage individual's hobbies and interests  **Intervention structure**  - Family/caregiver/other involvement  **Intervention content**  - Integration of emotion-focused strategies  Integration of cognitive-behavioural approaches  - Integration of social skills training | **Feasibility**  - *Intervention drop-out rate:* One participant dropped out at the mid-assessment point. | **Effect of time (no comparison group).**  Mental health outcomes using HAM-A; HAM-D; CGI-S; CGI-I; & BAI:  - CGI-S scores were significantly lower from baseline (M = 4.86, SD = 0.69) to post-treatment (M = 2.71, SD = 1.11), p < 0.001.  - Among participants (n = 7), including the one participant only completing treatment until the mid-assessment, four of the participants were classified as treatment responders (57%) based on report on the CGI-I at post-assessment.  - Results indicated a significant reduction in clinician-rated symptoms of anxiety on the HAM-A from baseline (M = 22.14, SD = 3.02) to post-treatment (M = 14.57, SD = 5.06), p < .006, (large effect).  - Clinician-rated symptoms of depression (HAM-D) did not reveal clinically significant reductions from pre-treatment (M = 6.71, SD = 5.61) to post-treatment (M = 5.71, SD = 3.40), p = .60.  - Analysis of participant reports of anxiety (BAI) did not reveal significant differences from pre-treatment (M = 12.33, SD = 7.29) to post-treatment (M = 9.33, SD = 5.85), p = .28. |

*Note.* **CBT** = Cognitive behavioural therapy, **EMDR** = Eye movement desensitisation and reprocessing, **M** = mean, **MBT-AS** = Mindfulness-based therapy for autism spectrum disorders, **NHS**= National Health Service, **OCD** = obsessive compulsive disorder, **RTSM** = Real-time stress management, **SD** = standard deviation, **T** = timepoint, **TAU** = Treatment as usual.

**Table S12.** Predictors of outcome (*N* = 4)

| **Authors (year)** | **Description of strategy** | **Adaptation (sub)categories** | **Predictors of outcome** |
| --- | --- | --- | --- |
| Bemmer et al., 2021 | Adapted group Cognitive Behavioural Therapy (CBT) for social anxiety. | **Intervention content**  - Simplified and structured content  - Integration of cognitive-behavioural approaches  - Integration of social skills training | **Demographics** (age, sex, Wechsler Test of Adult Reading IQ estimate and Autism Diagnostic Observation Schedule-2 total scores) and **pre-intervention social functioning** did not statistically significantly predict changes on the Liebowitz Social Anxiety Scale—Self-Report (LSAS-SR) total score (p=0.421). |
| Blainey et al., 2017 | Adapted individual Cognitive Behavioural Therapy (CBT). | **Communication accommodations**  - Clear communication  - Use of simple, written material and visual aids  **Accommodate individual differences**  - Tailor practice to individual needs and preferences  **Intervention structure**  - Format of intervention  - Family/caregiver/other involvement  **Intervention content**  - Integration of social skills training | The individual difference factors (**attendance of therapy sessions, gender, age that the individual was diagnosed with ASD, presence of co-occurring ADHD, depression or anxiety disorder and the nature of other presenting problems including aspects of personality disorder, anger and psychotic features**) were shown to have no effect (p > 0.05) on change in general distress (Clinical Outcomes in Routine Evaluation-Outcome scores) over the course of therapy. |
| Russell et al., 2013 | Adapted individual CBT for OCD | **Communication accommodations**  - Use of simple and preferred language  - Use of simple, written material and visual aids  **Intervention content**  - Simplified and structured content  - Integration of emotion-focused strategies  - Integration of cognitive-behavioural approaches | **Age, verbal IQ, and Autism Diagnostic Observation Schedule scores** were not statistically significantly associated with treatment outcome (the percentage change in total OCD severity scores). |
| Sizoo et al., 2017 | Adapted group CBT and adapted mindfulness-based stress reduction | **Communication accommodations**  - Plan in advance  - Clear communication  - Use of simple and preferred language  **Intervention content**  - Taking it slow | **Age** and **sex** were not statistically significant predictors for the treatment effect on anxiety and depression in both CBT and adapted mindfulness-based stress reduction groups. |
